# Age-specific Contribution of Contacts to Transmission of SARS-CoV-2 in Germany

**DOI:** 10.1101/2021.12.13.21267716

**Authors:** I. Rodiah, P. Vanella, A. Kuhlmann, V. K. Jaeger, M. Harries, G. Krause, W. Bock, B. Lange

## Abstract

**Introduction:** Current estimates of pandemic spread using infectious disease models in Germany for SARS-CoV-2 often do not use age-specific infection parameters and are not always based on known contact matrices of the population. They also do not usually include setting-based information of reported cases and do not account for age-specific underdetection of reported cases. Here, we report likely pandemic spread using an age-structured model to understand the age- and setting-specific contribution of contacts to transmission during all phases of the COVID-19 pandemic in Germany.

**Methods:** We developed a deterministic SEIRS model using a pre-pandemic contact matrix. The model is optimized to fit reported age-specific SARS-CoV-2 incidences from the Robert Koch Institute, includes information on setting-specific reported cases in schools and integrates age and pandemic period-specific parameters for underdetection of reported cases deduced from a large population-based seroprevalence study.

**Results:** We showed that taking underreporting into account, younger adults and teenagers are the main contributors to infections during the first three pandemic waves in Germany. Overall, the contribution of contacts in schools to the total cases in the population was below 10% during the third wave.

**Discussion:** Accounting for the pandemic phase and age-specific underreporting seems important to correctly identify those parts of the population where quarantine, testing, vaccination, and contact-reduction measures are likely to be most effective and efficient. In the future, we will aim to compare current model estimates with currently emerging during-pandemic age-specific contact survey data.

## Introduction

Epidemiological models have been essential to predict the spread of SARS-CoV-2 and are frequently used to inform decision-makers on the effectiveness of interventions. Many estimations in projections and scenario modeling use compartment models that divide a population by health status and assume transition rates from one health status to another [2]. Simulations are based on assumptions or data about the probability of disease transmission, incubation period, recovery rates, and mortality rates. Based on deterministic differential equations, multiple studies have modeled the current spread of SARS-CoV-2 (e.g. [9, 21, 27]). Some models consider age groups [11, 21, 22, 27, 29, 31], are agent-based [24, 37], or include mobility [30]. Others consider additional disease compartments [21] and vaccination [31]. However, current models in Germany are not sufficiently accounting for age-specific estimates of disease severity and underdetection of reported cases, which leads to underestimation as well as overestimation of the contribution of contacts in different age and population groups to infection dynamics and deaths [39].

The population’s age structure is known as one of the key determinants of acute respiratory diseases, especially when it comes to infection severity. For instance, children are considered to be responsible for a large part of the transmission of influenza [4] but the majority of hospitalizations and deaths occur among people of ages over 65 years [25, 34]. Similarly, COVID-19 mortality among people who have been tested positive for the SARS-CoV-2 is substantially higher in older age groups and near zero for young children [6, 36, 38]. Moreover, the infectiousness of an individual has been reported to vary with time after infection [12, 16], which is known to affect epidemic spread [17, 20]. As for all respiratory virus infections, contact patterns in the population are also relevant.

Underdetection of actual infection activity by notified case reports to authorities is a well-known limitation, but not often included in modeling efforts, even less using age-specific underreporting estimates [21, 30]. For Germany, population-based studies suggest that actual infection activity is heterogeneous across regions, time points, and age groups [13]. However, both age and underdetection of infections are highly relevant for predicting regional infection events, especially for different contact areas and for estimating the effectiveness of interventions [15] as well as for predicting hospitalizations and deaths correctly.

In the work presented here, we incorporate age-specific underdetection ratios in a classical age-specific infection model and analyze the impact on transmission dynamics. The model has a circular structure, which allows for reinfections, long-term complications, and delayed deaths. Moreover, we consider the social contact patterns since it impacts the spread of disease. Our model allows us to assess the age-dependent contribution of contacts to the transmission of COVID-19 in Germany. We assess the sensitivity of infection transmission by applying underdetection ratios from large population-based seroprevalence studies of adults [13] and children [18, 35]. We then analyze and compare the transmission dynamics across age groups during the first three infection waves in Germany, taking underdetection into account. We also estimate time-dependent transmission rates and the contribution of contacts within the school setting for periods where specific data on schools are available [44].

## Data and Methods

### Data

We considered data on officially reported SARS-CoV-2 infections by the German health institute [43] (Robert Koch Institute; RKI). The age distribution of the German population is obtained from data for 2020 reported by the Federal Statistical Office [41]. We used weekly age-specific reported data of SARS-CoV-2 infections during the first three infection waves in Germany, i.e. calendar week 5 (starting on 27 January) 2020 to calendar week (starting on 23 June) 20 of 2021. Weekly reported SARS-CoV-2 infections by age group are shown in Supplementary A – Figure 1.

To investigate the contribution of school contacts, we use the weekly reported SARS-CoV-2 infections among students and teachers from the Standing Conference of Ministers of Education and Cultural Affairs website (KMK) [44]. The data used are from calendar week 8 to 20 of 2021 (Supplementary A – Figure 2). We assumed that the proportion of new infections in schools for different age groups is determined by the proportion of student and teacher numbers reported by the Federal Ministry of Education and Research [40] and the Federal Statistical Office [42].

Based on pandemic phase- and age-specific underdetection ratios derived from population-based studies among adults [13] and children [18, 19, 35], we estimated average age-specific underdetection ratios for different phases of the pandemic in Germany (see Table 1), which we implemented in our model. Gornyk et al [13] investigated seroprevalence estimates for SARS-CoV-2 that indicated an age-specific underreporting ratio (infected to reported) of COVID-19 transmission in a large (>25000 participants, seven large regions in Germany) population-based seroprevalence study and we used their estimates for age-specific underdetection ratios in adults. For children, we used seroprevalence studies available for the south of Germany during the first and second waves [18, 19, 35]. For the third wave, we assumed a significantly reduced level of underdetection in this age group, based on reported cases during the period of the introduction of large-scale testing in child care institutions, as no seroprevalence estimates for children were available. To apply underdetection in our model, we correlated the age-specific underdetection ratios of each stage to the reported number of infected individuals.

**Table 1.** Underdetection ratios by age group for different SARS-CoV-2 waves of transmission (Source: [13, 18, 19, 35])

| Group | First Wave<br>(March – June 2020) | Second Wave<br>(July 2020 – February 2021) | Third Wave<br>(March – May 2021) |
| --- | --- | --- | --- |
| 0 – 4 years | 8 | 6 | 2 |
| 5 – 14 years | 6 | 4 | 1.5 |
| 15 – 34 years | 6 | 4 | 2 |
| 35 – 59 years | 4 | 3 | 2 |
| 60 – 79 years | 4 | 4 | 3 |
| 80+ years | 2 | 1.5 | 1.5 |

Since our age-structured model allows us to adjust the transmission rates among different age groups, we applied social contact patterns to the transmission rates. We used POLYMOD data for Germany, originally made available by Mossong et al [26], to construct a symmetric contact rate matrix by age group consistent with our model. The contact rate *c*_*ij*_ is related to the social contact matrix by

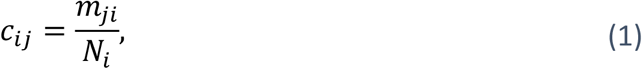

where the element *m*_*ij*_ of the social contact matrix (see Supplementary A – Figure 3) represents the average number of contacts made by an individual in the age group with individuals in the age group *j* during one day and *N*_*i*_ is the population size of age group *i*.

### Model

The developed model is an adaptation of a deterministic SEIRS (Susceptible-Exposed-Infectious-Recovered-Susceptible) model to the specific properties of SARS-CoV-2 infections. It distinguishes healthy (susceptible) individuals, infected but not yet infectious (exposed) individuals, symptomatic and asymptomatic patients. Furthermore, we considered compartments for hospitalizations, patients in the intensive care units (ICUs), and long-COVID (i.e. suffering eventual symptoms after officially recovering from the infection). Last, patients will recover or die. We also assume that there is a reinfection process in the transmission. In our model, we split the recovery compartment into a compartment for those recovered from COVID and a long-COVID compartment, since we assumed that both have a different reinfection rate. Figure 1 illustrates the model structure.

**Figure 1.**
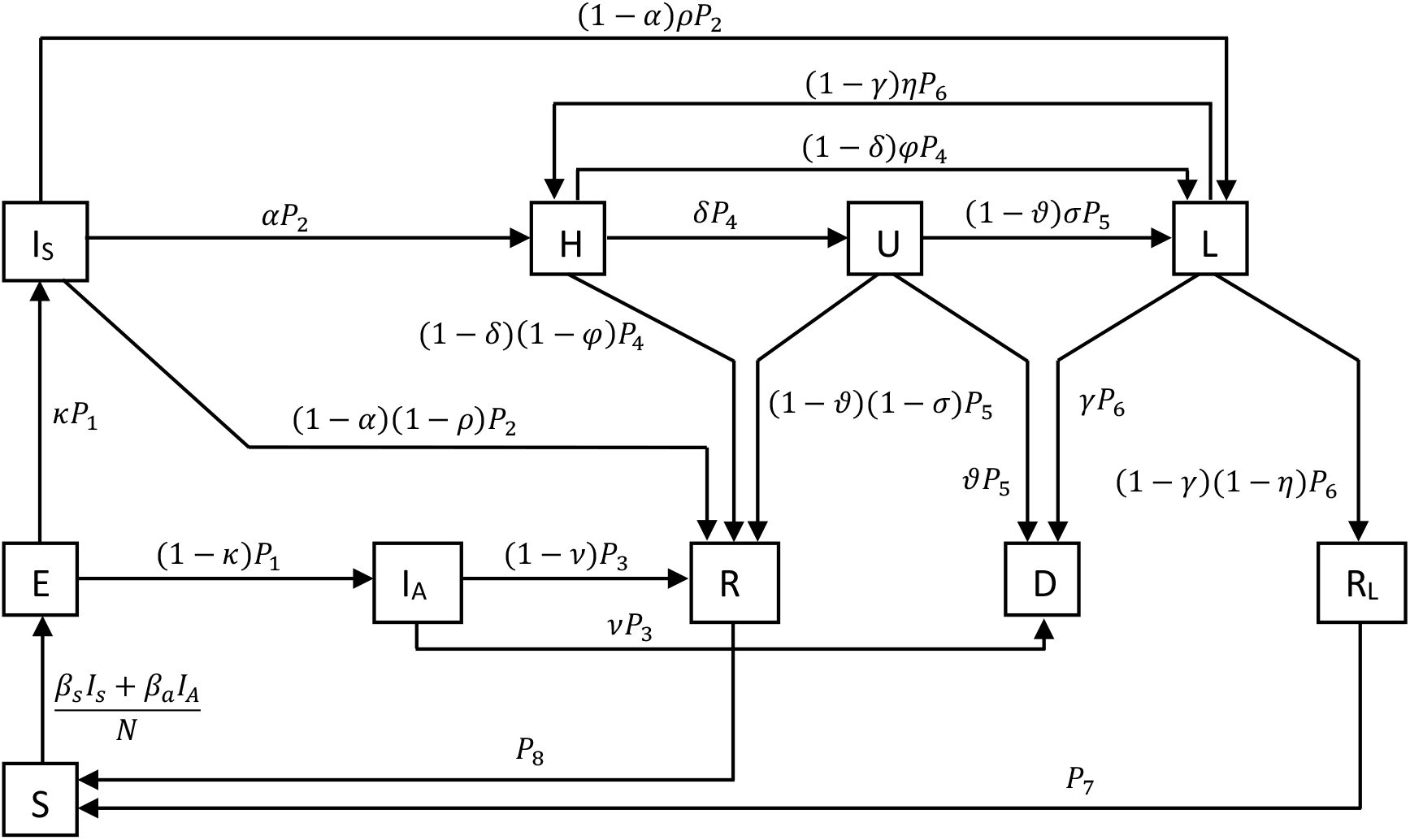
Schematic illustration of the extended SEIRS model

The population is split into six age groups, as given in Table 2, in accordance with data provided by RKI. An individual in age group *i* is classified either as susceptible (*S*_*i*_), exposed (*E*_*i*_), asymptomatically infectious 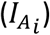, symptomatically infectious 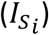, hospitalized (*H*_*i*_), in intensive care (*U*_*i*_), suffering under long-COVID (*L*_*i*_), fully recovered 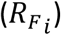, recovered from long-COVID 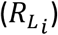, or dead (*D*_*i*_). The population size of age group *i* is given by *N*_*i*_. The developed model is given by the following differential equation system,

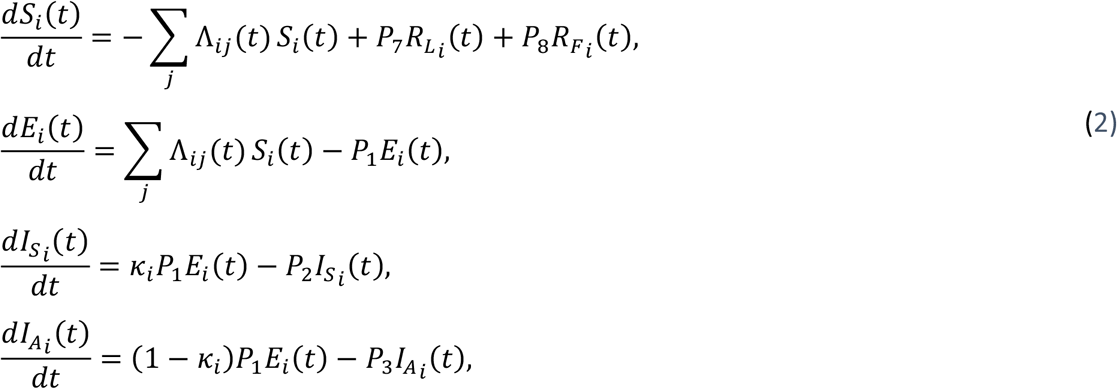

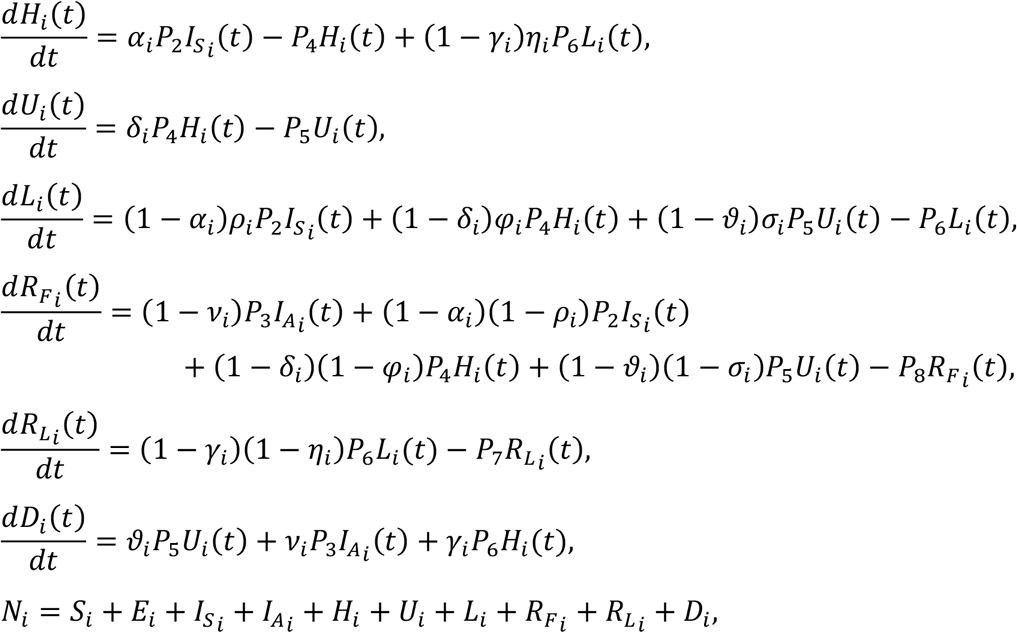

where

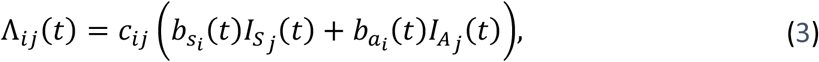

**Table 2.** Age groups in our model

| $i$ | Group |
| --- | --- |
| 1 | 0 – 4 years |
| 2 | 5 – 14 years |
| 3 | 15 – 34 years |
| 4 | 35 – 59 years |
| 5 | 60 – 79 years |
| 6 | 80+ years |

Typically, 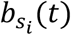 and 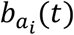 (*t*) denote the time-dependent risk of infection per contact in age group *i* for symptomatically and asymptomatically infectious individuals, respectively. In our model, we called them scaling parameters since we calibrated them to scale the contact matrix to account for the effects of interventions and behavioral change over time. *P*_1_ – *P*_8_ are health state transition rates. The Greek letters denote the transition probabilities. Supplementary B gives an overview of the model parameters.

The basic reproduction number *R*_0_ is defined as the expected number of secondary infections produced by a single infected individual in a completely susceptible population [6], which is used to describe the transmissibility of infections [8]. *R*_0_ can be derived by employing the next generation matrix method [7]. The compartments with infected individuals are divided into two categories, which are the appearance of new infections in the compartment and the transfer of the infected into and out of the compartment. The Jacobian [7] of the transmission matrix *F* and transition matrix *V* describes the generation of new infections and the transfer across compartments. The elements *m*_*ij*_ of *M* = *FV*^-1^ are related to the expected number of secondary infections in compartment *i* caused by an infected individual of compartment *j*. During an epidemic, susceptible individuals gradually become infected. Therefore, the effective reproduction number is defined as a time-varying variable that quantifies the instantaneous transmissibility of infections. It also includes the effects of interventions and behavioral changes. The effective reproduction number *R*_*t*_ for the age-structured model is the dominant eigenvalue of the matrix *M* given by

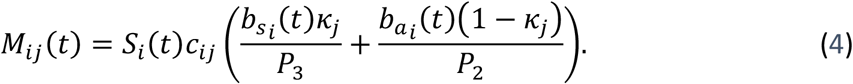

Here, *R*_*t*_ is estimated numerically for a set of parameters and a susceptible population.

### Parameter Estimation

The model parameters are fit by ordinary least squares (OLS). The time-dependent scaling parameters (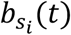 and 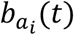) are estimated by calibrating them weekly to age-specific reported new SARS-CoV-2 infections. Based on the fitting results, we observe the estimated force of infection corresponding to the age groups. The time-dependent marginal force of infection in age group *i* with respect to contacts with age group *j* are estimated by matrices obtained from equation (3). The force of infection by age group is the row sum of matrices (3) over age group *j*. We also estimated the effective reproduction number over time by calculating equation (4) using the estimated scaling parameters.

Inserting the fitted parameters into the model, we predicted the number of new cases by age group. In addition, we used the same method to fit the fatality rate and predict the number of deaths. To evaluate the model, we used the weighted interval score (WIS) by Bracher et al [5], which is one of the evaluation parameters used in the European COVID-19 Forecast Hub [39].

### Estimation of the age-dependent contribution

By applying social contact patterns and the time-dependent scaling parameters among the different age groups into our model, we estimated the number of new cases in the age group *i* generated by each contact in the age group *j* over time through matrices obtained by multiplying equation (3) with the number of susceptibles by age group,

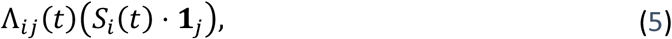

where 1_*j*_ denotes a row vector of ones. This allows us to investigate among whom COVID-19 spreads over time. The results show the contribution of contacts to transmission in the overall population. We divided the observation time into three phases representing each wave of infection in Germany. Therefore, we were able to analyze the influence of age-specific contacts in the infection dynamics of individuals of different age groups at the national level from a temporal perspective.

### Implementation of the school setting

For the implementation of the school setting, we split the population into nine age groups, as given in Table 3.

**Table 3.** Age groups in model of school setting

| $i$ | Group |
| --- | --- |
| 1 | 0 – 4 years |
| 2 | 5 – 9 years |
| 3 | 10 – 14 years |
| 4 | 15 – 19 years |
| 5 | 20 – 24 years |
| 6 | 25 – 29 years |
| 7 | 30 – 59 years |
| 8 | 60 – 79 years |
| 9 | 80+ years |

We assumed that the age groups 2 – 8 have direct relevance in the school context (as student and teaching staff). Those are divided into the subgroups “non-school” or “school”. Using OLS, we calibrated our model with the weekly reported COVID-19 cases among students and teachers in schools. The estimated forces of infection in schools are calculated via equation using the school contact matrix by POLYMOD and the number of infections in the school.

### Implementation of underdetection of infections by reported cases

We estimated the true new infections by multiplying the weekly age-specific reported cases of COVID-19 with the age-specific underdetection ratios from Table 1,

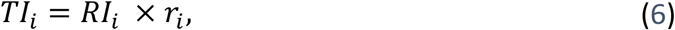

where *r*_*i*_ is the underdetection ratio of age group *i, TI*_*i*_ and *RI*_*i*_ denote the estimation of truly new and reported infected of age group *i*, respectively. Then, we used the same method for fitting data to estimate the time-dependent transmission parameters among different age groups. We investigated the contribution of contacts to transmission, accounting for the underdetection in the overall population. Thus, we can analyze the sensitivity of infection transmission by applying underdetection ratios for Germany.

## Results

### Age-dependent contribution of contacts to cases

Figure 2 shows the time-dependent scaling parameters by age group derived from our model over calendar week 5 of 2020 to calendar week 20 of 2021 in Germany. During the first wave, the scaling parameters were highest for all age groups when the spread of infection began. In the second and third waves, the scaling parameters remained stationary for each age group, except for the elderly age group of 80 years and above. The scaling parameter for the elderly increased in the second wave and then decreased in the third wave. Figure 3a shows the effective reproduction number *R*_*t*_, calculated by fitting parameters according to the age groups. The value of *R*_*t*_ was estimated to be around 6.9 in calendar week 9 of 2020 when the spread of infection began in the first wave. At the beginning of calendar week 12 of 2020, *R*_*t*_ decreased below 2. The *R*_*t*_ was estimated to be in the range of 0.7 – 2 in the second wave and between 0.6 – 1.4 in the third wave.

**Figure 2.**
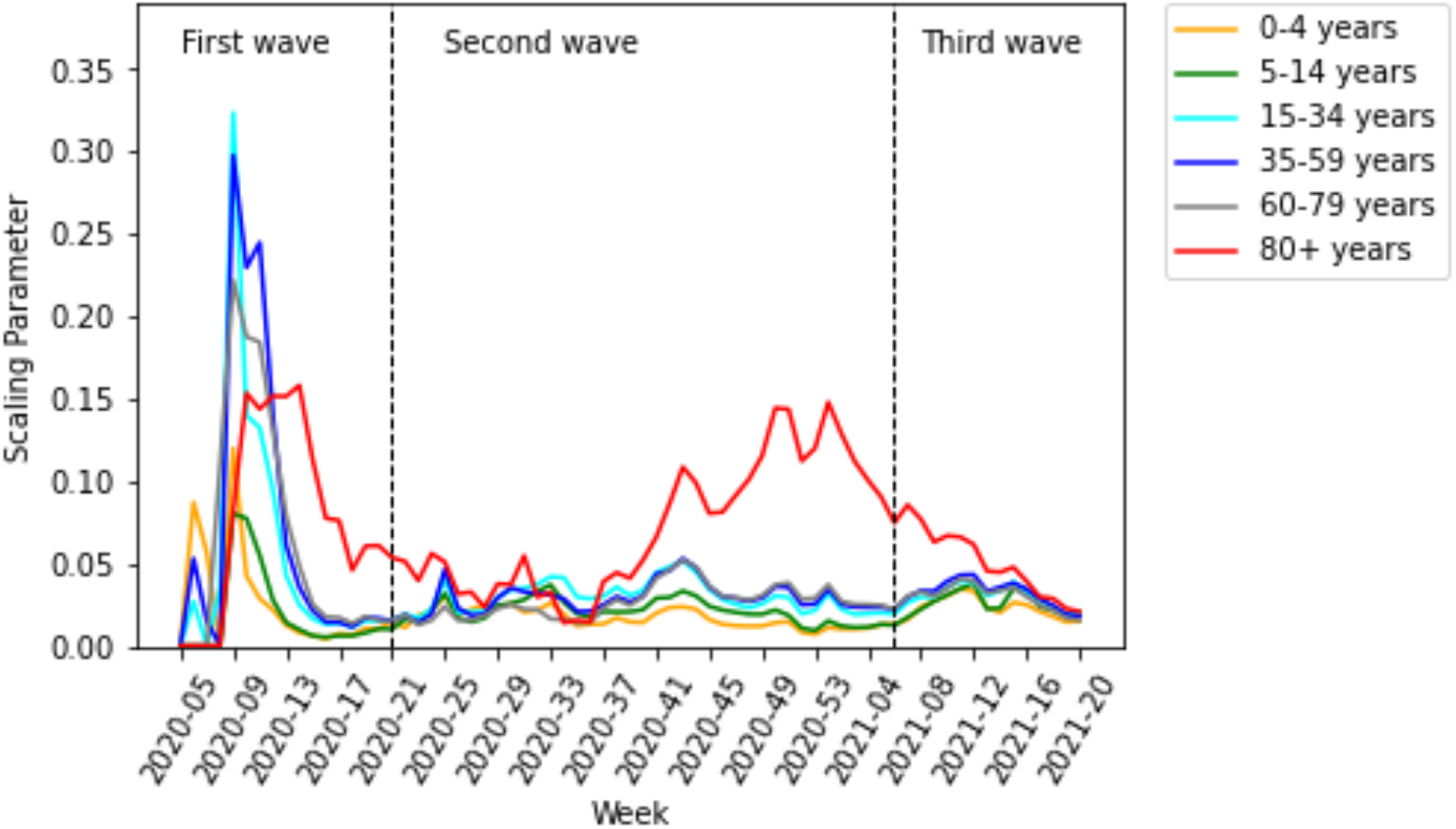
Estimated weekly scaling parameters per contact by age group

**Figure 3.**
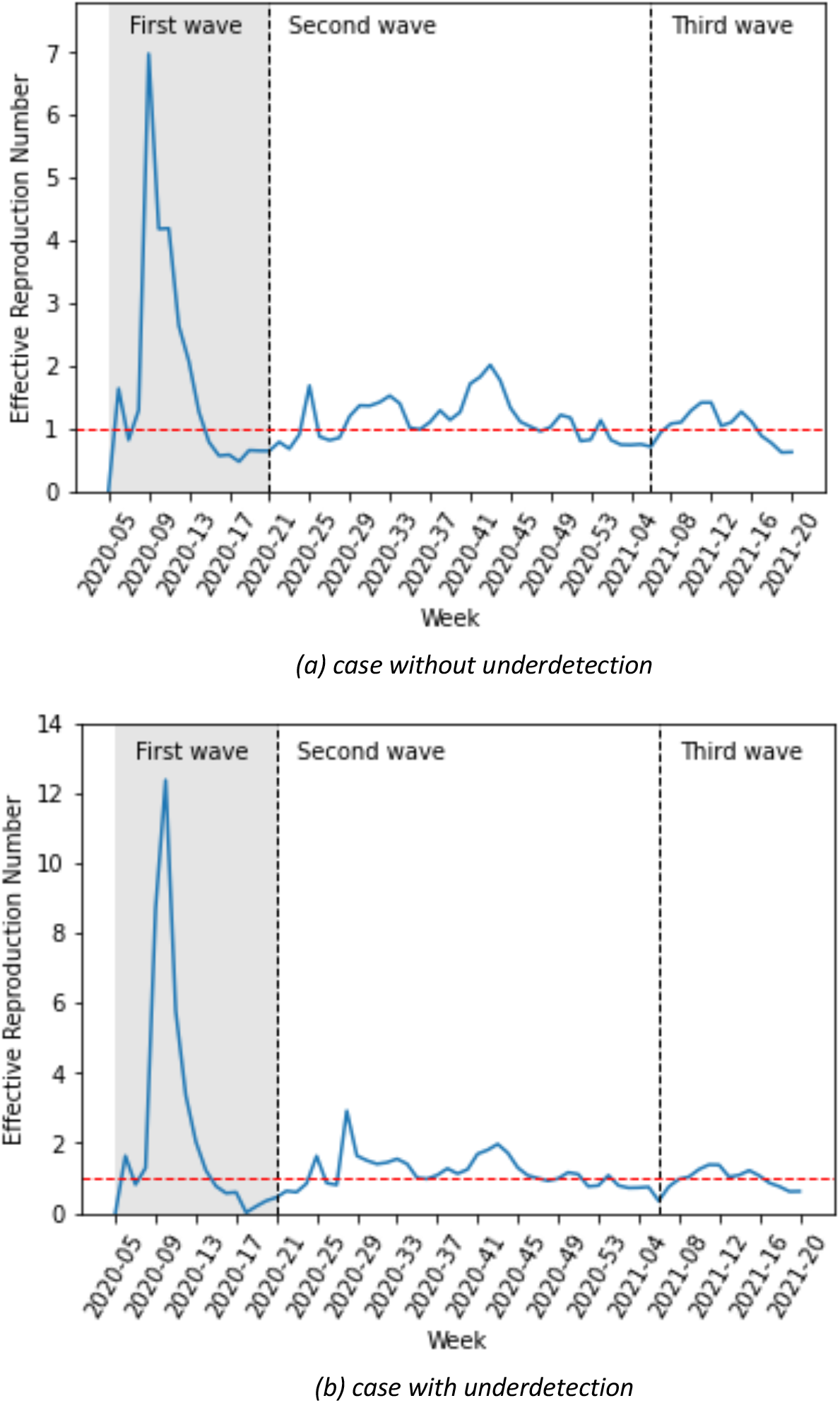
The effective reproduction number is calculated by fitting parameters for case (a) without and (b) with underdetection In the first few weeks of the first wave, there were artifacts due to a low number of cases, increased testing, etc. Therefore, the results of weekly calibration are unrealizable during the first few weeks.

Here, we described the estimated marginal force of infection, which is the individual contribution of contacts to the age-specific transmission rate. More detailed results are shown in heat maps in Supplementary C. The element of a matrix represents an estimator for the force that an individual in the age group on the vertical axis will become infected from individuals in the age group on the horizontal axis. We also showed the estimated contributions of contacts to the transmission over time in heat maps in Supplementary D. Some contributions of contacts for the transmission at a fixed time for all waves are shown in Figure 4. The element of a matrix represents an estimated number of infected in the age group on the vertical axis due to contact with individuals in the age group on the horizontal axis. For instance, in Figure 4b, we estimated that there were about 18800 new infections aged 35 – 59 years due to contacts with individuals aged 15 – 35 years in week 51 of 2020.

**Figure 4.**
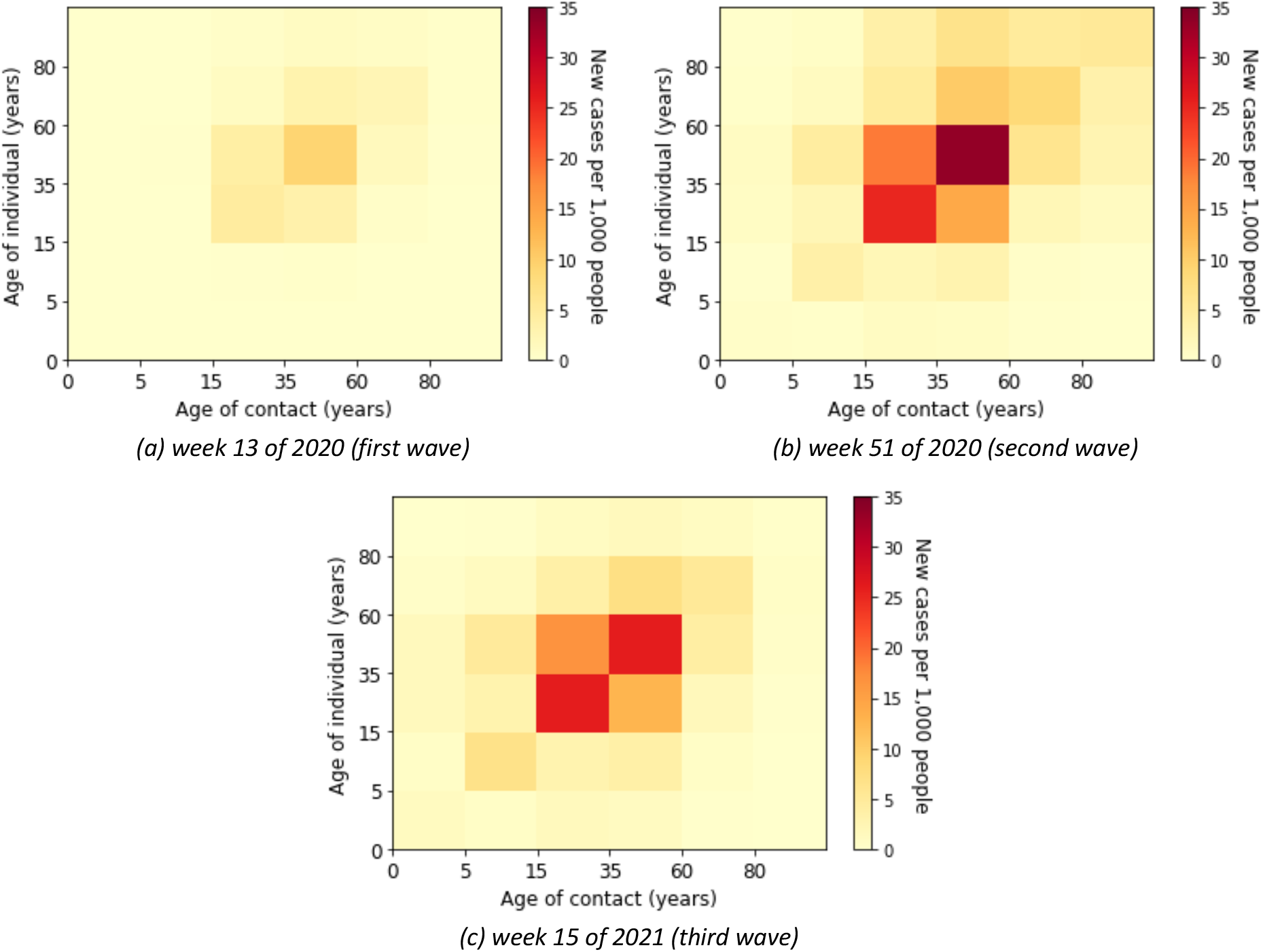
Estimated absolute contribution of contacts to the transmission

### First wave

The individual contribution of contacts to the overall transmission rates was predominantly from contacts in young adults, adults, and the elderly (see Supplementary C – Figure 4) in the first wave, i.e. weeks 5 until 21 of 2020. The absolute contribution of contacts to the transmission was predominantly in the adult group (Supplementary D – Figure 7).

### Second wave

In the second wave, the reported number of cases was very high. The age-specific forces of infection by age group are shown in Supplementary C – Figure 5. The individual contribution of contacts to the transmission rates for the second wave occurred predominantly in young adults until elderly groups. In weeks 50 of 2020 until 2 of 2021, the contribution to the transmission was highest in the elderly that had contact with the other age groups above 14 years. In Supplementary D – Figure 8, we show that the absolute contribution of contacts to the transmission was mainly in the young adult and adult groups.

### Third wave

The age-specific forces of infection by age group for the third wave (i.e. weeks 7 until 20 of 2021) are shown in heat maps in Supplementary C – Figure 6. Different from the first and second waves, the individual contribution of contacts to the transmission rates occurred predominantly in children and young adults. The absolute contribution of contacts to the transmission in the third wave shows rather similar trends as in the second wave (see Supplementary D – Figure 9).

Figure 5a shows the estimated force of infection by age group. In the first wave, the trends of the adult age groups appear to be homogeneous. The peak of transmission in elderly groups in the first wave appeared with a delay in comparison to the other age groups. However, they differed across adult age groups in the third wave. The trends of the children’s group in the third wave are higher than in the previous wave. In contrast, the trends of the elderly groups are lower in the third wave than in the previous wave.

**Figure 5.**
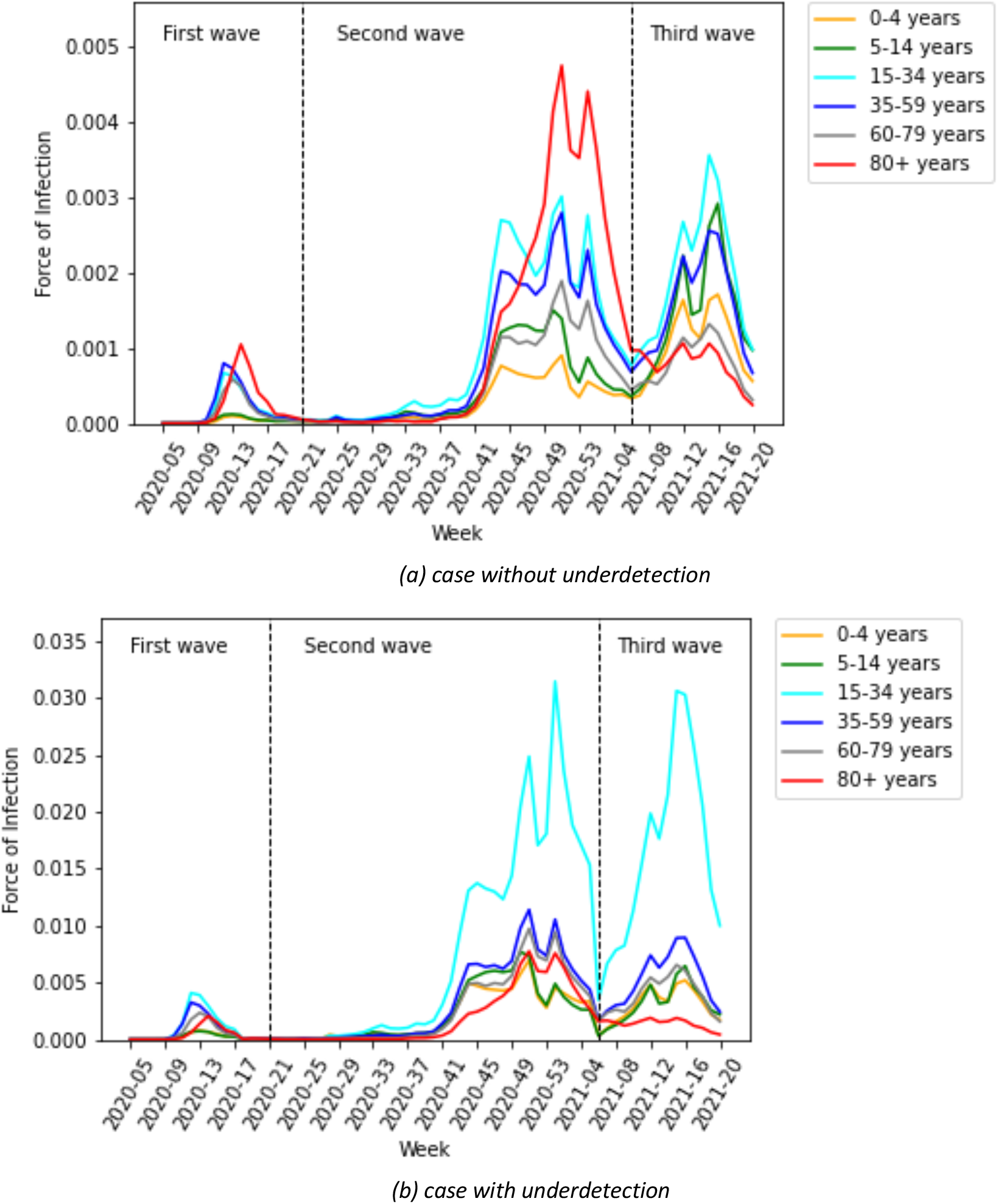
Estimated force of infection by age group for case (a) without and (b) with underdetection

### Contribution of age-dependent contacts to cases accounting for underdetection

The effective reproduction number in the case of underdetection is shown in Figure 3b. The trend of *R*_*t*_ for underdetection appears to be similar to the case without underreporting. Almost every time, the value of *R*_*t*_ for underdetection is smaller, except at certain times. At the beginning of the first wave, the value of *R*_*t*_ for underdetection is higher. In calendar week 9 of 2020, the estimation of *R*_*t*_, when accounting for underreporting, is 1.9 times higher. The value of *R*_*t*_ is also higher at calendar week 28 of the second wave. At the other times, the estimation of *R*_*t*_ ranges between 0.7 – 2.9 in the second wave and between 0.35 – 1.4 in the third wave.

The individual forces of infection by age group are shown in heat maps in Supplementary E. The estimated absolute contributions of contacts to the transmission over time in heat maps are shown in Supplementary F. Accounting for the underdetection, Figure 6 shows some absolute contributions of contacts to the transmission at a fixed time for all waves. The figure has the same interpretation as Figure 4, however, with different scales. We can observe that the dominant contribution of contacts to the transmission is shifted to the younger age group (young adult group), which is due to underdetection. In Figure 5b, we show the estimated force of infection for the underreported cases. The forces of infection by age group differ significantly from the case without underreporting. Here, the estimated force of infection is much higher in the young adult group than in the other age groups in the second and third waves. In addition, the estimated force of infection in the elderly is lower in the second wave than in the case without underreporting. The delay of an increasing force of infection in older age groups is more pronounced in Figure 5b.

**Figure 6.**
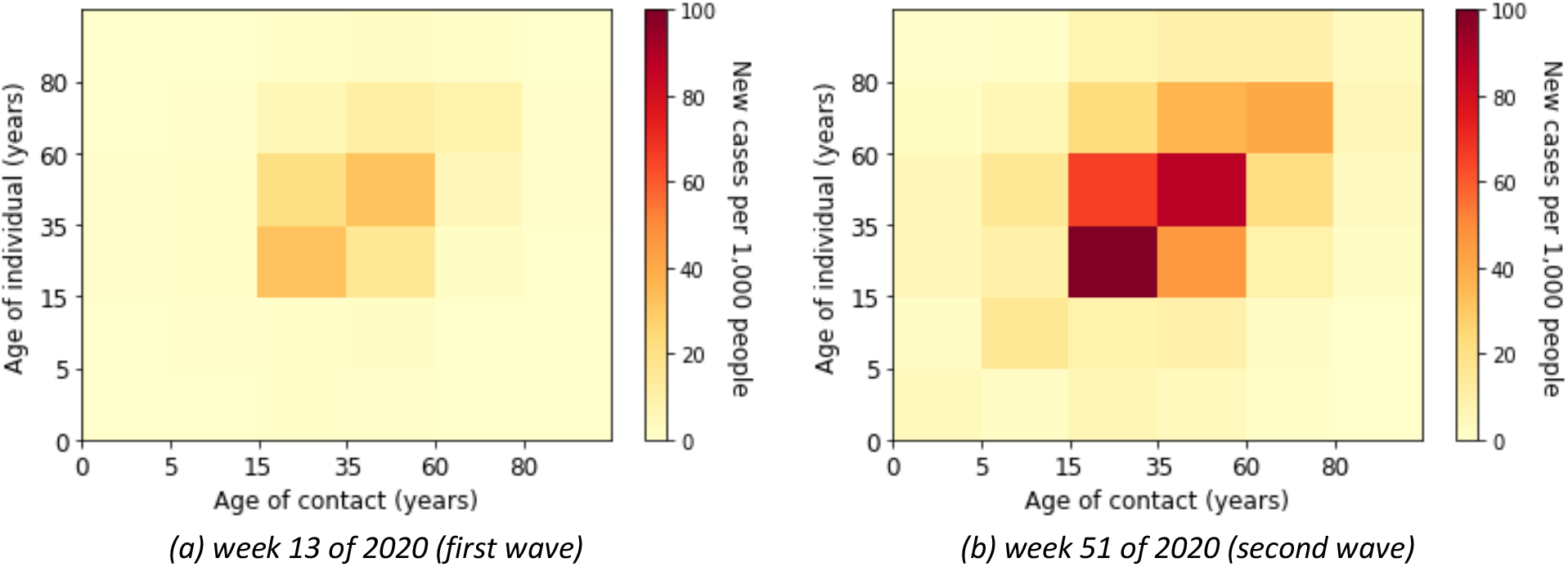
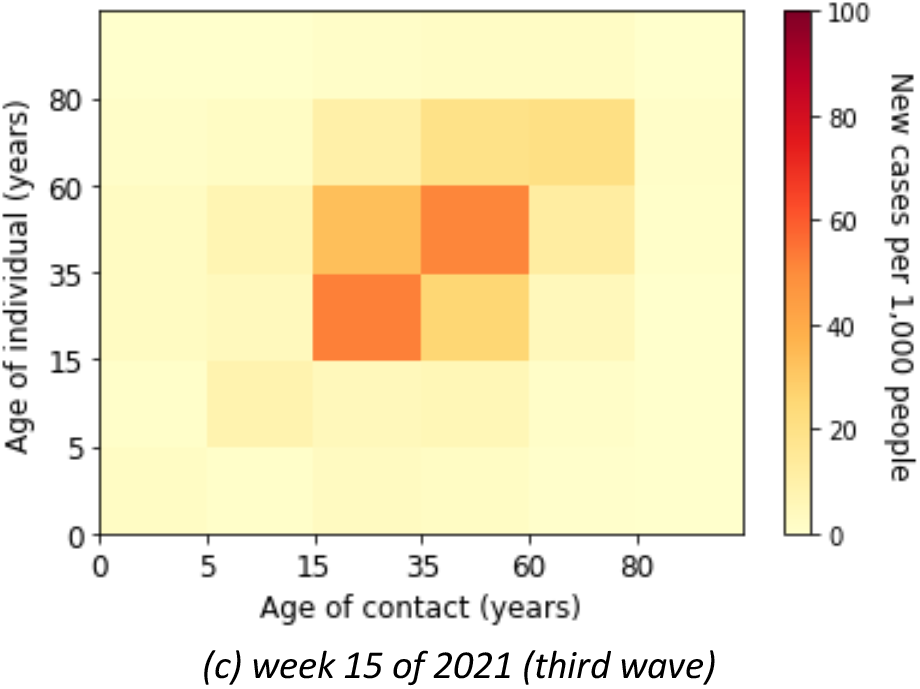
Estimated absolute contribution of contacts to the transmission accounting for underdetection

### Contribution of school setting-dependent contacts to cases

Here, we investigated the contribution of schools to the transmission of COVID-19 during the third wave in the German population. Using reported data in schools and a school contact matrix, we obtained the estimated forces of infection in schools by age group in Figure 7a. There is a clear trough in calendar weeks 13 – 14 of 2021 due to the Easter holiday. Otherwise, trends appear to be very heterogeneous across age groups. We show the estimated proportion of infection due to contacts with infected people in schools in Figure 8a. The proportion of the contribution of contacts in schools to the overall cases in the population during the third wave is less than 10%. Accounting for underdetection ratios in the third wave, the estimated forces of infection in schools by age group are illustrated in Figure 7b. The force of infection, accounting for the underdetection in the age group 5 – 9 years, was lower than for the case without underdetection. Meanwhile, the forces of infection accounting for underdetection in the age groups 25 – 29 years and 30 – 59 years were higher for than the case without underdetection. Figure 8b shows the proportion of the contribution of contacts in schools with underdetection ratios to the overall cases in the population.

**Figure 7.**
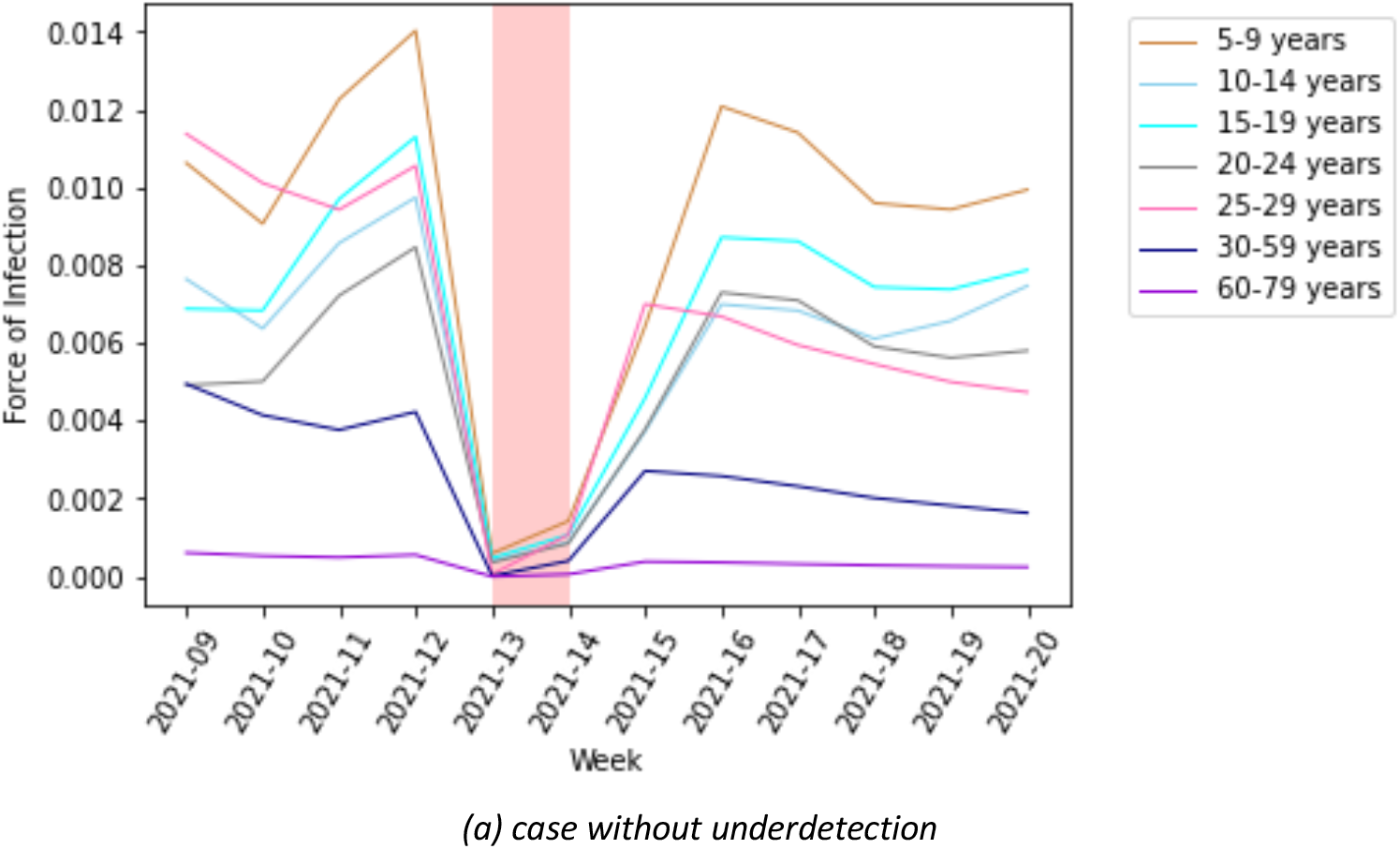
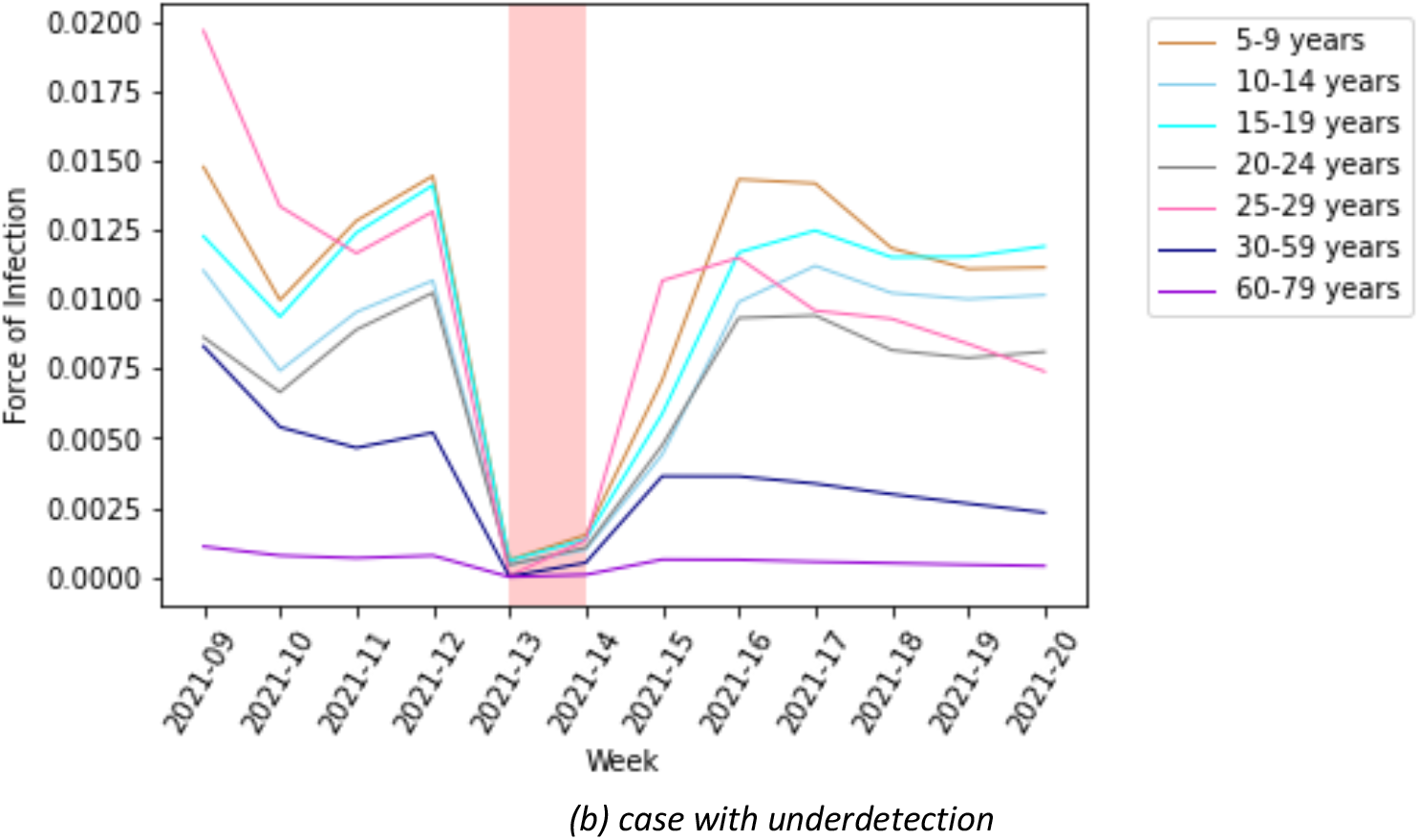
Estimated force of infection in schools by age group in the third wave for case (a) without and (b) with underdetection

**Figure 8.**
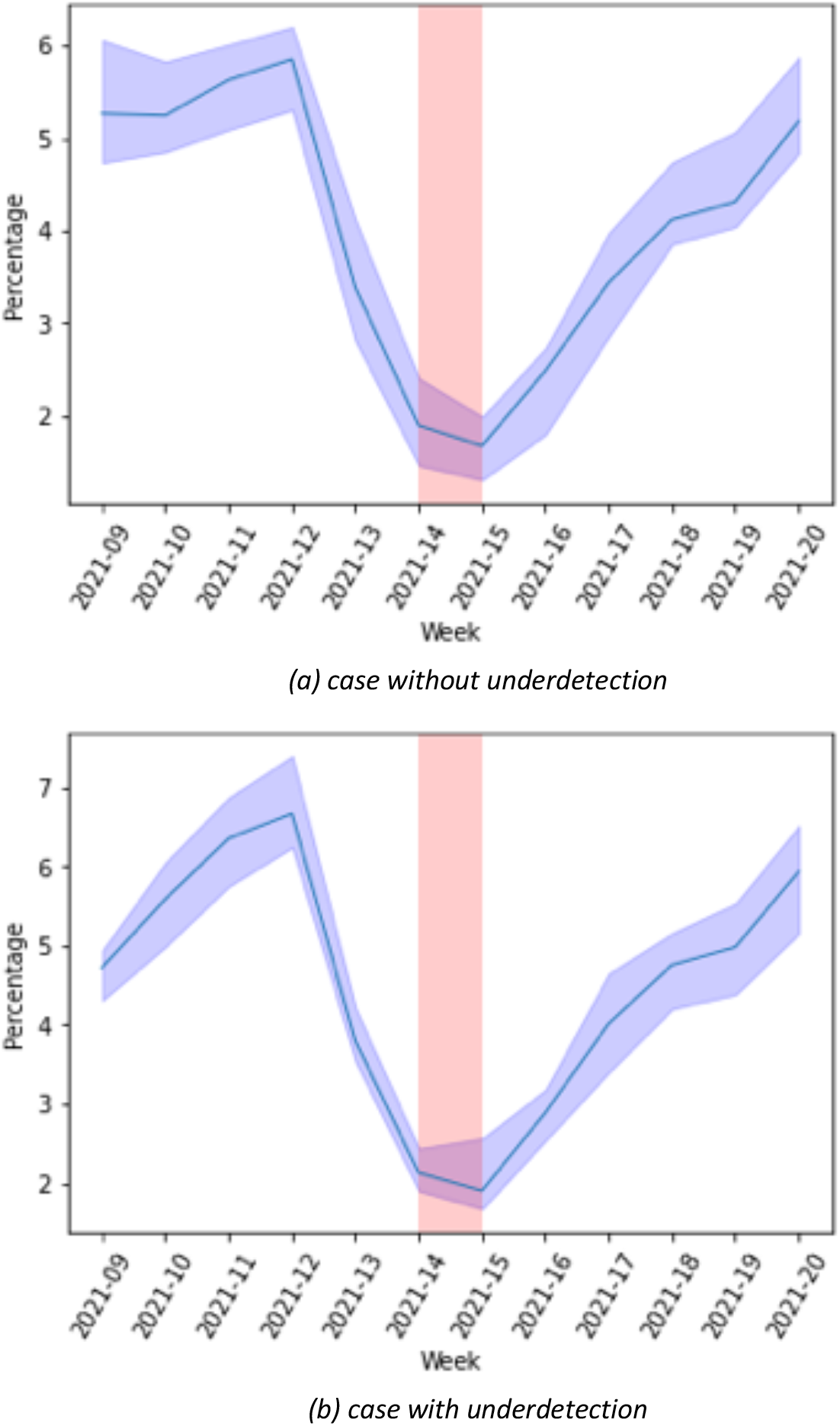
Estimated proportion of infection (CI: 95%) due to contact with infected people in schools in the third wave for case (a) without and (b) with underdetection

## Discussion

We presented estimations on the course of the pandemic based on an age-specific compartmental model that accounts for differential underreporting and assesses the contribution of age- and school setting-specific contacts to overall transmission throughout the pandemic in Germany.

The value of the effective reproduction number in the first wave period may appear rather high compared to the other periods. An obvious reason for the high reproduction number in the first wave could have been caused by one or several super-spreading incidents, such as a carnival event [14]. In addition, there were artifacts due to increased testing as the first wave progressed and low case numbers. The decrease in the effective reproduction number during the first three infection waves may indicate a reduction in contacts in the population, for example, due to the success of the non-pharmaceutical interventions. In the third wave, the force of infection in elderly groups to the transmission was much lower due to the impact of vaccination and potentially higher natural infection acquisition.

The contribution of contacts to the overall transmission across all phases of the pandemic varied but occurred predominantly in the adult group if not accounting for underdetection. Accounting for underreporting, however, we identified a substantial part of infections after contacts in the young adult and teenage groups and – compared to estimates not accounting for underreporting – a higher proportion of infections in the overall population was due to contacts in children and teens.

The difference between model estimations using underreporting and not using underreporting is decreasing towards the third wave with higher use of testing strategies targeted towards younger populations such as working age and school-age populations in Germany [1].

To understand pandemic spread in Germany, estimations assessing detailed regional spread have been provided for both the first and second waves. Lippold et al have used county-level data to provide predictions for future spread [23]. Doblhammer et al showed that during the second wave, political affiliations and socioeconomic indicators were associated with higher incidences of SARS-CoV-2 [10]. A recent estimation of spreading dynamics of SARS-CoV-2 in Germany assessed regional heterogeneity in the effectiveness of measures on infection dynamics and identified three distinct regional clusters of spreading patterns [32].

We believe that the large differences in model estimations integrating age-specific underreporting and those not from our assessments indicate that integrating age-specific underreporting of cases would benefit such efforts, in particular, if the effectiveness of interventions or predictions are based on these. In particular, for assessments of the current epidemiological situation, these estimates should be integrated to be able to capture recent epidemiological situations. Accounting for underreporting needs to be done in an age-specific manner. Modeling without accounting for demographics likely underestimates infection activity in younger adults and relatively young populations significantly. The underestimation of the contribution from the young adult group may provide a larger explanation for the overshoot of epidemic activity in Germany in the older age groups in both the second and fourth waves. In both waves, Germany saw relatively higher than expected epidemic activity overall starting with high epidemic activity in the younger age groups in summer. If this epidemic activity was relevantly underdetected (more than in the other age groups) this would explain the quicker than expected increase in cases and deaths in autumn as an additional explanation to the known seasonality of SARS-CoV-2. This would indicate that screening, testing, and contact tracing in this younger age group (young adults and teens) is of particular importance.

Including cases that are known to have been detected in schools, we estimated the contribution of contacts in schools to be quite volatile, yet below 10% during calendar weeks 10 to 22 in 2021. Interestingly in contrast to what we found for age groups, this estimate was not highly affected by including underdetection estimates. This is possibly due to the mixing of age groups in this setting, lowering the underdetection effect. Also, the lack of an effect of underreporting estimates is probably caused by us not being able to include information for the third wave which in Germany had overall low underdetection and not as high age-specific underdetection differences due to testing strategies both at the workplace and in schools. Information on cases in schools is not available for previous calendar weeks. To our knowledge, no other estimations of the contribution of contacts to cases in the population from the existing data in Germany have been performed. Estimates of reduction of transmission by school closures [3, 33] for the first and second waves would indicate a larger contribution of school contacts to transmission, however, a part of this contribution is likely due not to cases in schools but surrounding environments and relatives. It is also likely that the contribution of contacts in school to overall population transmission decreased in the third wave compared to previous months in Germany, as hygiene and testing measures were established within schools during that time.

The estimations made here are limited by the model used attempting to capture social dynamics through an epidemic compartmental model. In particular, we scaled the contact matrix in a certain way that likely has an impact on the contribution of the age groups. The model is also limited by not including the vaccinated compartment. The predictive ability of the model, however, has been analyzed by contributing to a forecast to the European COVID-19 Forecast Hub [39], which, in comparison to other models in Germany, shows above-average performance (calendar week 14 to 38 of 2021, our model had a relative WIS 0.61 and 0.52 for weekly forecasts of cases and death). Another limitation is that, particularly for the third wave, we had to make assumptions regarding the underdetection in children, that are not – contrary to the other underreporting estimates – based on seroprevalence but rather on comparisons of reported cases during times with and times without large scale testing in schools.

Despite these limitations, we believed that the estimation presented here adds to the understanding of how the actual contribution of different age groups during the pandemic unfolded in Germany and could enhance both scenario and predictive modeling efforts.

## Conclusions

We showed that taking underreporting into account, younger adults and teenagers are main contributors to infections during the first three pandemic waves in Germany. Overall, the contribution of contacts in schools to the total cases in the population was below 10% during the third wave. Accounting for age-specific underreporting seems important to correctly identify those parts of the population where quarantine, testing, vaccination and contact-reduction measures are likely to be most effective and efficient.

## Data Availability

All data produced in the present work are contained in the manuscript.

## Acknowledgments

We sincerely thank the scientific team behind the MuSPAD study, Daniela Gornyk, Stephan Glöckner, Monika Strengert, Tobias Kerrines, Gerhard Bajora, Stefanie Castell, Kerstin Frank, Knut Gubbe, Jana-Kristin Heise, Pilar Hernandez, Oliver Kappert, Winfried, Kern, Thomas Illig, Norman Klopp, Henrike Maaß, Julia Ortmann, Barbora Kessel, Gottfried Roller, Monika Schüter, Torsen Tonn, Michael Ziemons, Yvonne Kemmling, for supporting the underdetection of reported data.

## Authors’ contributions

Conceptualization: IR and BL; Methodology: IR; Software: IR; Validation: PV and AK; Formal Analysis: IR; Investigation: IR; Resources: IR; Data Curation: IR, PV, and MH; Writing – Original Draft: IR, PV, and BL; Writing – Review & Editing: NN; Visualization: IR; Supervision: BL and WB; Project Administration: IR; Funding Acquisition: BL and PV

