## Supplementary for "Age-specific Contribution of Contacts to Transmission of SARS-CoV-2 in Germany": Supplementary_CovidModeling.pdf

### Supplementary A. Data

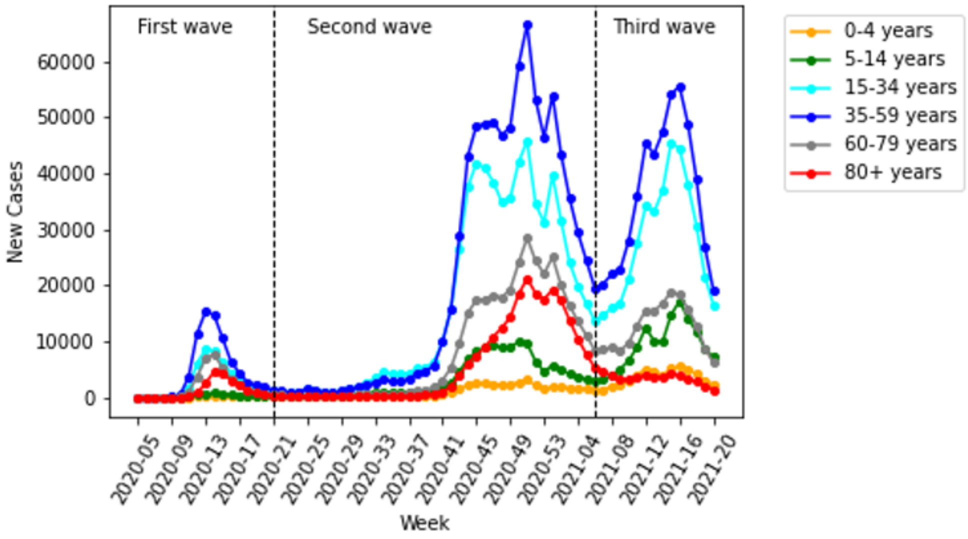


*Figure 1. The weekly reported cases for different age groups (Source: RKI [42])*

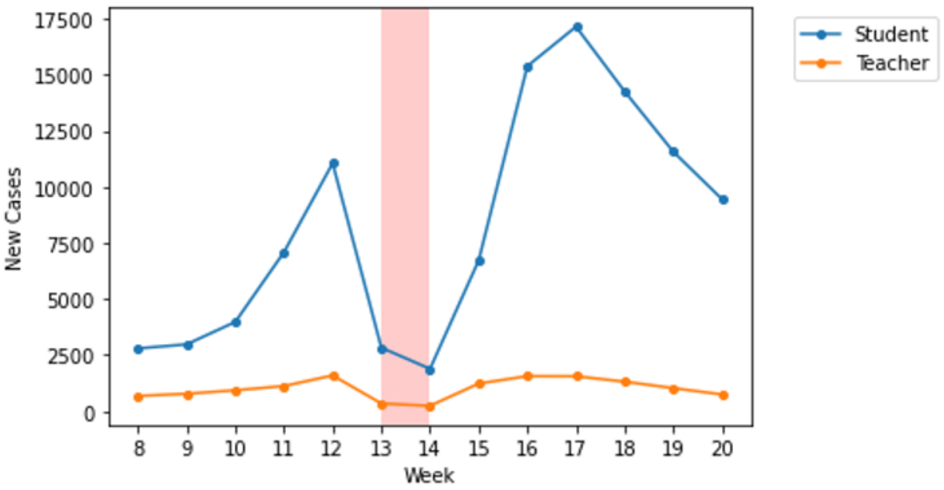


*Figure 2. The weekly reported cases for student and teacher in the third wave of 2021 (Source: KMK [43])*

*Table 1. The proportion students in 2019 for different age groups (Source: BMBF [39])*

| 5 – 9 years | 10 – 14 years | 15 – 19 years | 20 – 24 years |
|---|---|---|---|
| 20.47% | 28.15% | 26.51% | 17.31% |

*Table 2. The proportion of teaching staff 2019/2020 for different age groups (Source: [41])*

| <30 years | 30 – 39 years | 40 – 49 years | 50 – 59 years | >60 years |
|---|---|---|---|---|
| 6.52% | 28.06% | 26.14% | 26.74% | 12.26% |

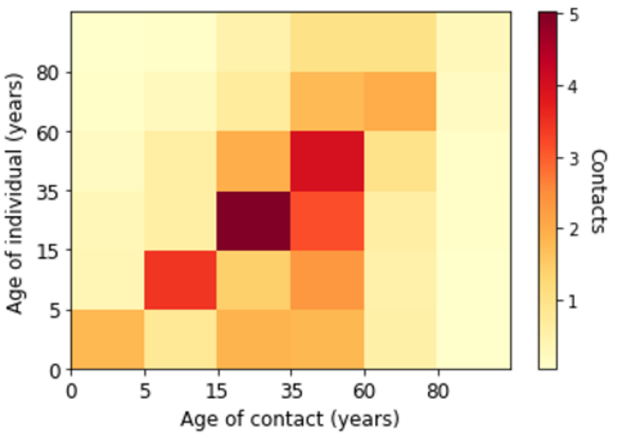

*Figure 3. Contact matrix in Germany for different age groups according to POLYMOD (Sources: [26])*

### Supplementary B. Parameters of Model

*Table 3. Parameterization by literature reviews (Source: RKI)*

| Parameter | Description |
|---|---|
| $P_1$ | The inverse of the incubation period (5 days) |
| $P_2$ | The inverse of the time a symptomatic patient recovers or requires hospitalization (4 days) |
| $P_3$ | The inverse of the time an asymptomatic patient recovers or dies (9 days) |
| $P_4$ | The inverse of the time a patient spends at a hospital before discharge (7 days) |
| $P_5$ | The inverse of the time span spent in ICU (10 days) |
| $P_6$ | The inverse of the time of a patient with long-term complication discharges (14 days) |
| $P_7$ | The inverse of the time span of a recovered individual with long-term complication before reinfection (90 days) |
| $P_8$ | The inverse of the time span of a fully recovered individual before reinfection (360 days) |
| $\beta_s$ | The transmission from a symptomatic infected |
| $\beta_a$ | The transmission from an asymptomatic infected |
| $\kappa$ | The fraction of symptomatic infected |
| $\alpha$ | The proportion of the symptomatic cases requiring hospitalization |
| $\delta$ | The percentage of hospitalized patients requiring ICU |
| $\vartheta$ | The proportion of the patients in ICU who will die |
| $\nu$ | The proportion of the asymptomatic who will die |
| $\rho$ | The proportion of the symptomatic cases having a long-term complication |
| $\varphi$ | The proportion of the hospitalized patients having a long-term complication |
| $\sigma$ | The proportion of the patients in ICU having a long-term complication |

| | |
|---|---|
| $\eta$ | The proportion of the patients with a long-term complication requiring hospitalization |
| $\gamma$ | The proportion of the patients with a long-term complication who will die |

*Table 4. Parametrization by age group (Source: [29])*

| Group | Symptomatic cases requiring hospitalization | Hospitalized cases requiring ICU | Symptomatic cases having a long-term complication |
|---|---|---|---|
| 0 – 4 years | 0.01% | 5.0% | 0.01% |
| 5 – 14 years | 0.12% | 5.0% | 0.1% |
| 15 – 34 years | 3.2% | 6.3% | 10% |
| 35 – 59 years | 4.9% | 12.2% | 20% |
| 60 – 79 years | 15.2% | 30.3% | 40% |
| 80+ years | 27.3% | 70.9% | 50% |

### Supplementary C. Estimated Marginal Force of Infection in Contacts

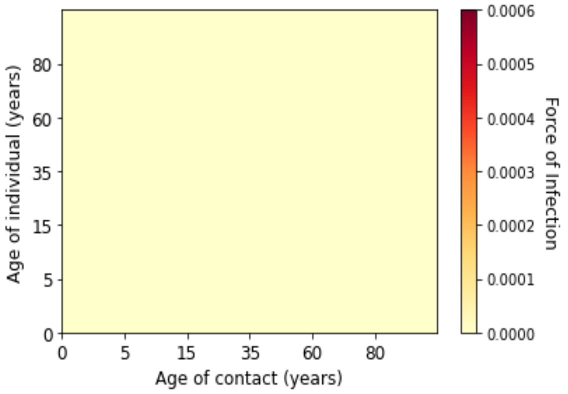


*(a) Week 6 of 2020*

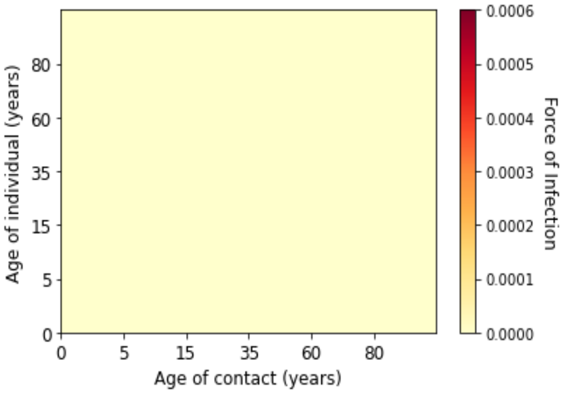


*(b) Week 7 of 2020*

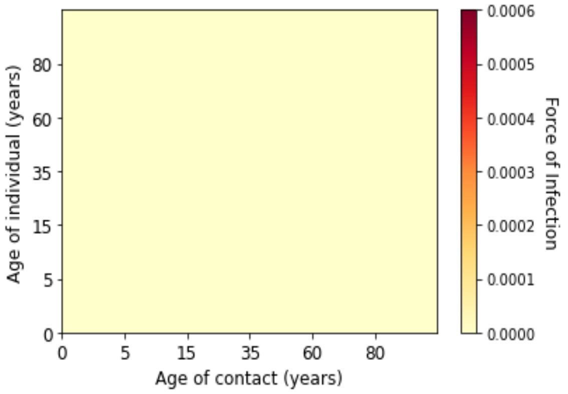


*(c) Week 8 of 2020*

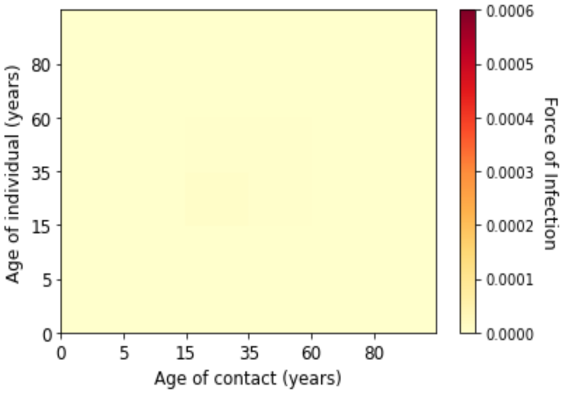


*(d) Week 9 of 2020*

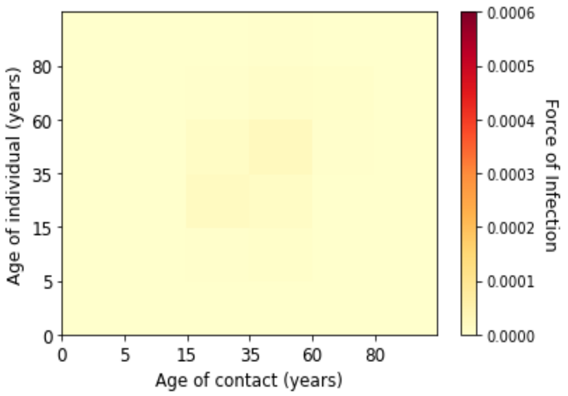


*(e) Week 10 of 2020*

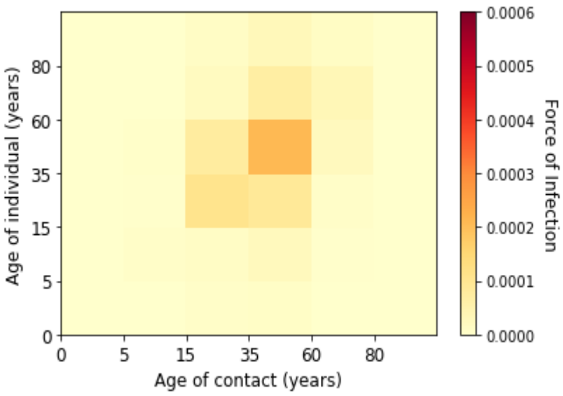


*(f) Week 11 of 2020*

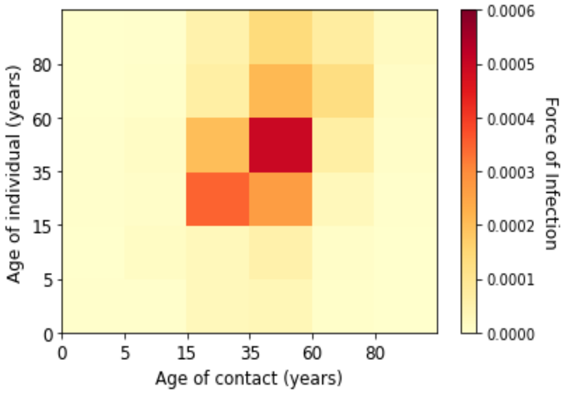


*(g) Week 12 of 2020*

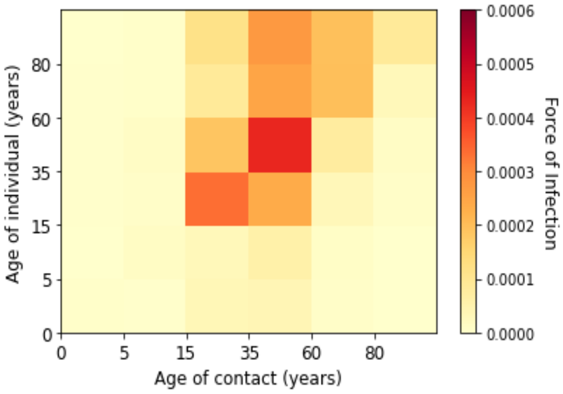


*(h) Week 13 of 2020*

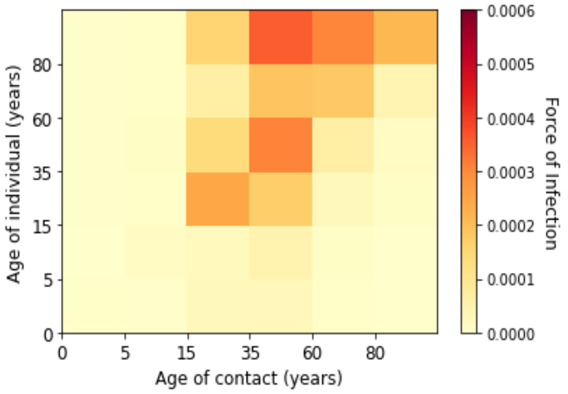


*(i) Week 14 of 2020*

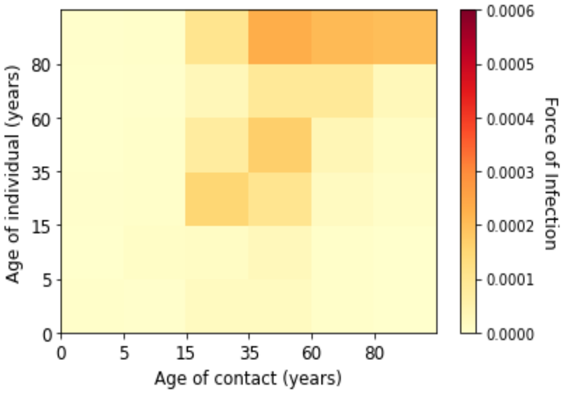


*(j) Week 15 of 2020*

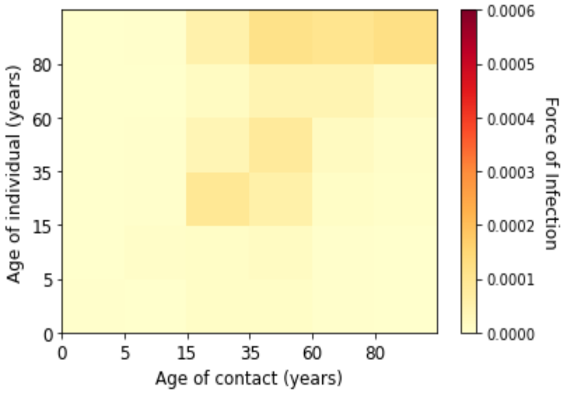


*(k) Week 16 of 2020*

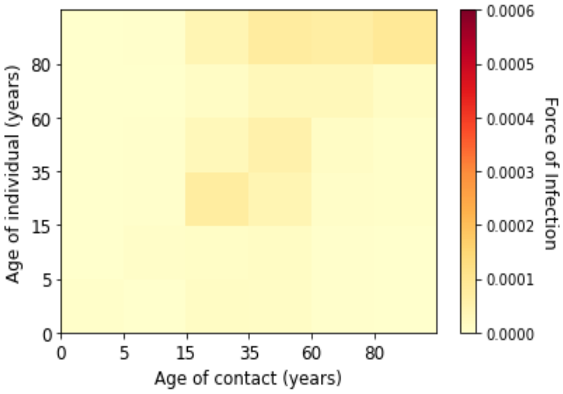


*(l) Week 17 of 2020*

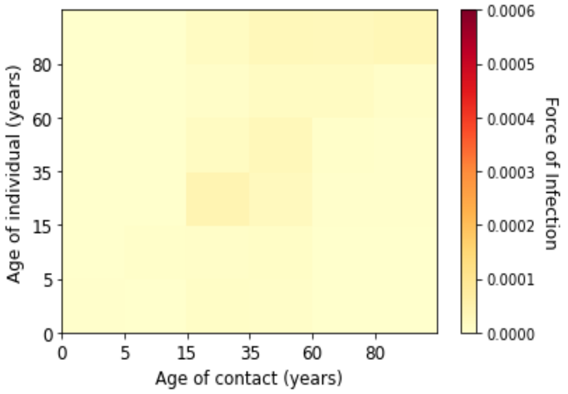


*(m) Week 18 of 2020*

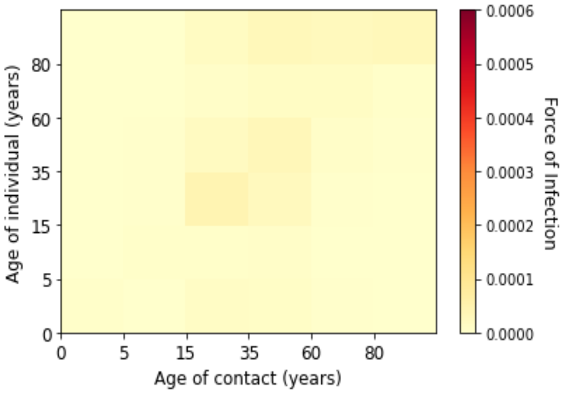


*(n) Week 19 of 2020*

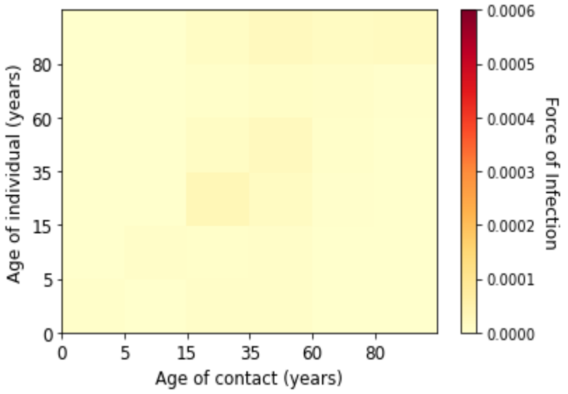


*(o) Week 20 of 2020*

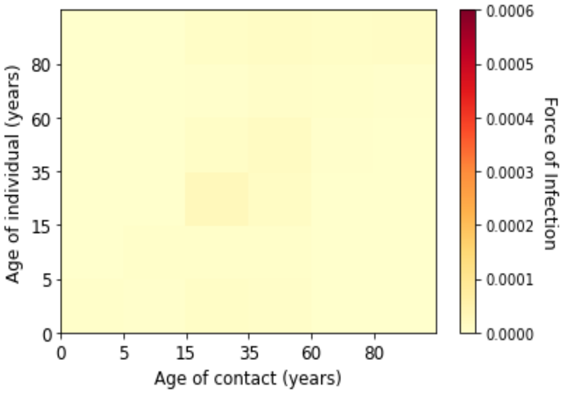


*(p) Week 21 of 2020*

*Figure 4. Estimated marginal force of infection for each age groups in the first wave*

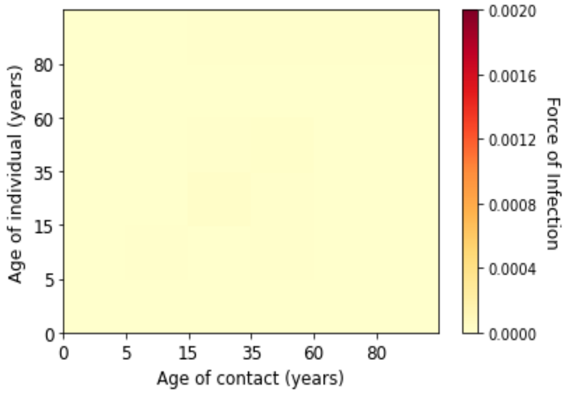


*(a) Week 22 of 2020*

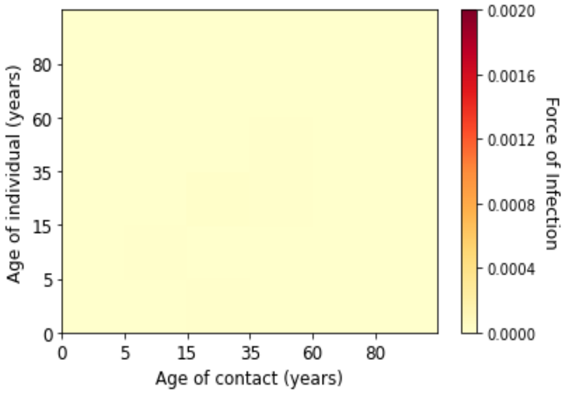


*(b) Week 23 of 2020*

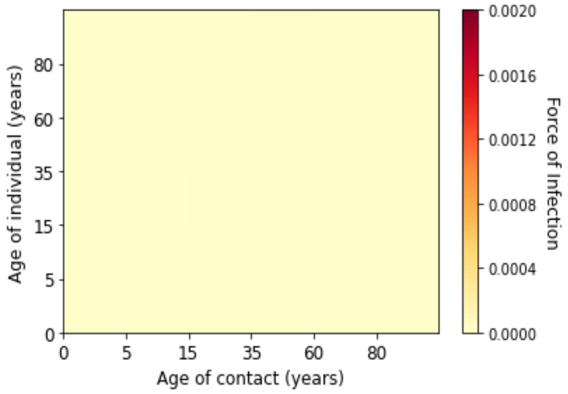


*(c) Week 24 of 2020*

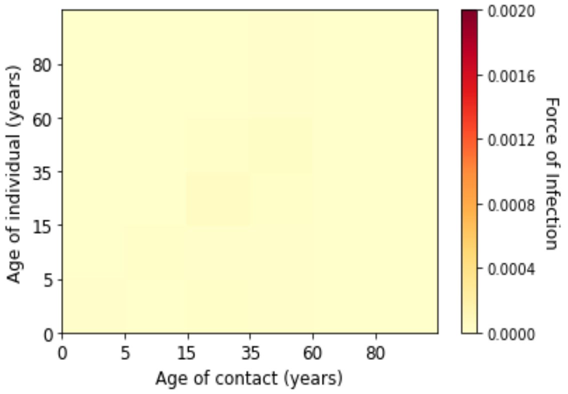


*(d) Week 25 of 2020*

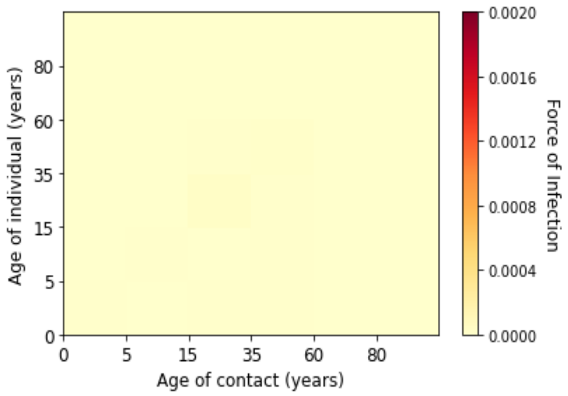


(e) Week 26 of 2020

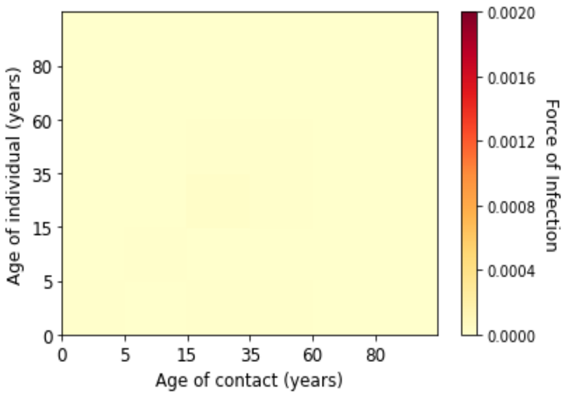


(f) Week 27 of 2020

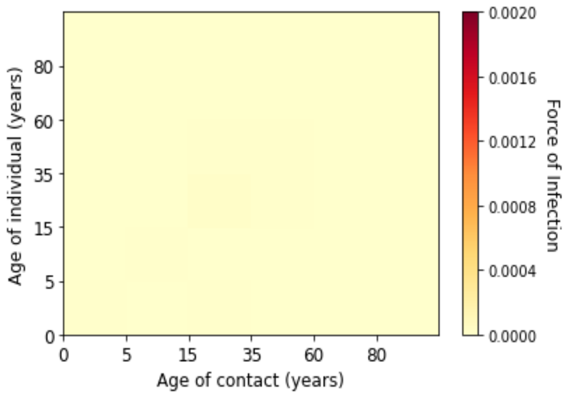


(g) Week 28 of 2020

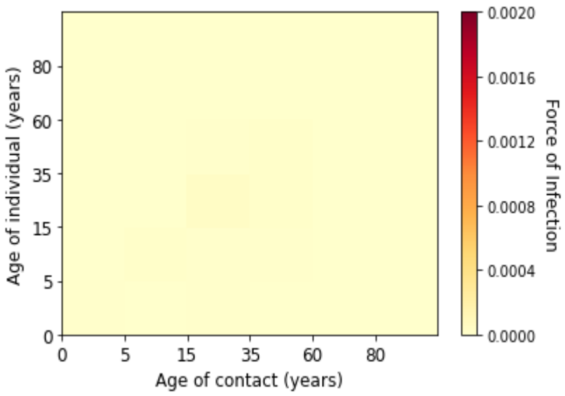


(h) Week 29 of 2020

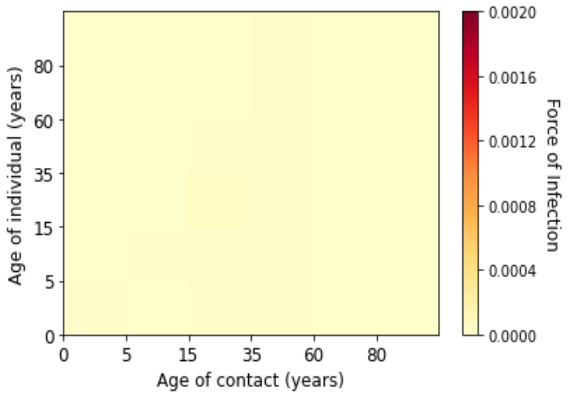


(i) Week 30 of 2020

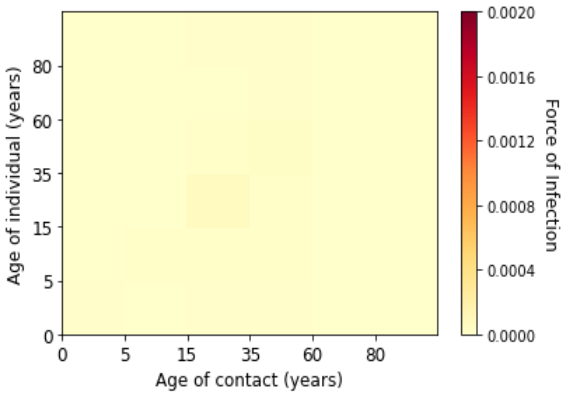


(j) Week 31 of 2020

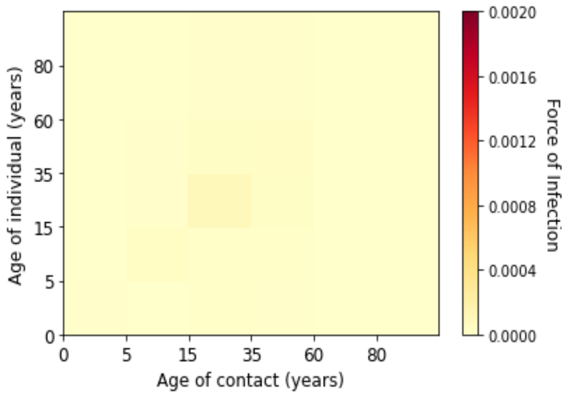


(k) Week 32 of 2020

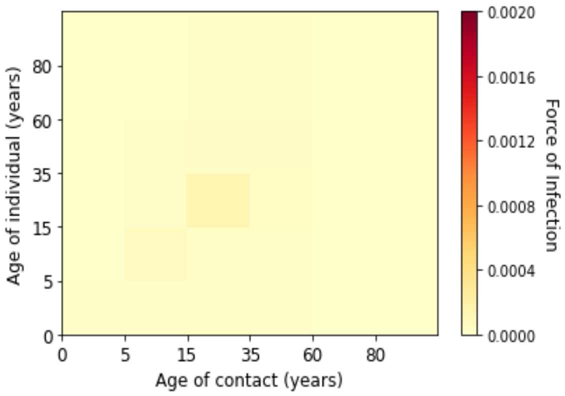


(l) Week 33 of 2020

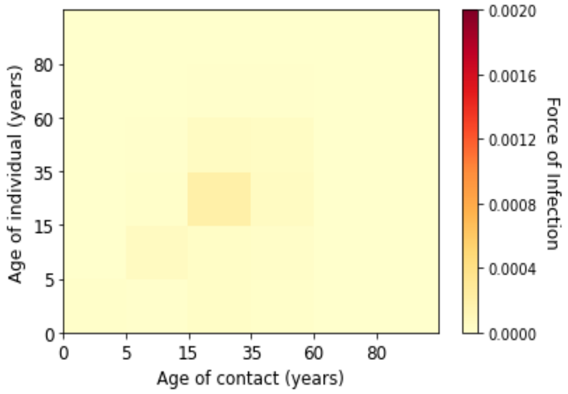

(m) Week 34 of 2020

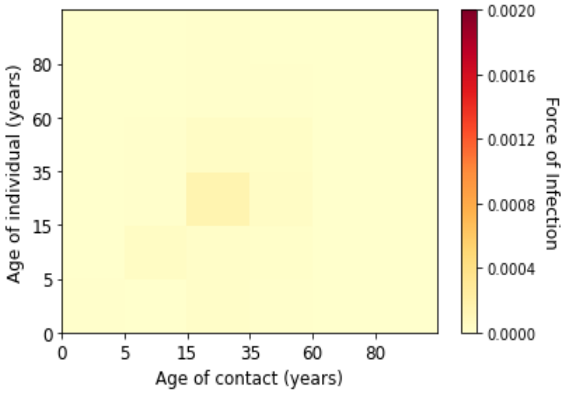

(n) Week 35 of 2020

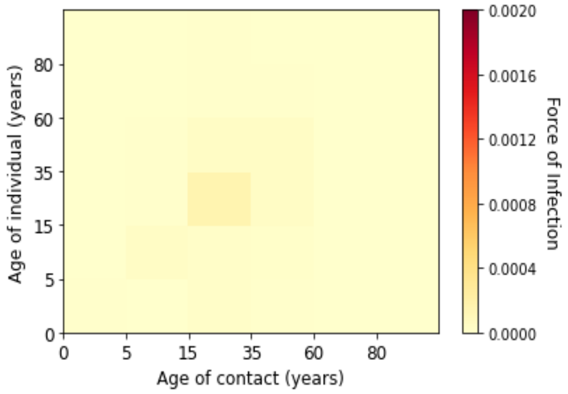

(o) Week 36 of 2020

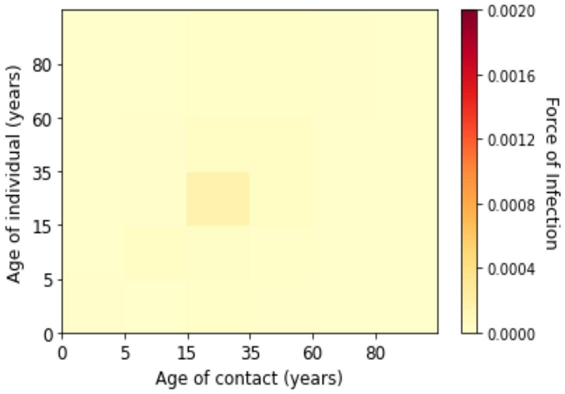

(p) Week 37 of 2020

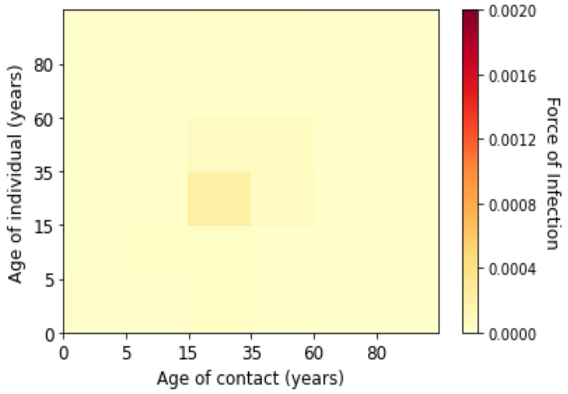

(q) Week 38 of 2020

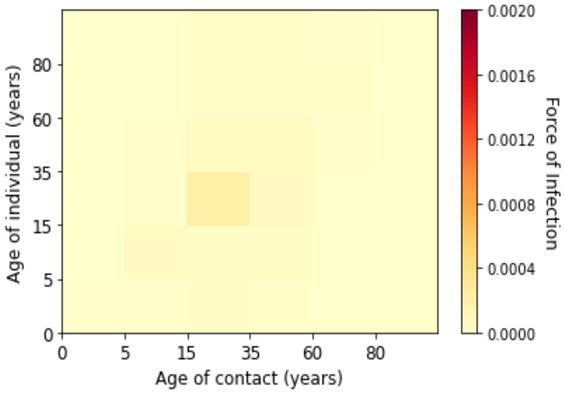

(r) Week 39 of 2020

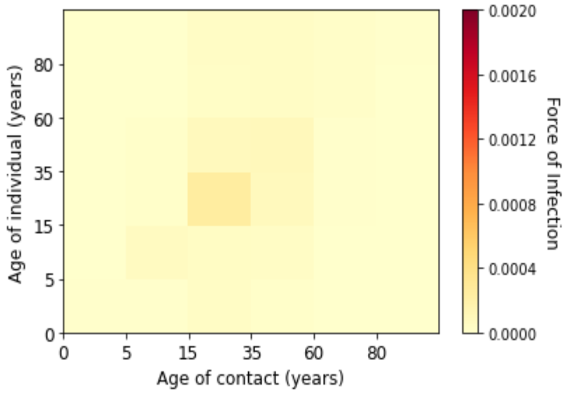

(s) Week 40 of 2020

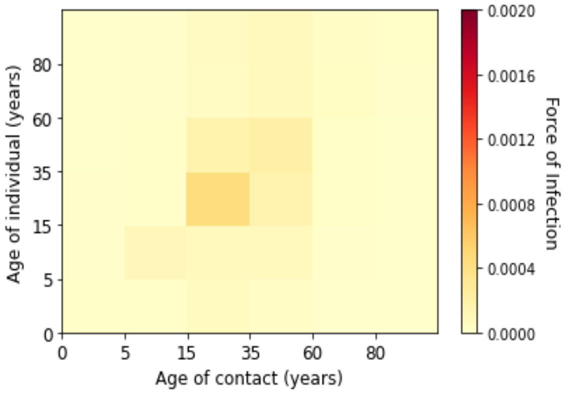

(t) Week 41 of 2020

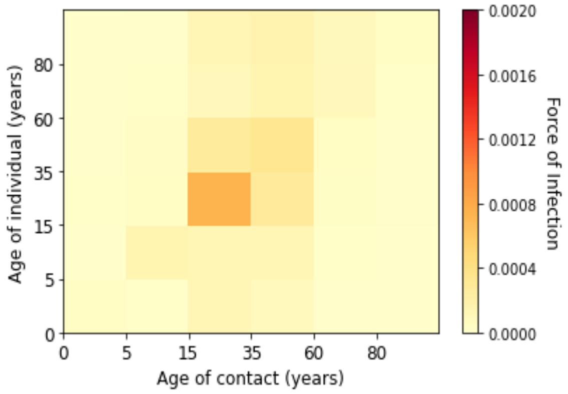


*(u) Week 42 of 2020*

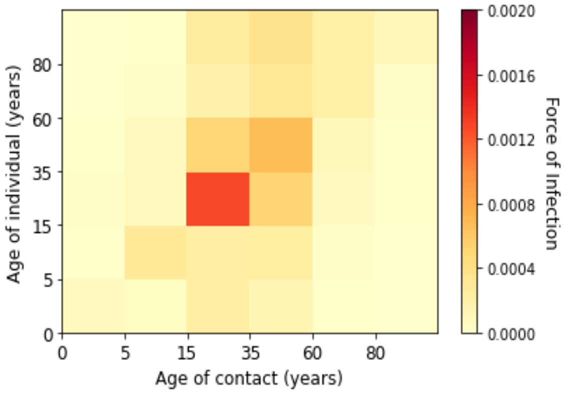


*(v) Week 43 of 2020*

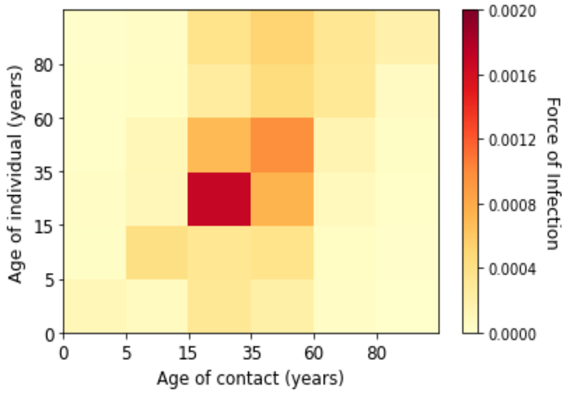


*(w) Week 44 of 2020*

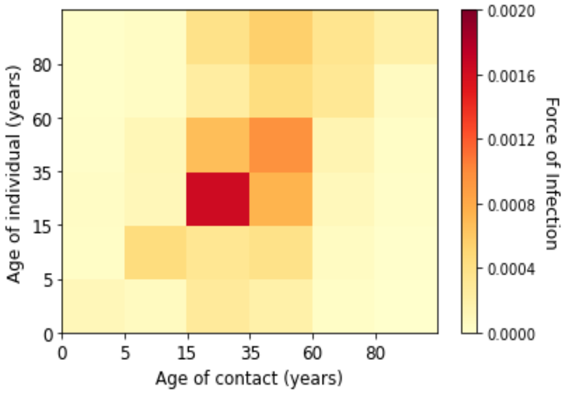


*(x) Week 45 of 2020*

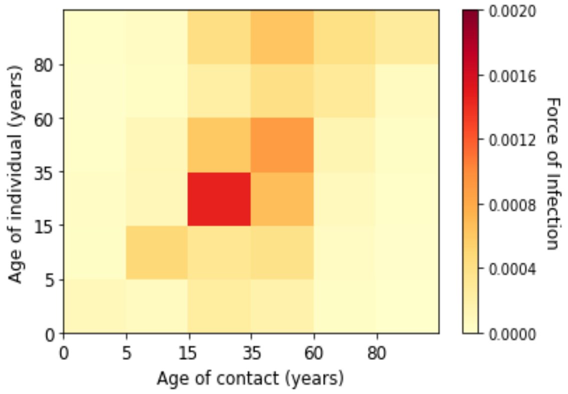


*(y) Week 46 of 2020*

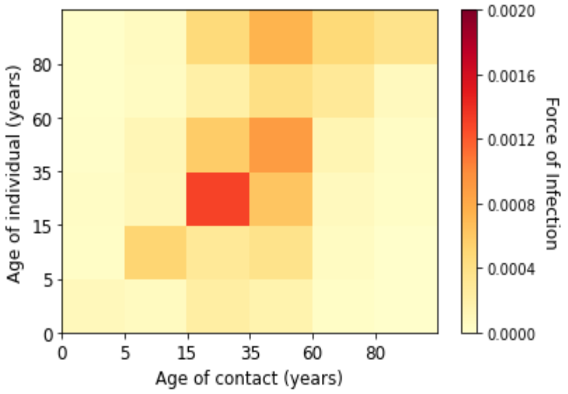


*(z) Week 47 of 2020*

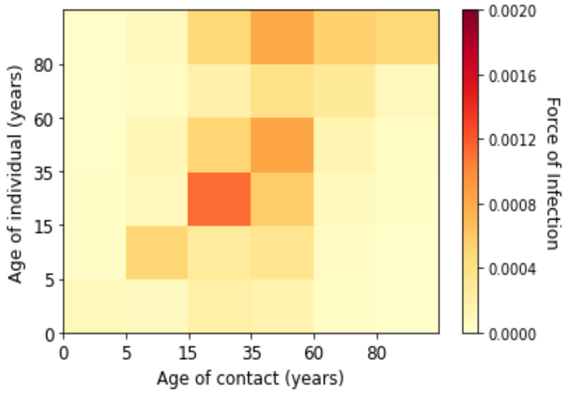


*(aa) Week 48 of 2020*

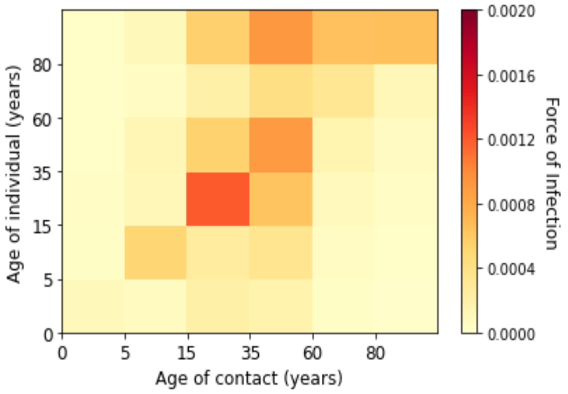


*(ab) Week 49 of 2020*

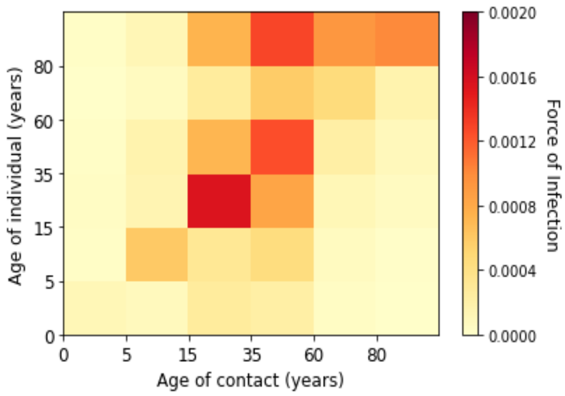


*(ac) Week 50 of 2020*

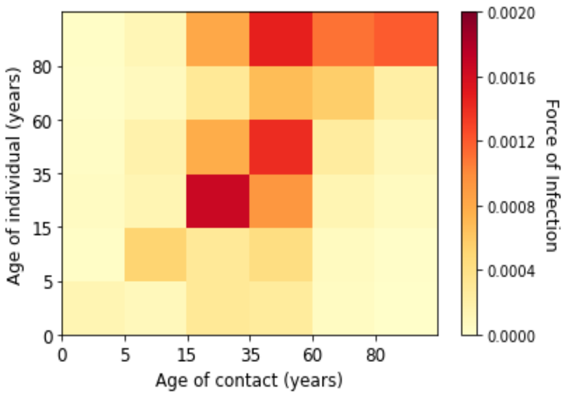


*(ad) Week 51 of 2020*

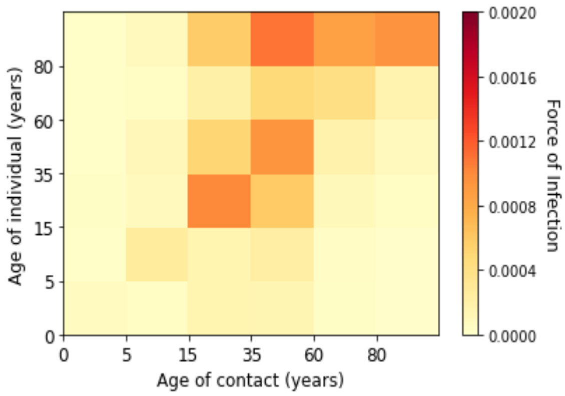


*(ae) Week 52 of 2020*

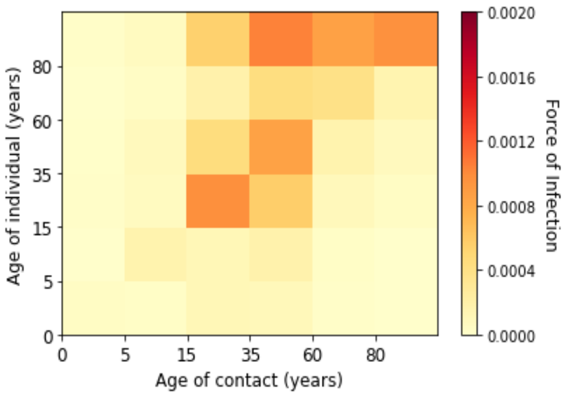


*(af) Week 53 of 2020*

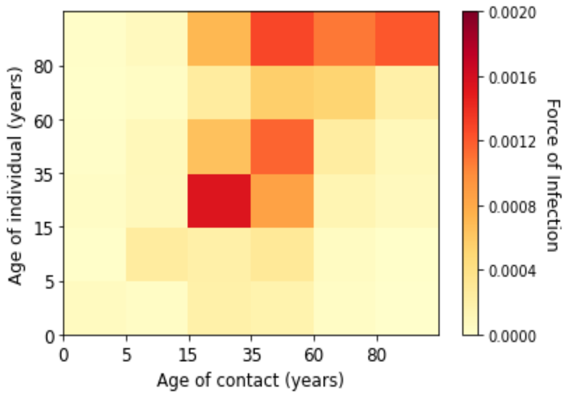


*(ag) Week 1 of 2021*

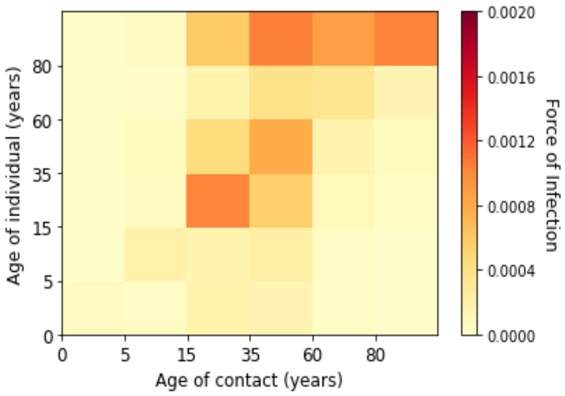


*(ah) Week 2 of 2021*

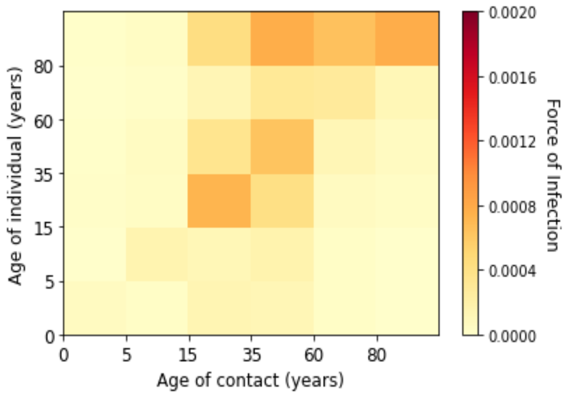


*(ai) Week 3 of 2021*

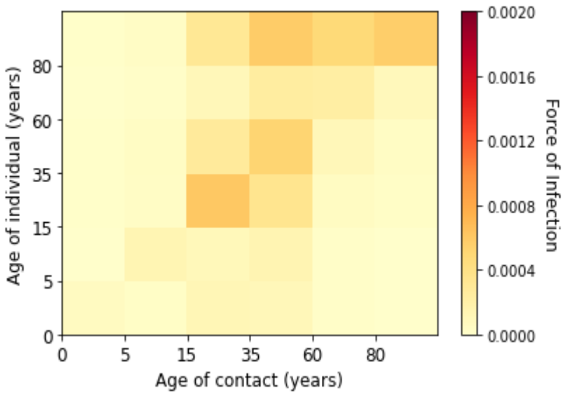


*(aj) Week 4 of 2021*

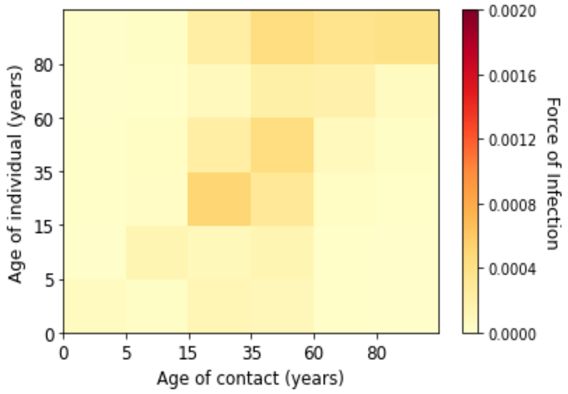


*(ak) Week 5 of 2021*

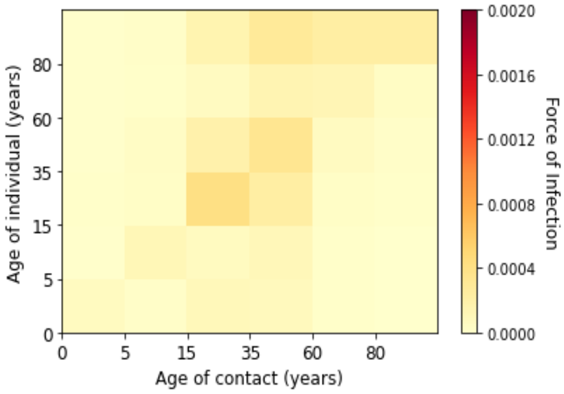


*(al) Week 6 of 2021*

*Figure 5. Estimated marginal force of infection for each age groups in the second wave*

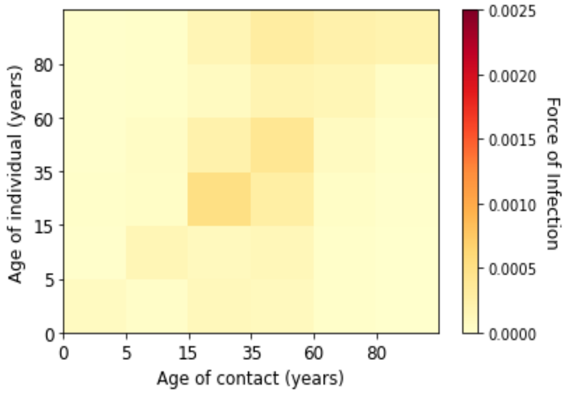


*(a) Week 7 of 2021*

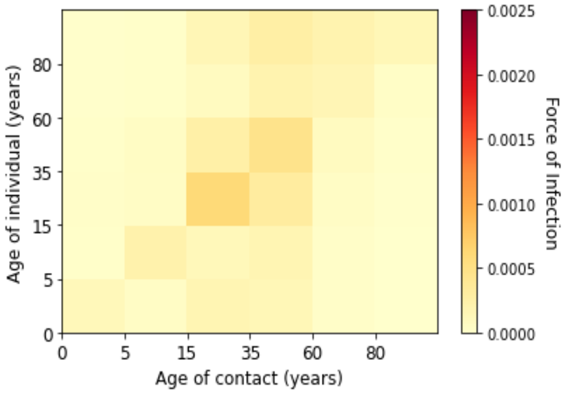


*(b) Week 8 of 2021*

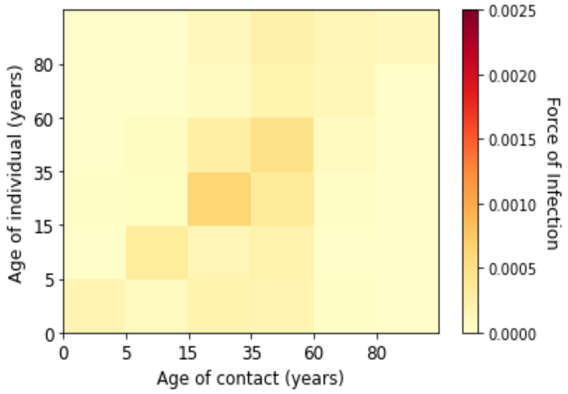


*(c) Week 9 of 2021*

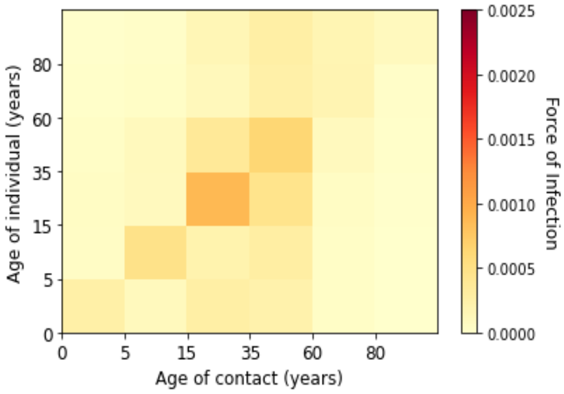


*(d) Week 10 of 2021*

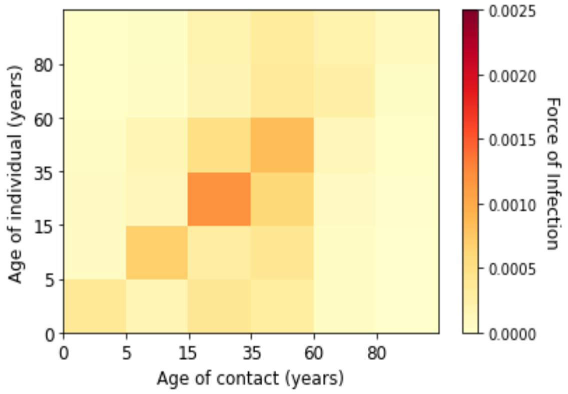


*(e) Week 11 of 2021*

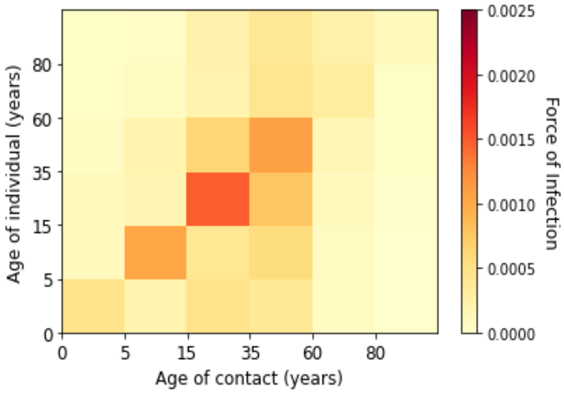


*(f) Week 12 of 2021*

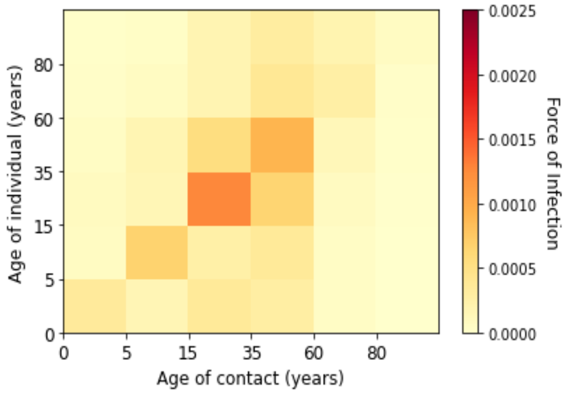


*(g) Week 13 of 2021*

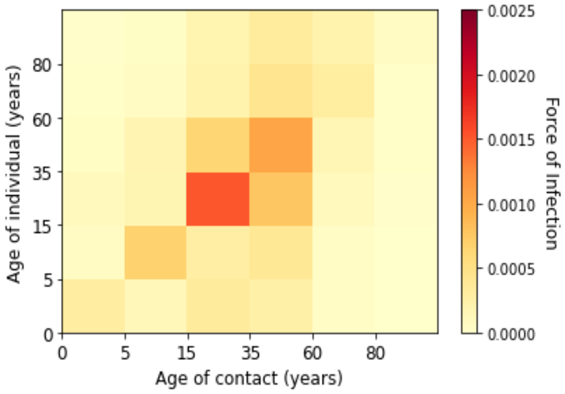


*(h) Week 14 of 2021*

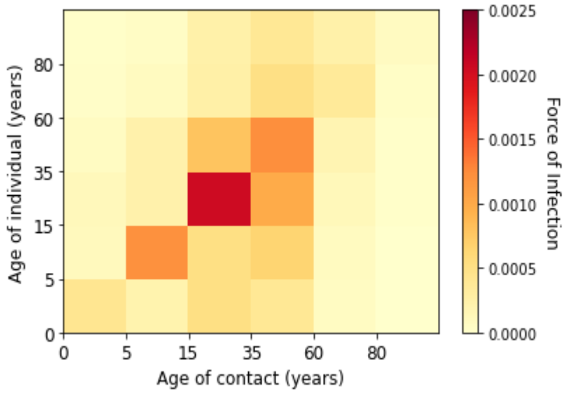


*(i) Week 15 of 2021*

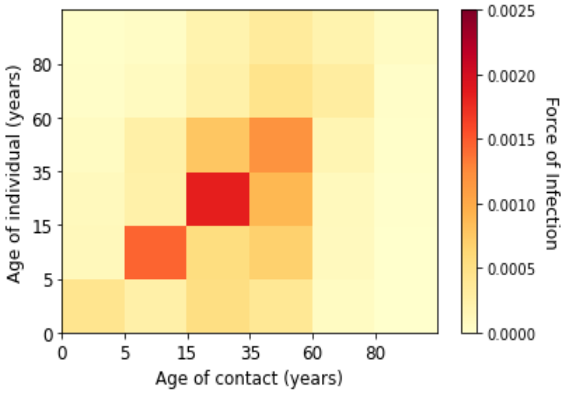


*(j) Week 16 of 2021*

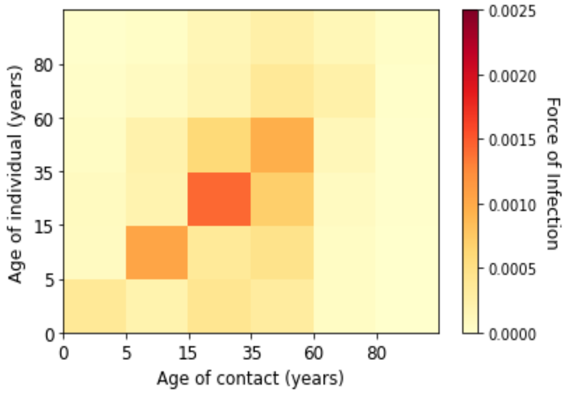


*(k) Week 17 of 2021*

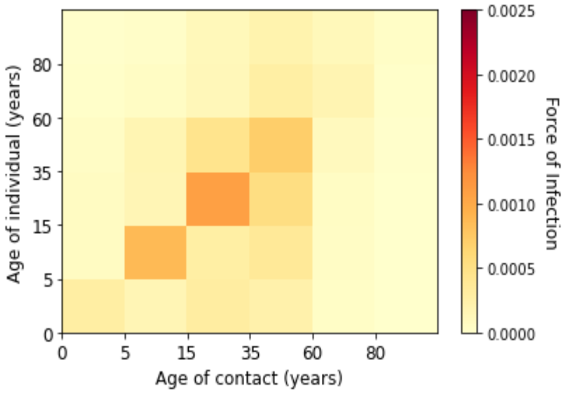


*(l) Week 18 of 2021*

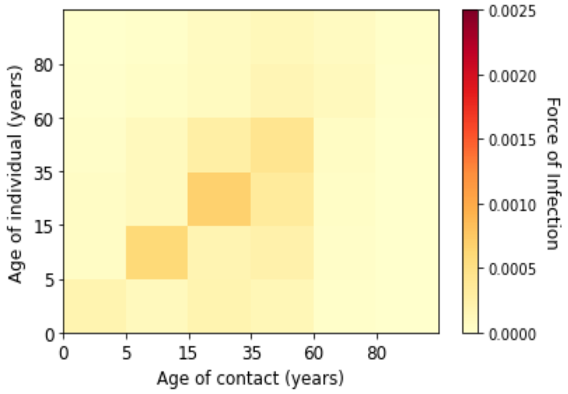


*(m) Week 19 of 2021*

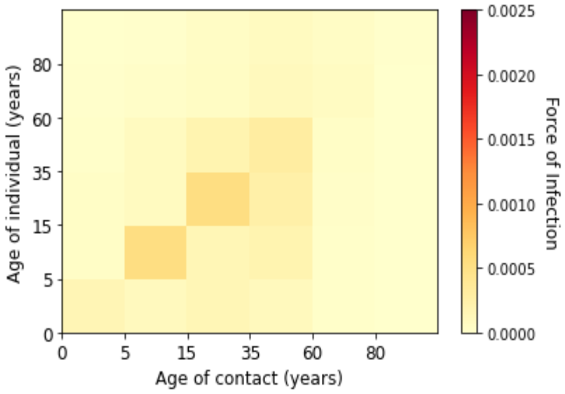


*(n) Week 20 of 2021*

*Figure 6. Estimated marginal force of infection for each age groups in the third wave*

### Supplementary D. Estimated Contribution of Contacts to Transmission

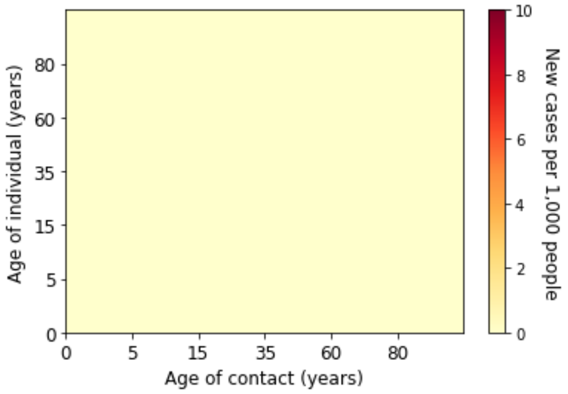


*(a) Week 6 of 2020*

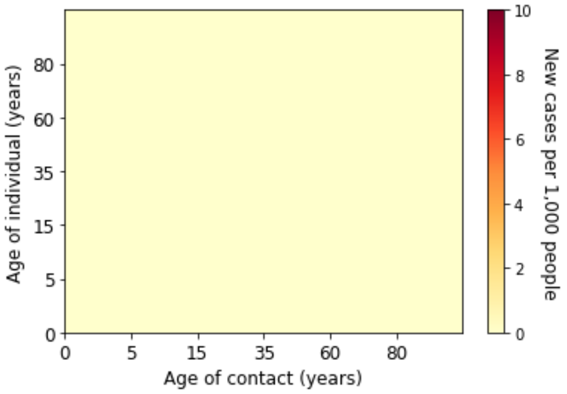


*(b) Week 7 of 2020*

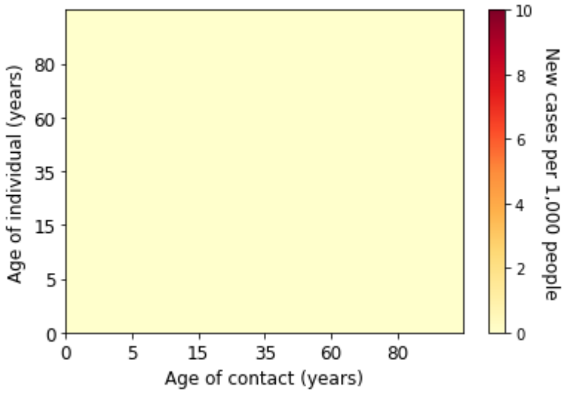


*(c) Week 8 of 2020*

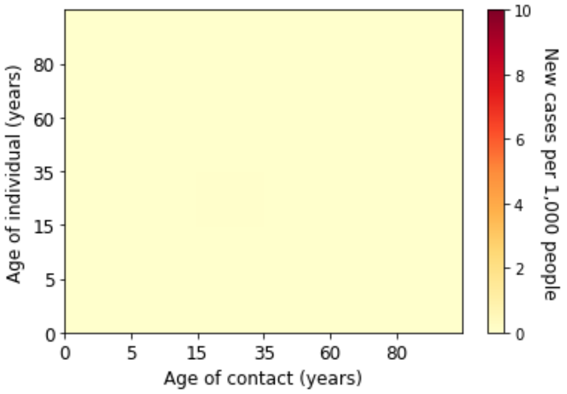


*(d) Week 9 of 2020*

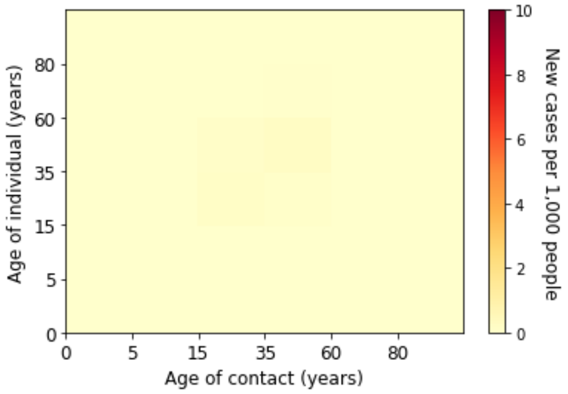


*(e) Week 10 of 2020*

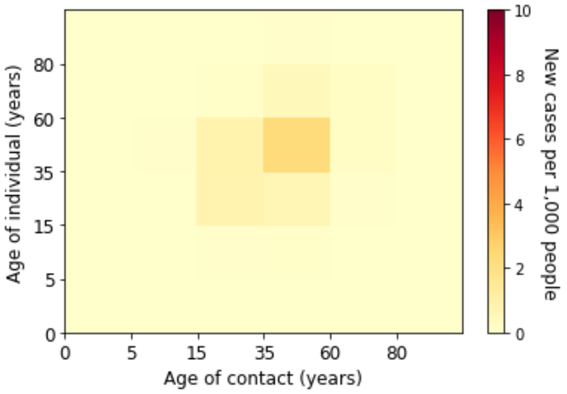


*(f) Week 11 of 2020*

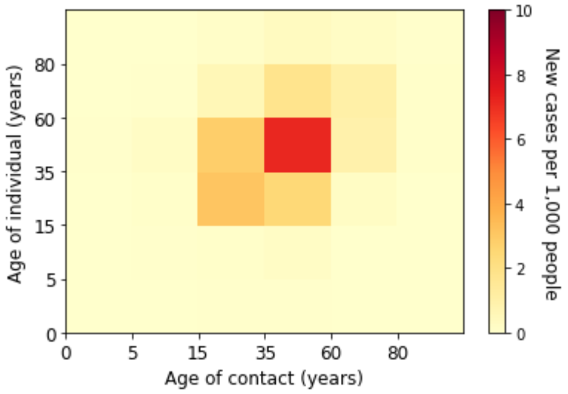


*(g) Week 12 of 2020*

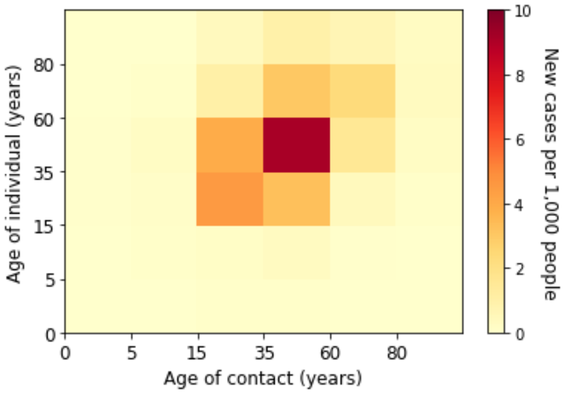


*(h) Week 13 of 2020*

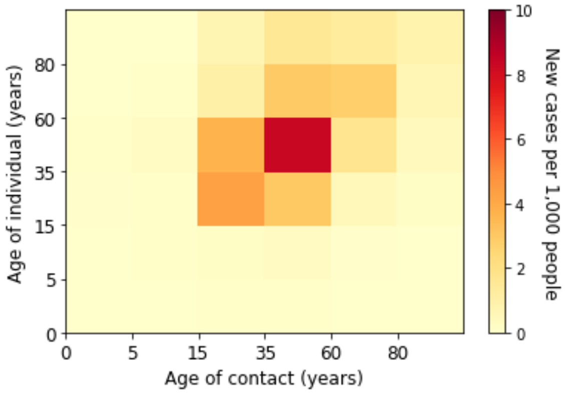


*(i) Week 14 of 2020*

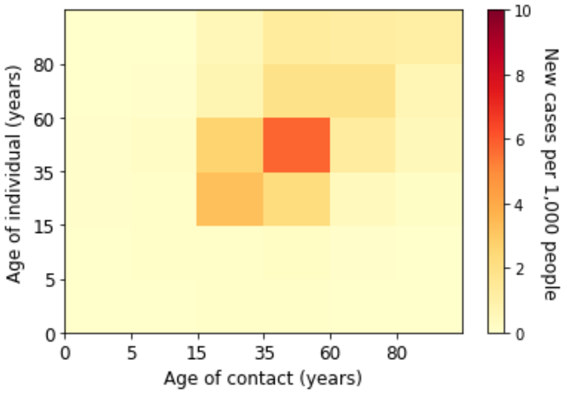


*(j) Week 15 of 2020*

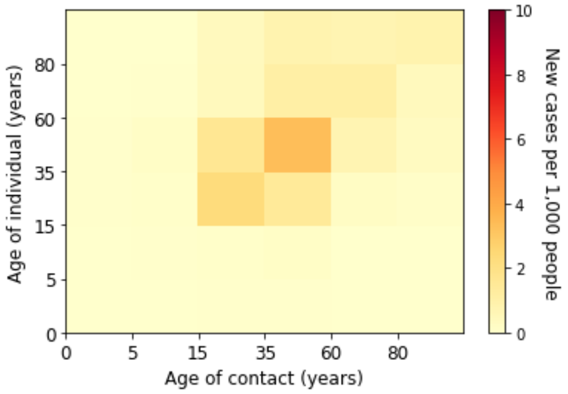


*(k) Week 16 of 2020*

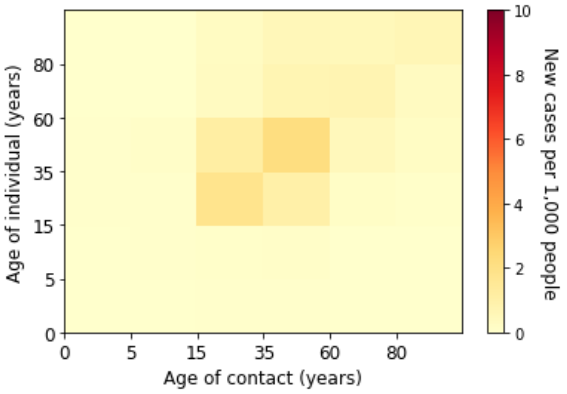


*(l) Week 17 of 2020*

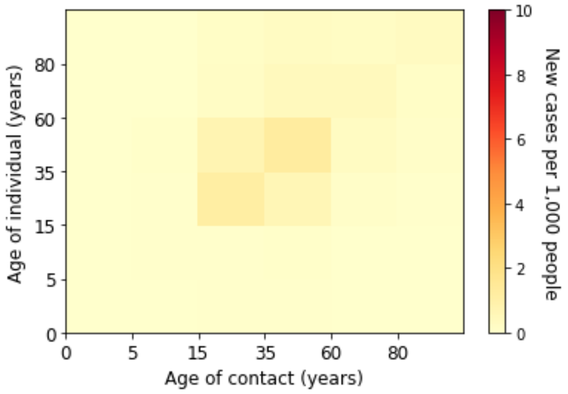


*(m) Week 18 of 2020*

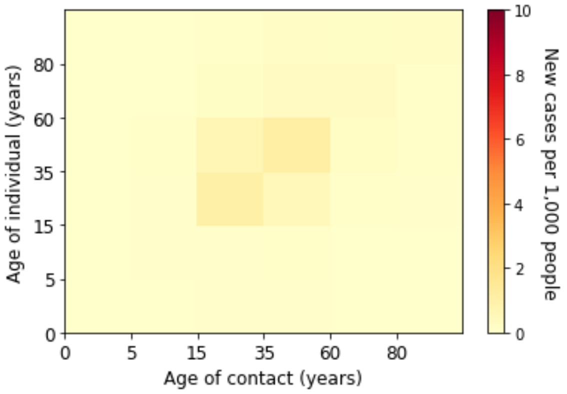


*(n) Week 19 of 2020*

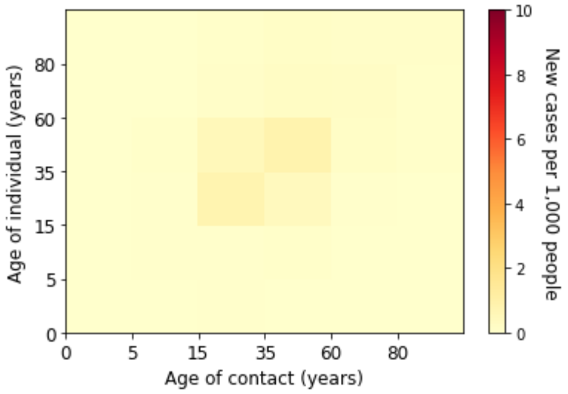


*(o) Week 20 of 2020*

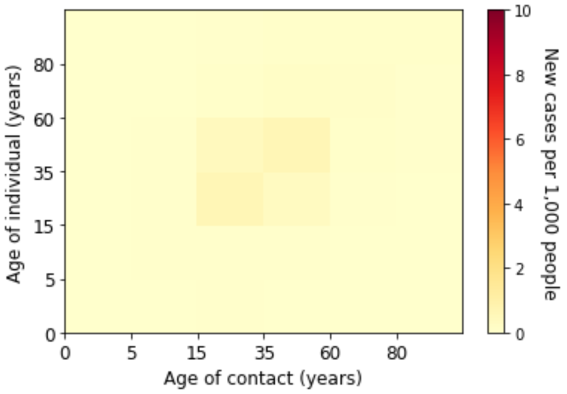


*(p) Week 21 of 2020*

*Figure 7. Estimated absolute contribution of transmission for each age groups in the first wave*

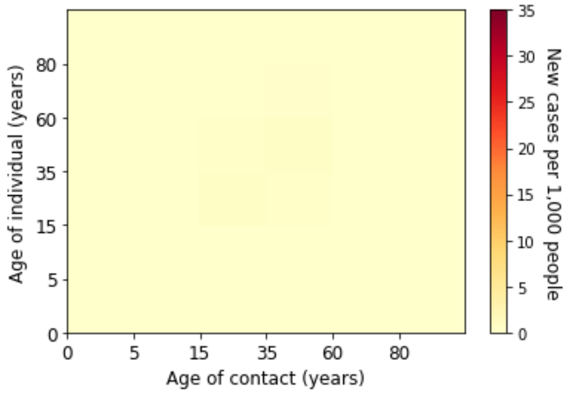


*(a) Week 22 of 2020*

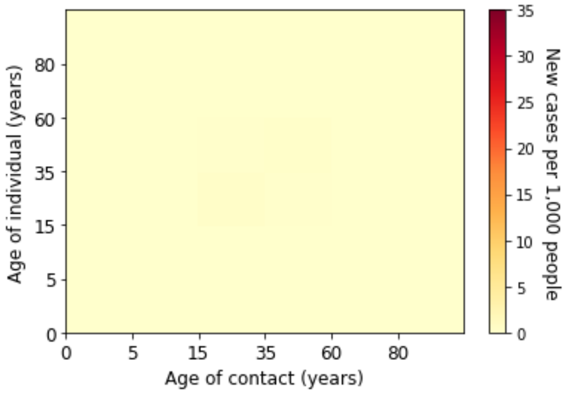


*(b) Week 23 of 2020*

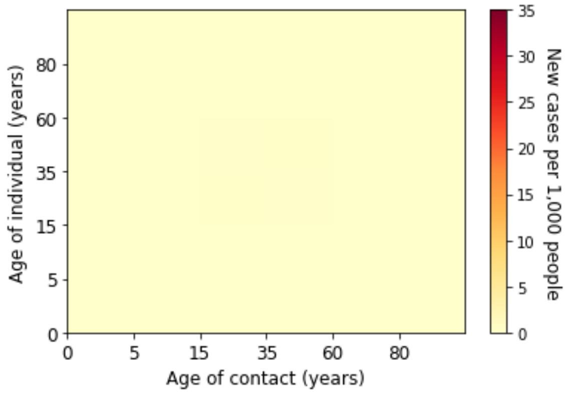


*(c) Week 24 of 2020*

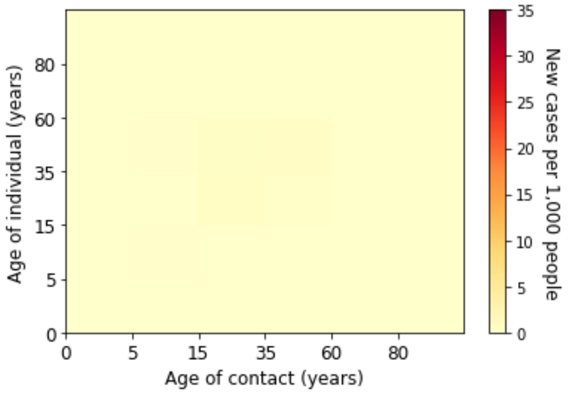


*(d) Week 25 of 2020*

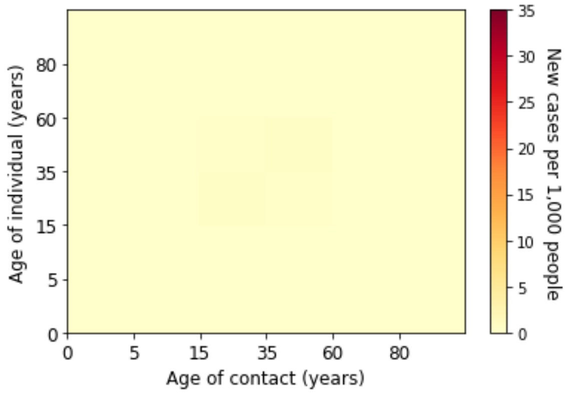


*(e) Week 26 of 2020*

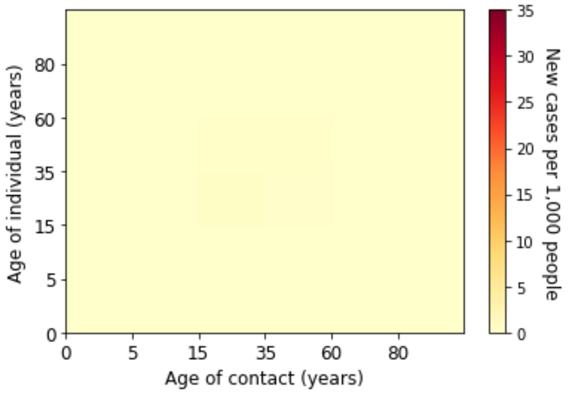


*(f) Week 27 of 2020*

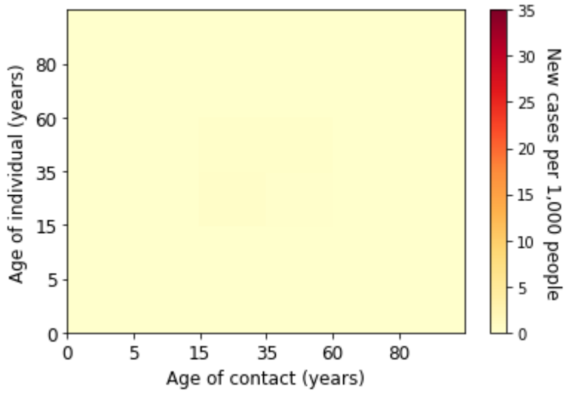


*(g) Week 28 of 2020*

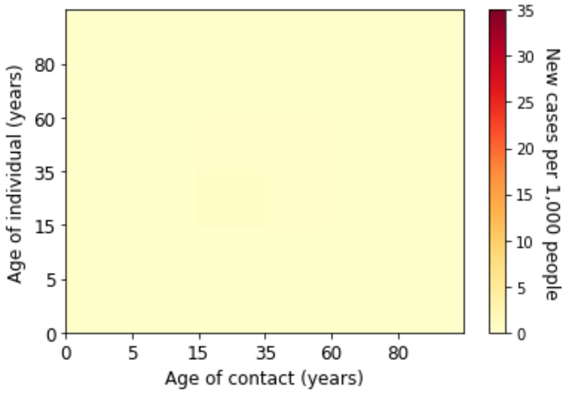


*(h) Week 29 of 2020*

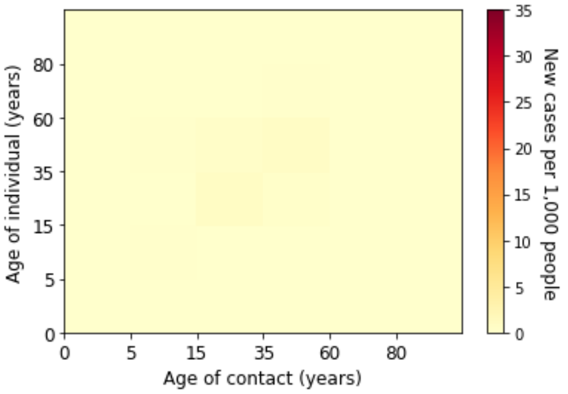


*(i) Week 30 of 2020*

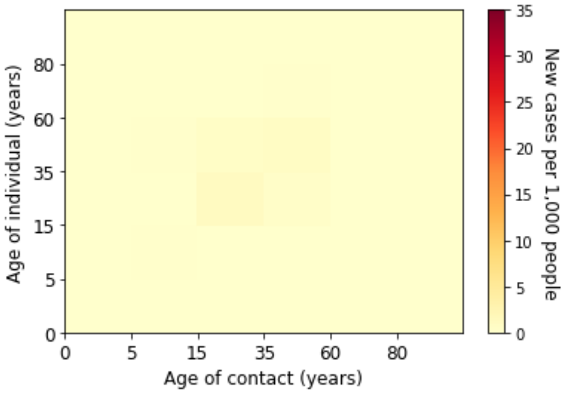


*(j) Week 31 of 2020*

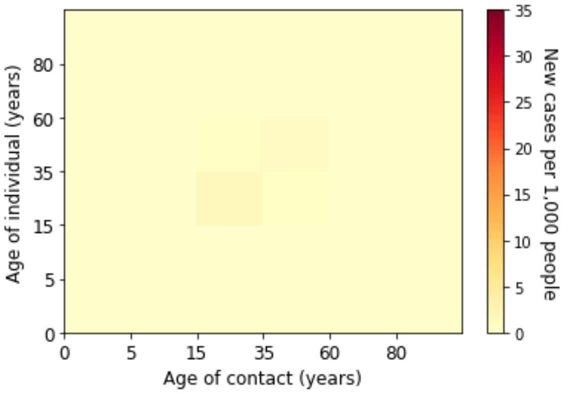


*(k) Week 32 of 2020*

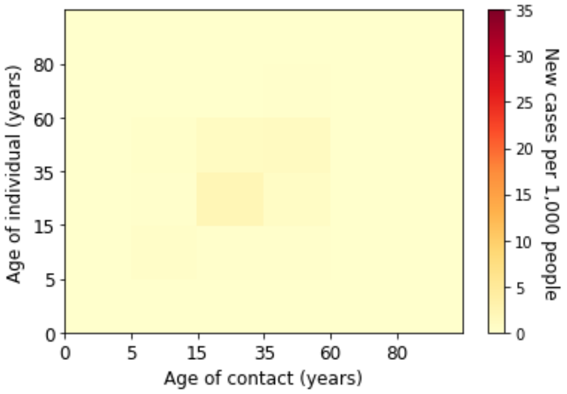


*(l) Week 33 of 2020*

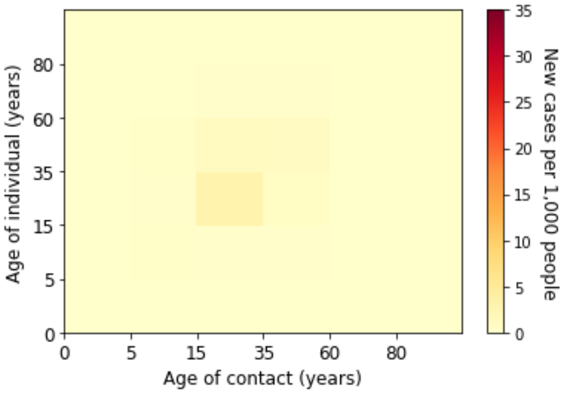


*(m) Week 34 of 2020*

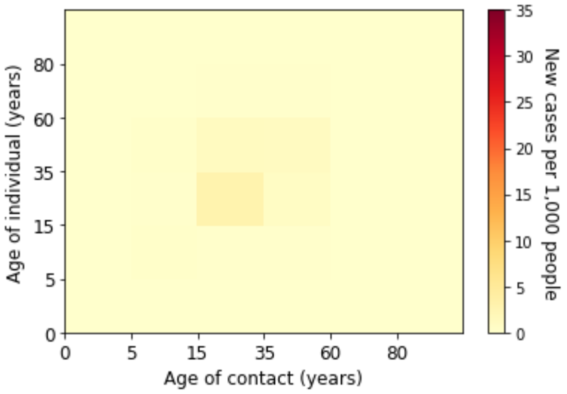


*(n) Week 35 of 2020*

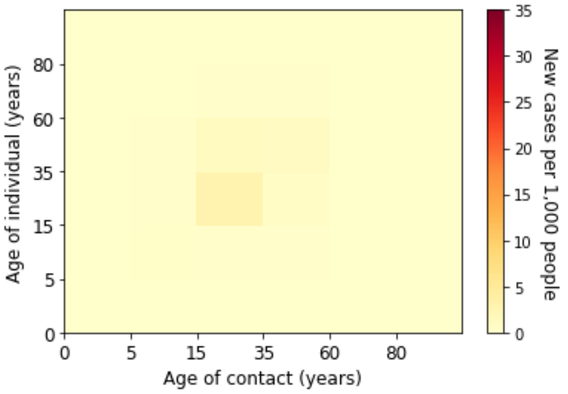


*(o) Week 36 of 2020*

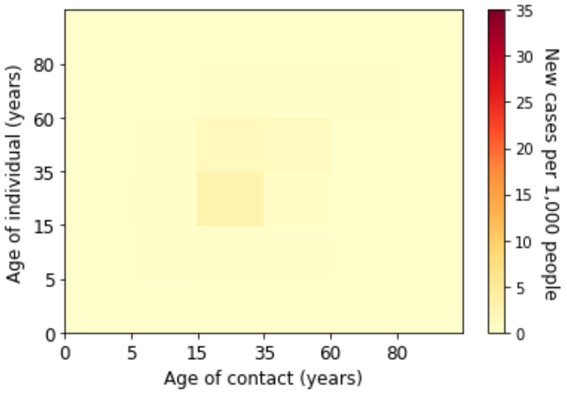


*(p) Week 37 of 2020*

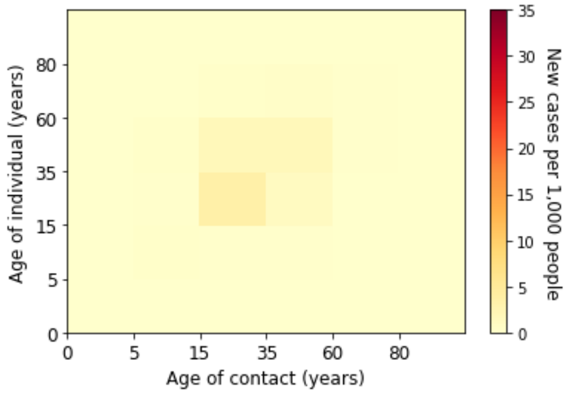


*(a) Week 38 of 2020*

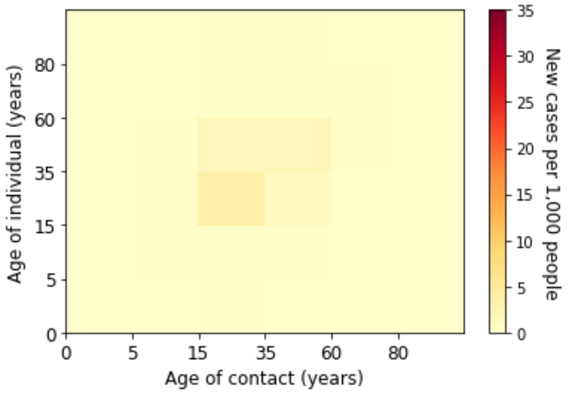


*(b) Week 39 of 2020*

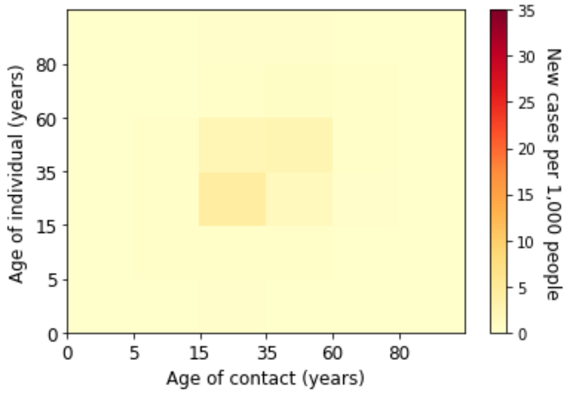


*(c) Week 40 of 2020*

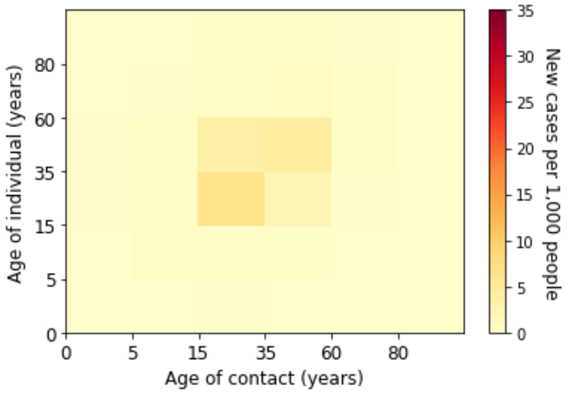


*(d) Week 41 of 2020*

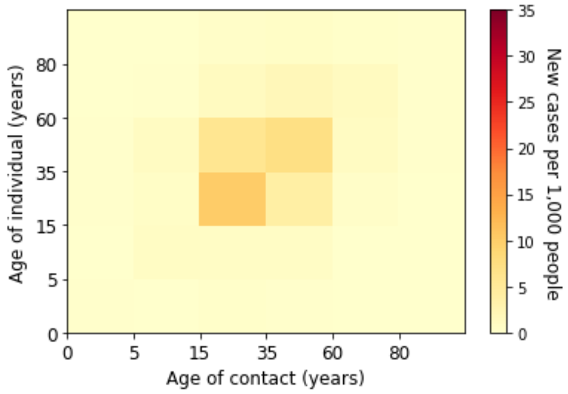


*(e) Week 42 of 2020*

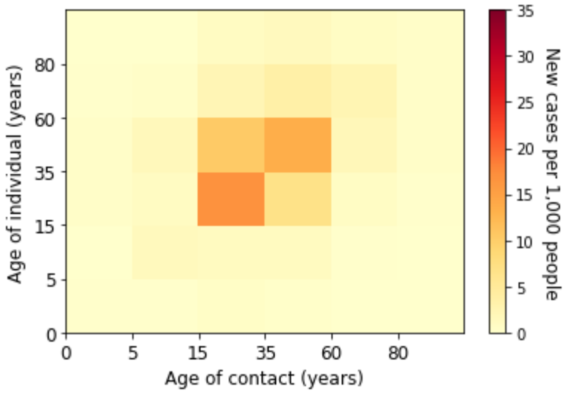


*(f) Week 43 of 2020*

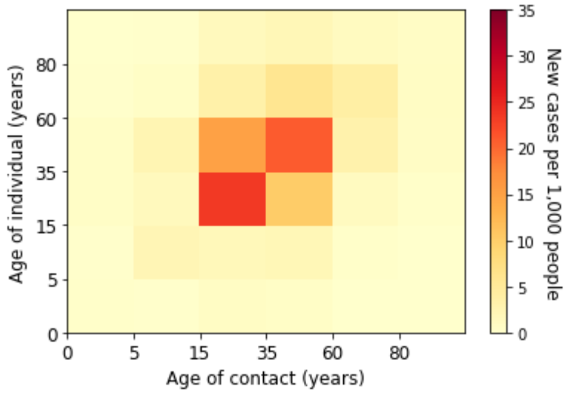


*(g) Week 44 of 2020*

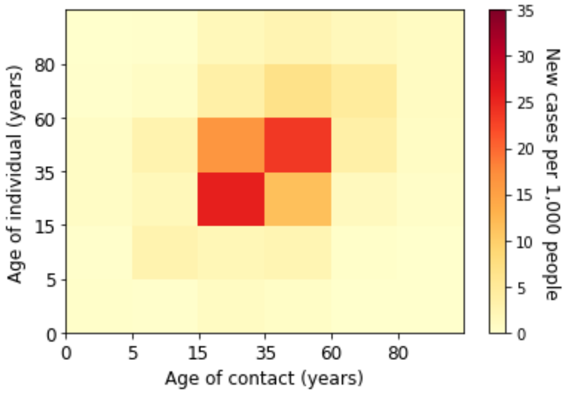


*(h) Week 45 of 2020*

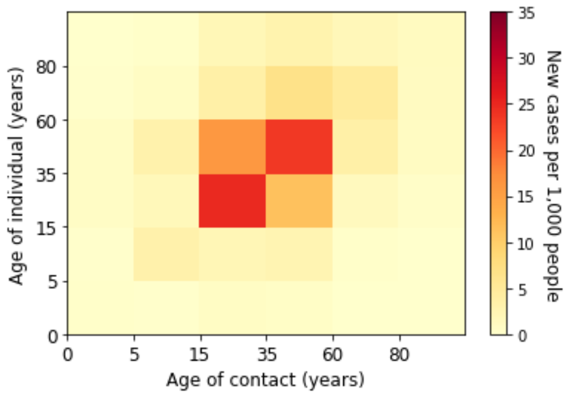


*(i) Week 46 of 2020*

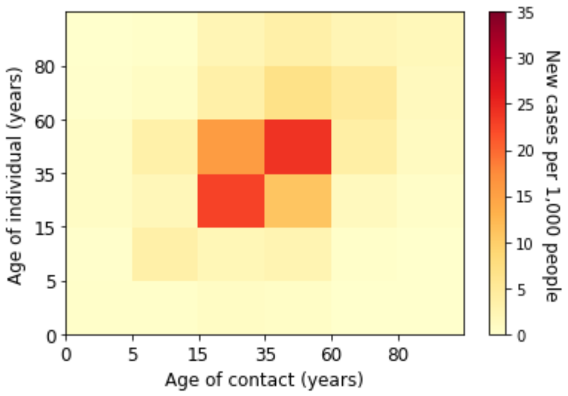


*(j) Week 47 of 2020*

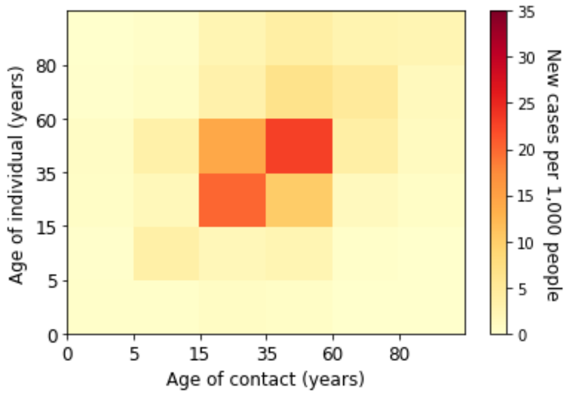


*(k) Week 48 of 2020*

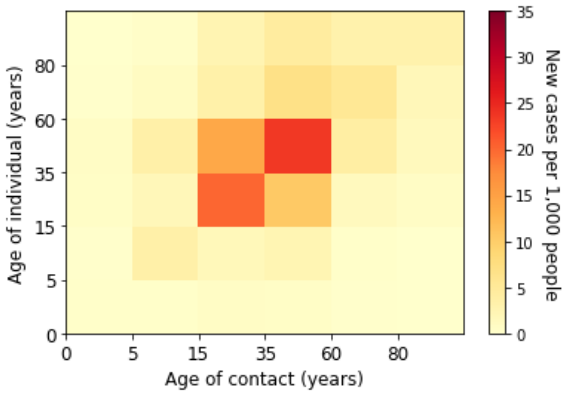


*(l) Week 49 of 2020*

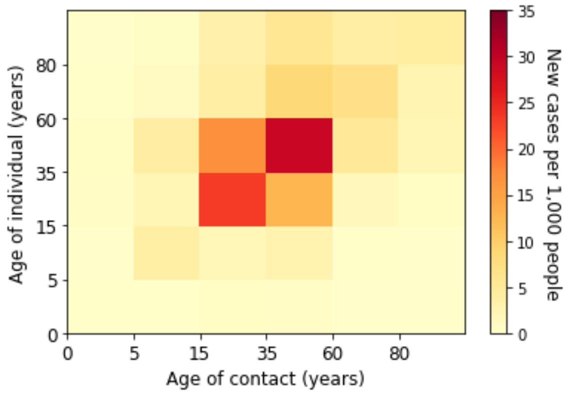


*(m) Week 50 of 2020*

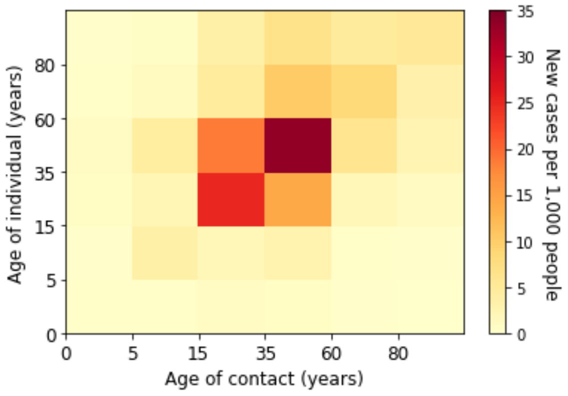


*(n) Week 51 of 2020*

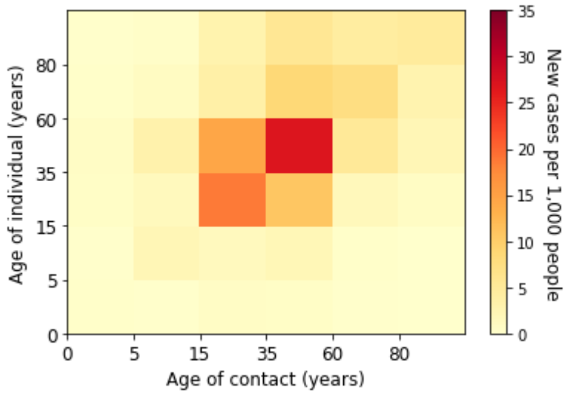


*(o) Week 52 of 2020*

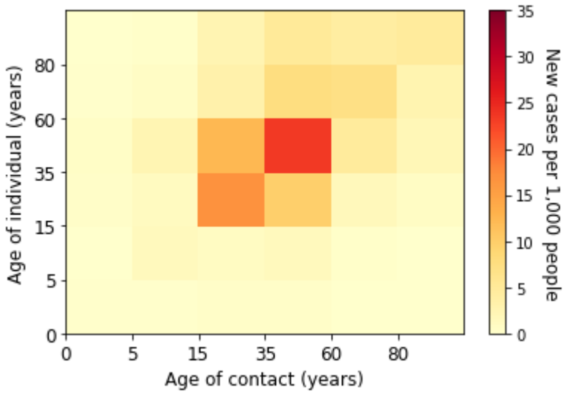


*(p) Week 53 of 2020*

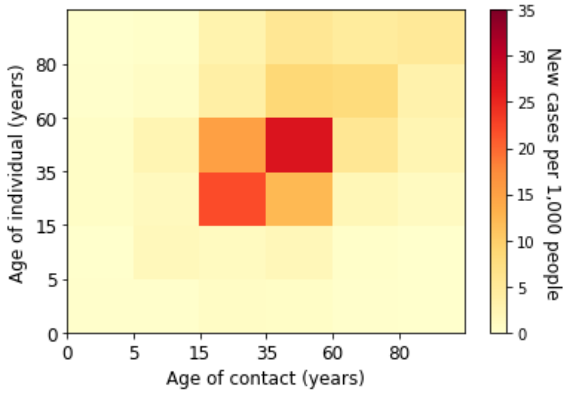


*(q) Week 1 of 2021*

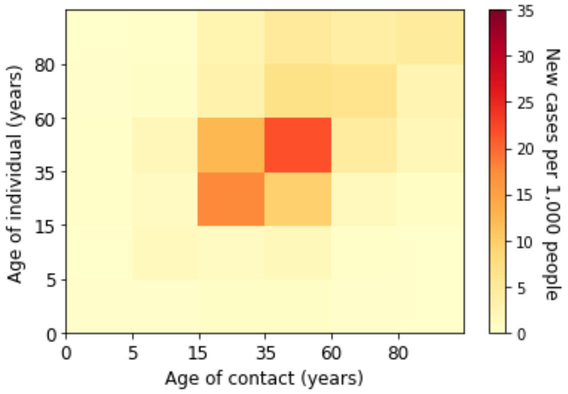


*(r) Week 2 of 2021*

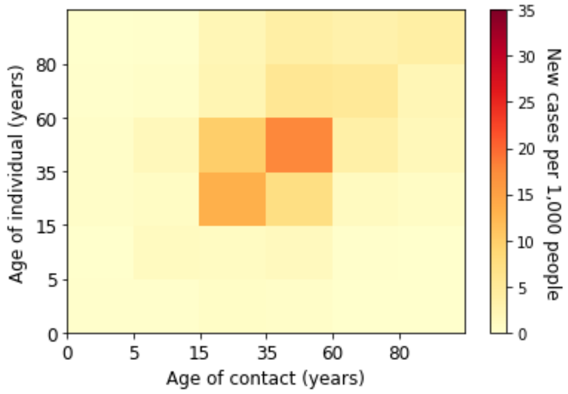


*(s) Week 3 of 2021*

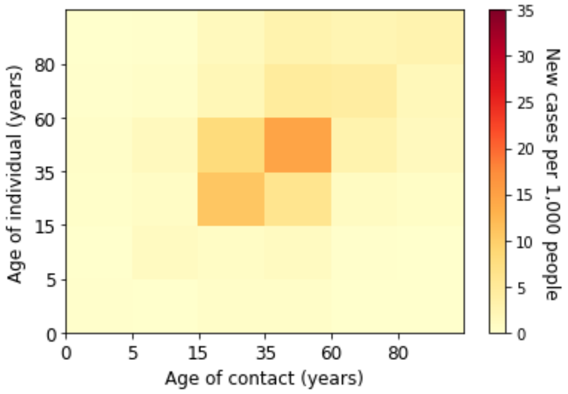


*(t) Week 4 of 2021*

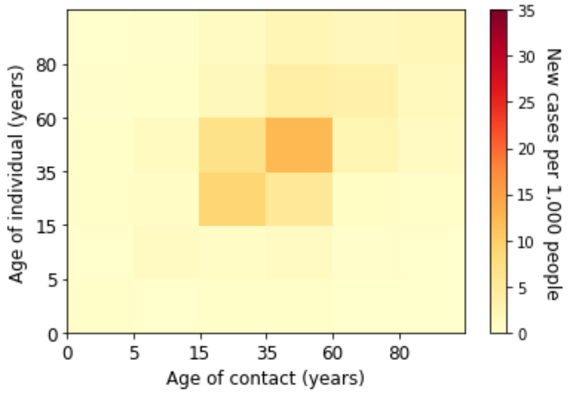


*(u) Week 5 of 2021*

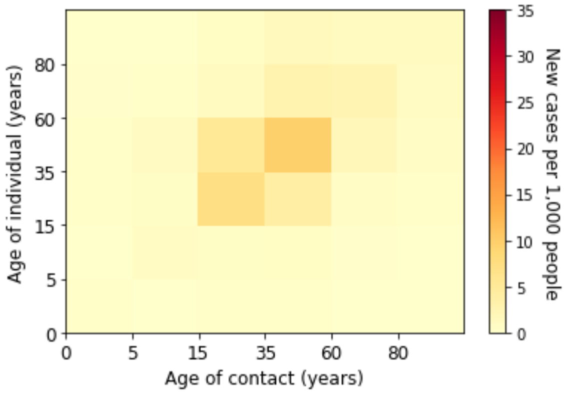


*(v) Week 6 of 2021*

*Figure 8. Estimated absolute contribution of transmission for each age groups in the second wave*

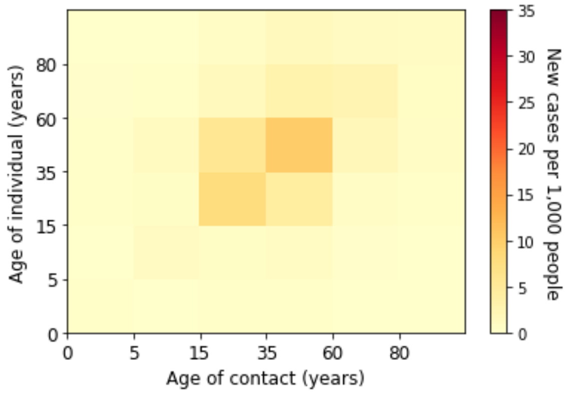


*(a) Week 7 of 2021*

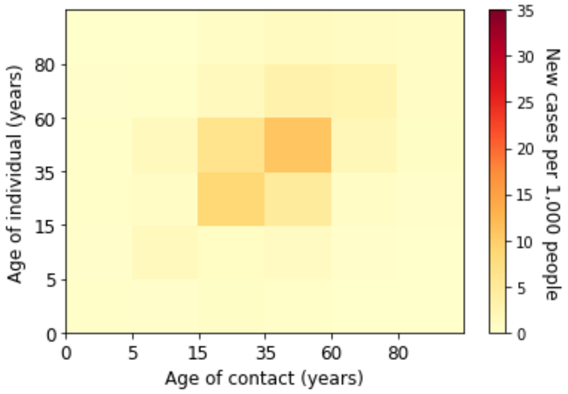


*(b) Week 8 of 2021*

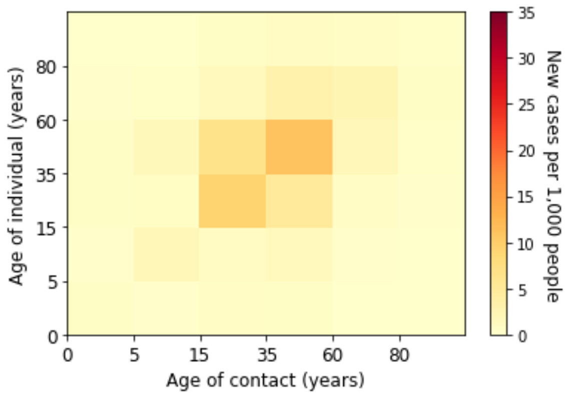


*(c) Week 9 of 2021*

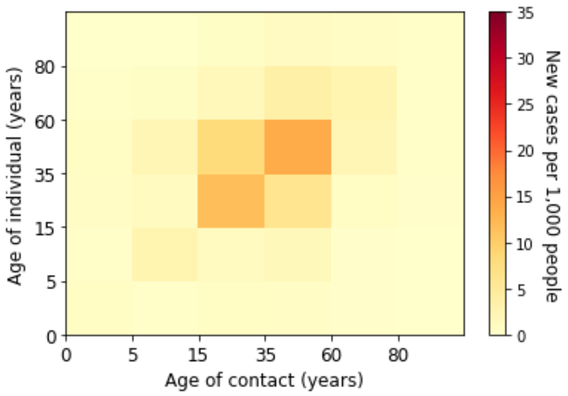


*(d) Week 10 of 2021*

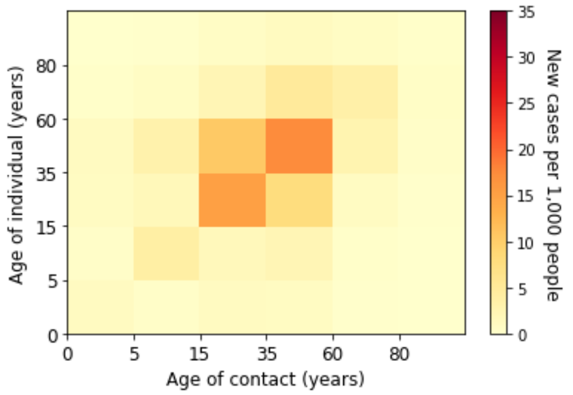


*(e) Week 11 of 2021*

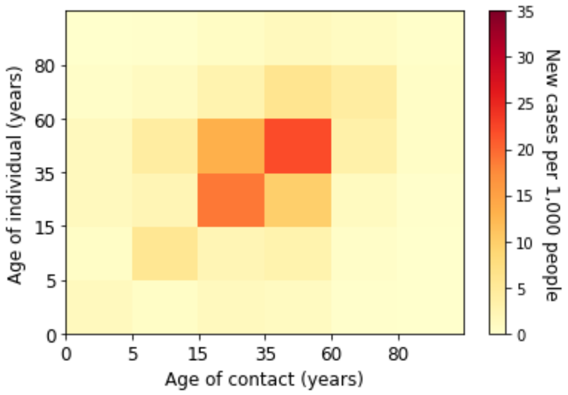


*(f) Week 12 of 2021*

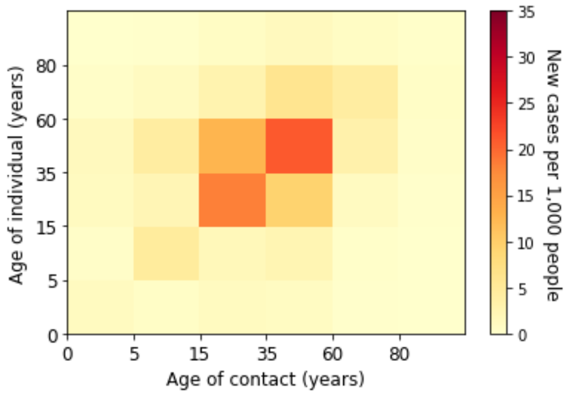


*(g) Week 13 of 2021*

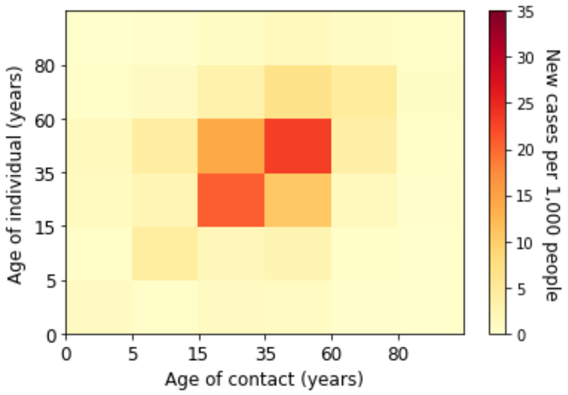


*(h) Week 14 of 2021*

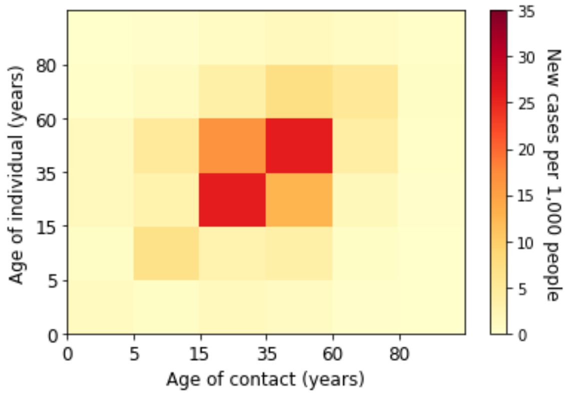


*(i) Week 15 of 2021*

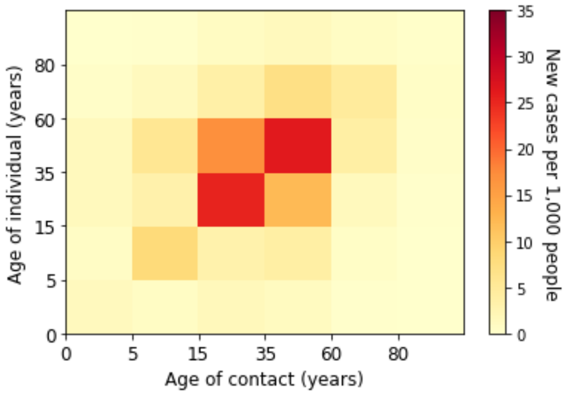


*(j) Week 16 of 2021*

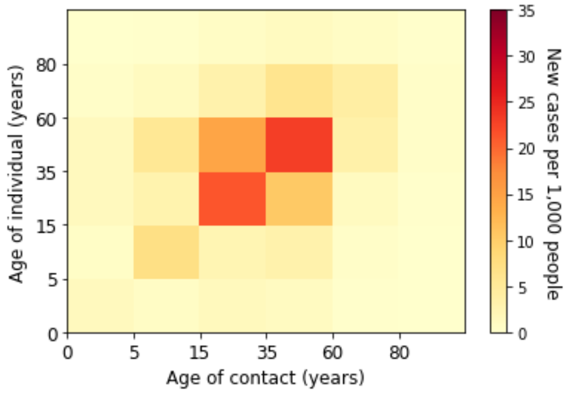


*(k) Week 17 of 2021*

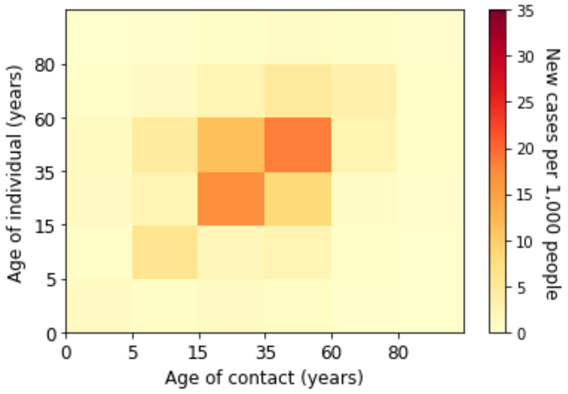


*(l) Week 18 of 2021*

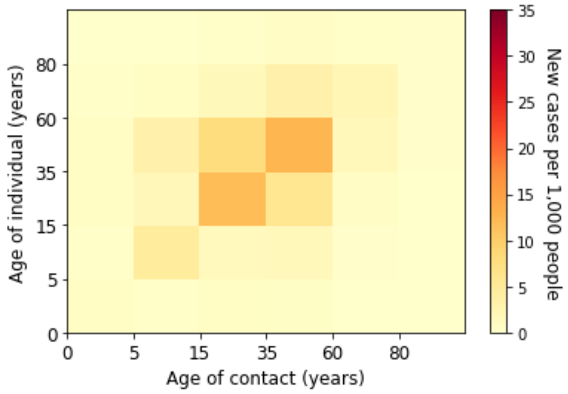


*(m) Week 19 of 2021*

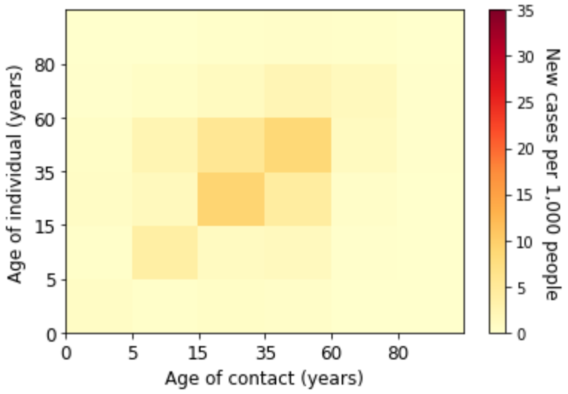


*(n) Week 20 of 2021*

*Figure 9. Estimated absolute contribution of transmission for each age groups in the third wave*

### Supplementary E. Estimated Marginal Force of Infection in Contacts with the Underdetection Ratios

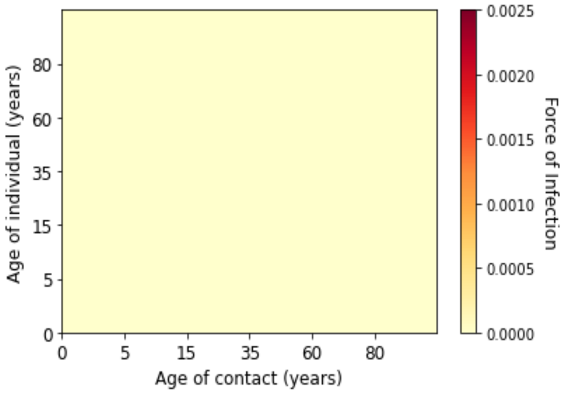


*(a) Week 6 of 2020*

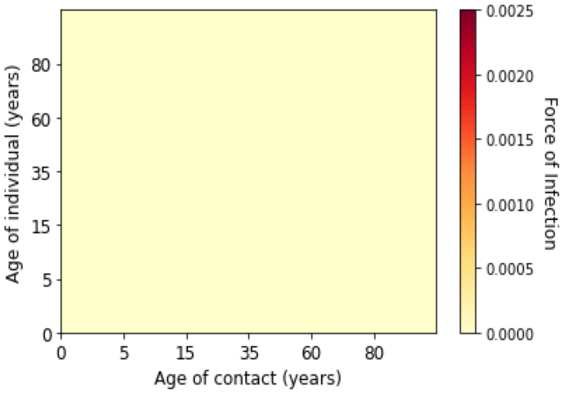


*(b) Week 7 of 2020*

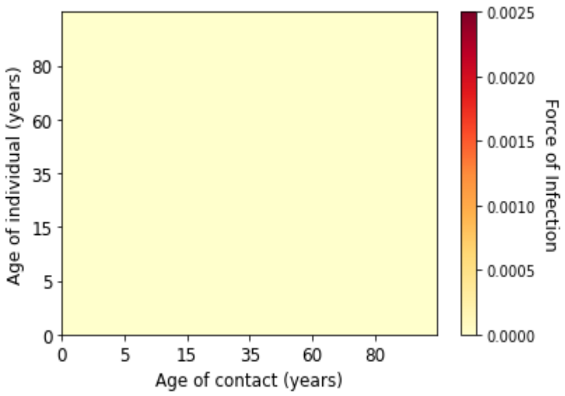


*(c) Week 8 of 2020*

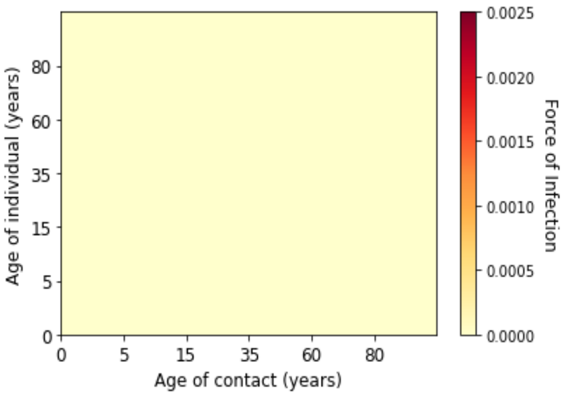


*(d) Week 9 of 2020*

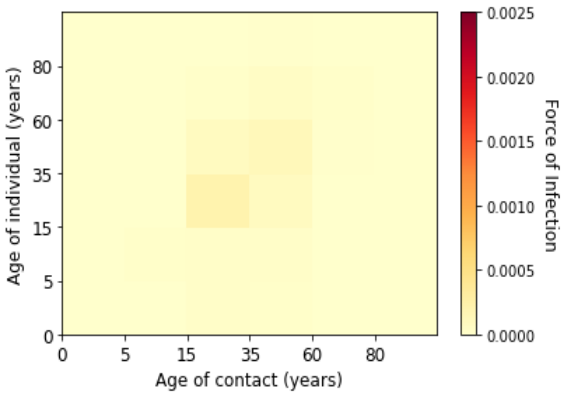


*(e) Week 10 of 2020*

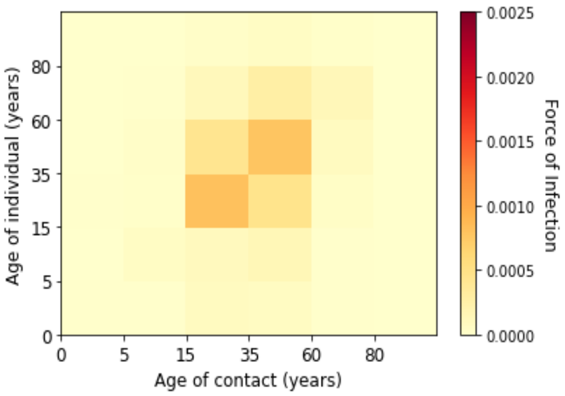


*(f) Week 11 of 2020*

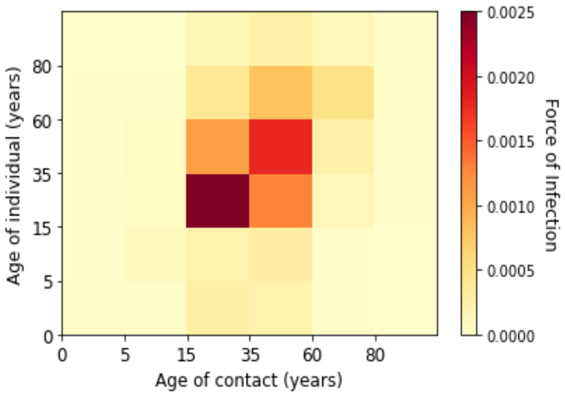


*(g) Week 12 of 2020*

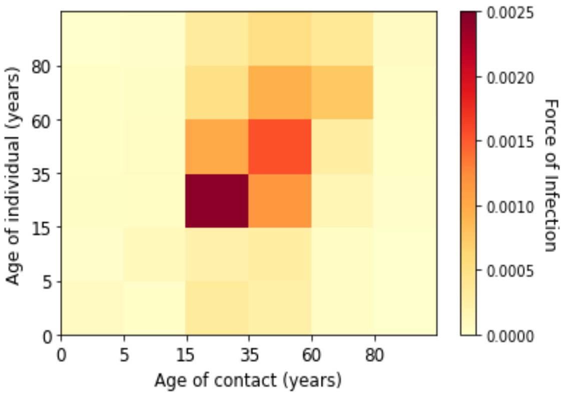


*(h) Week 13 of 2020*

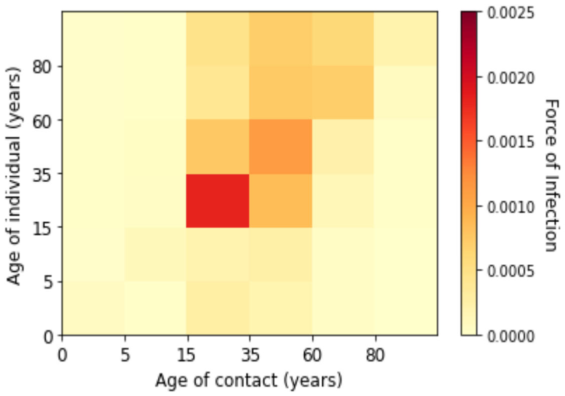


*(i) Week 14 of 2020*

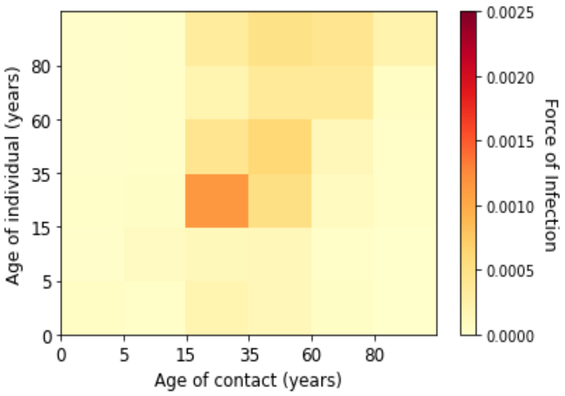


*(j) Week 15 of 2020*

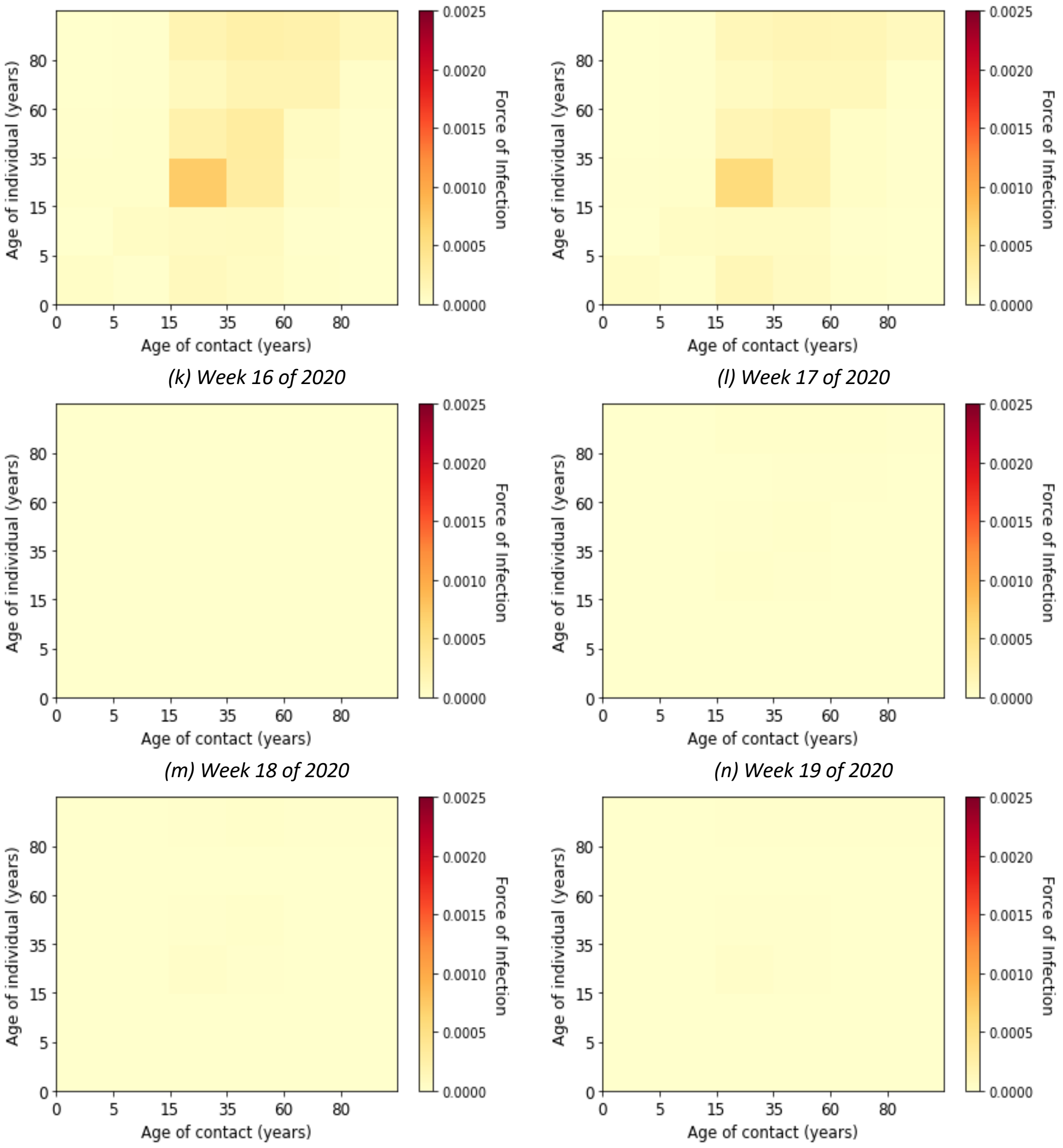


(k) Week 16 of 2020

(l) Week 17 of 2020

(m) Week 18 of 2020

(n) Week 19 of 2020

(o) Week 20 of 2020

(p) Week 21 of 2020

Figure 10. Estimated marginal force of infection for each age groups in the first wave

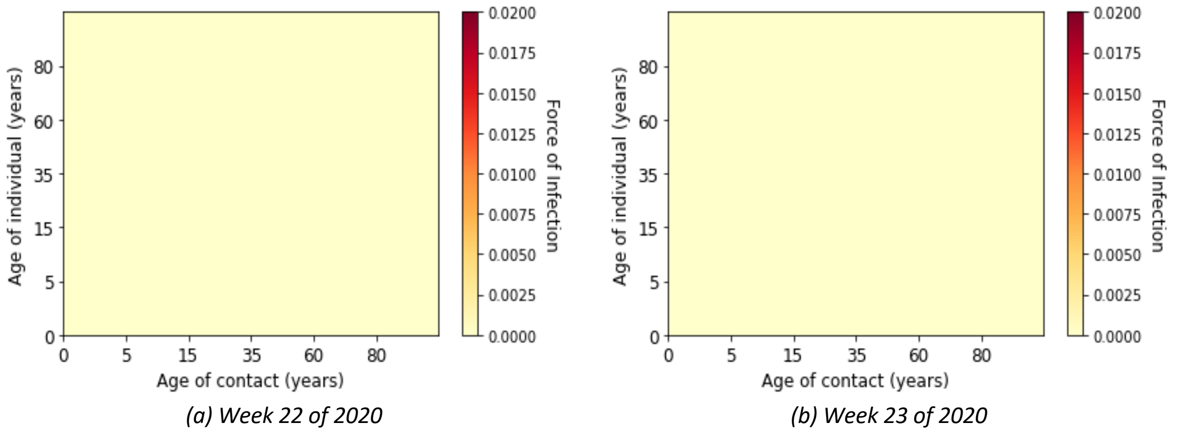


(a) Week 22 of 2020

(b) Week 23 of 2020

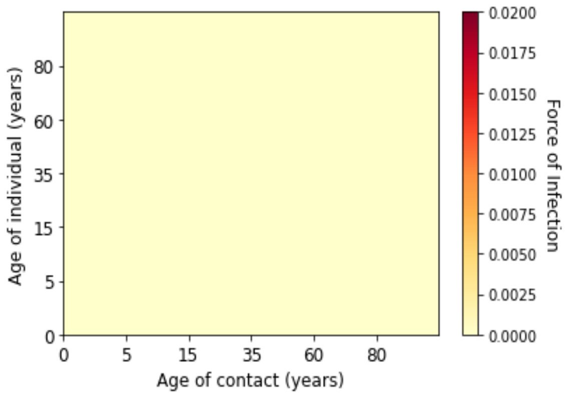


*(c) Week 24 of 2020*

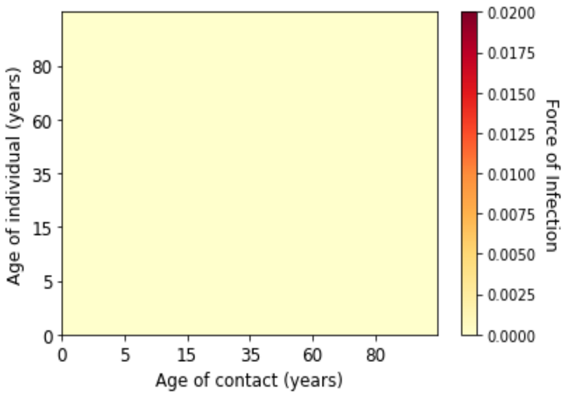


*(d) Week 25 of 2020*

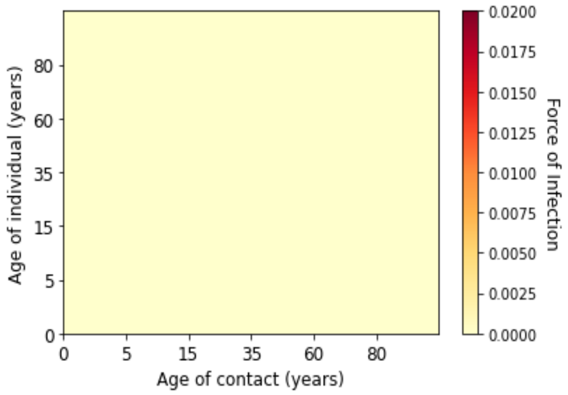


*(e) Week 26 of 2020*

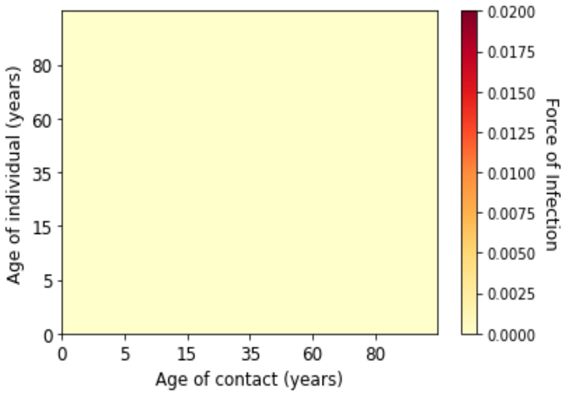


*(f) Week 27 of 2020*

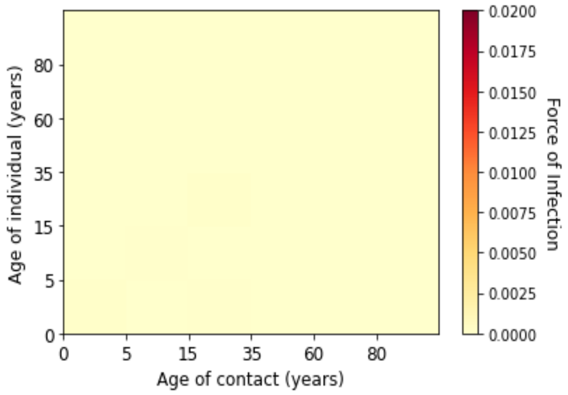


*(g) Week 28 of 2020*

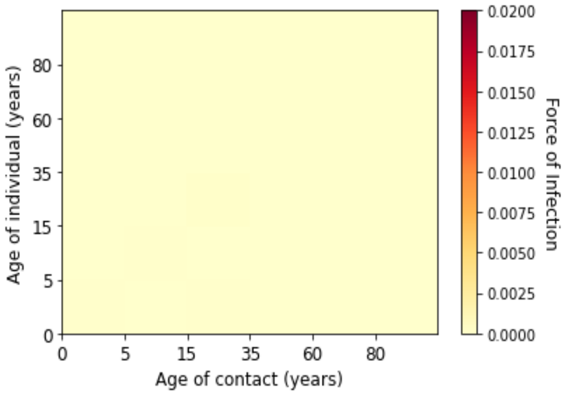


*(h) Week 29 of 2020*

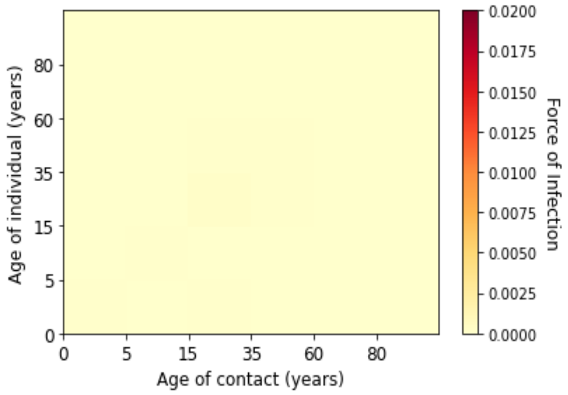


*(i) Week 30 of 2020*

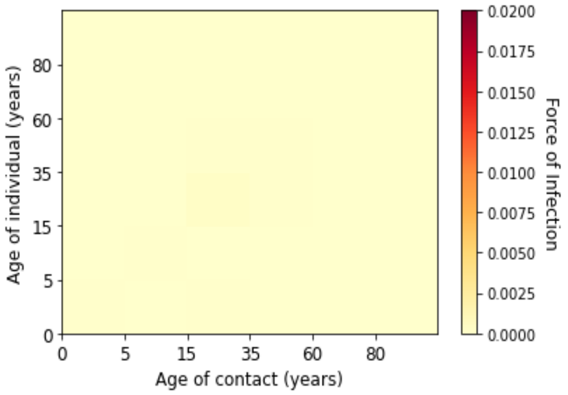


*(j) Week 31 of 2020*

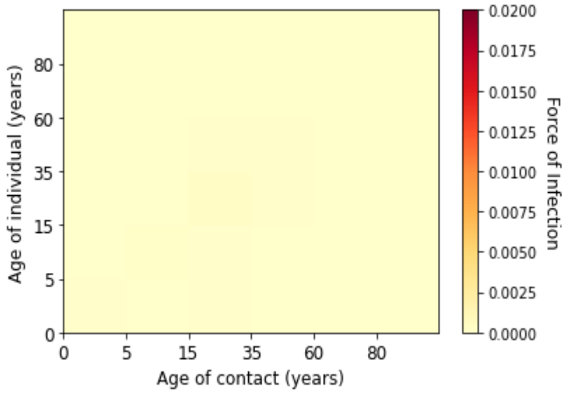


*(k) Week 32 of 2020*

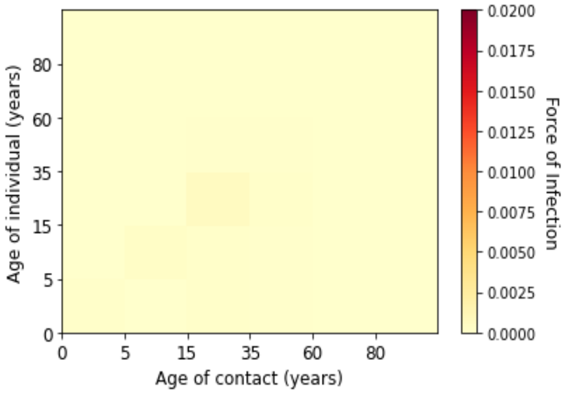


*(l) Week 33 of 2020*

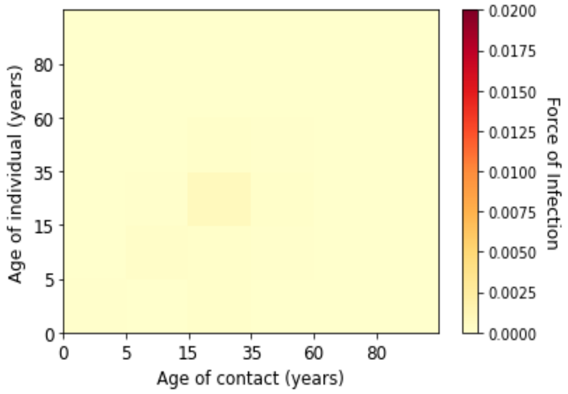


*(m) Week 34 of 2020*

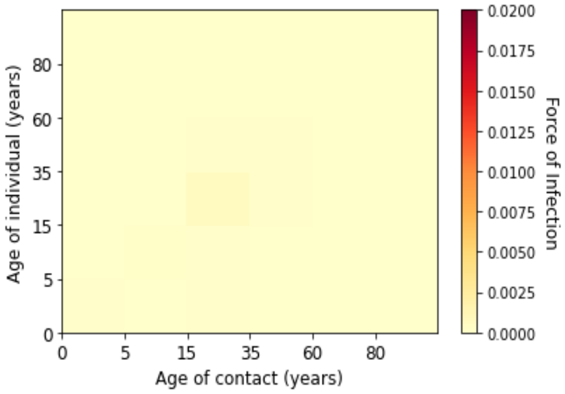


*(n) Week 35 of 2020*

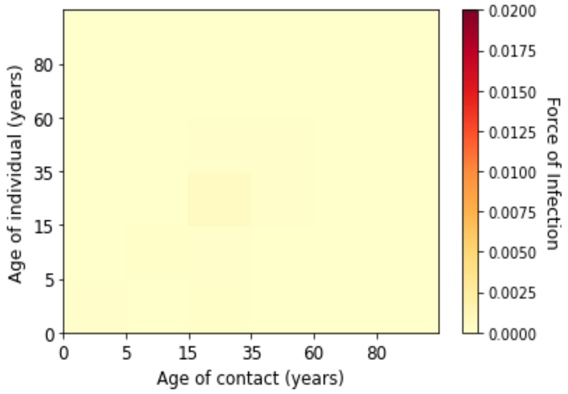


*(o) Week 36 of 2020*

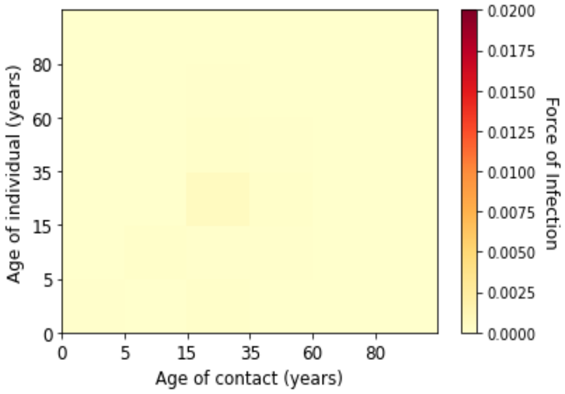


*(p) Week 37 of 2020*

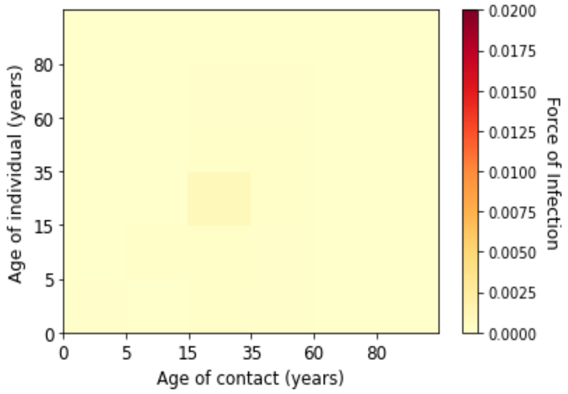


*(q) Week 38 of 2020*

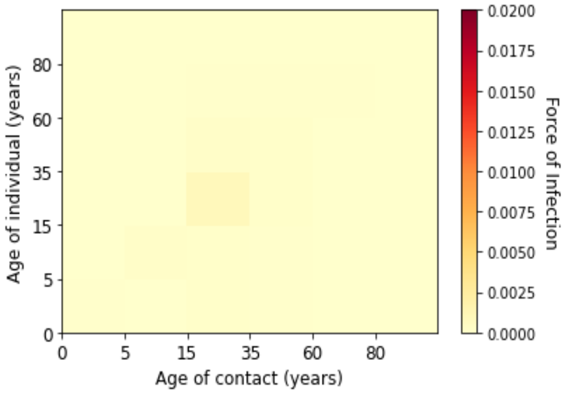


*(r) Week 39 of 2020*

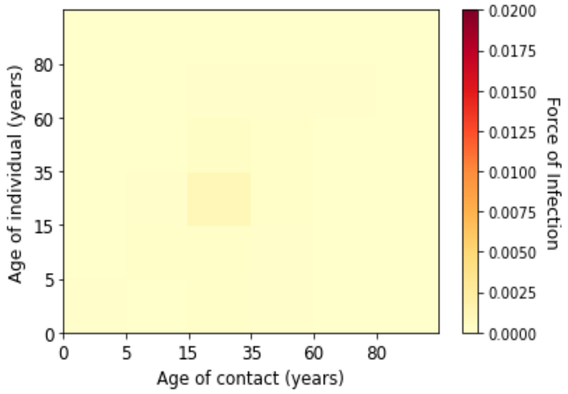


*(s) Week 40 of 2020*

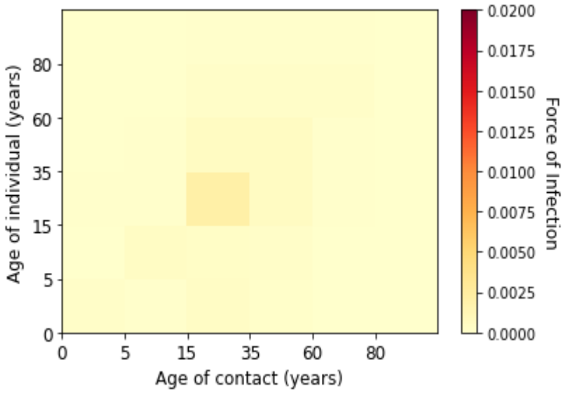


*(t) Week 41 of 2020*

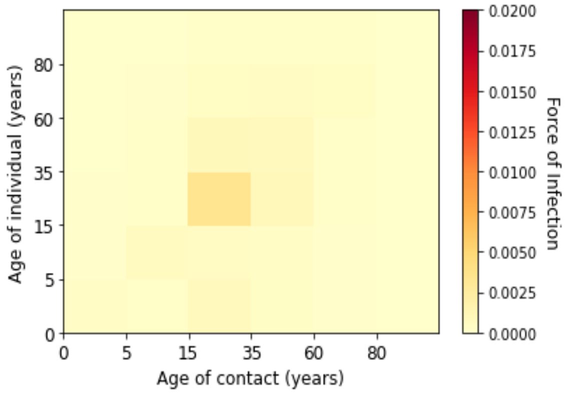


*(u) Week 42 of 2020*

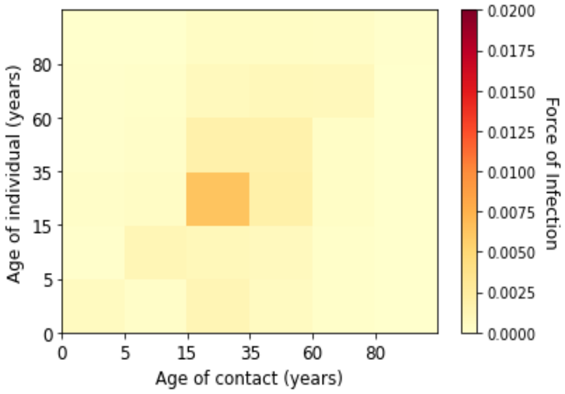


*(v) Week 43 of 2020*

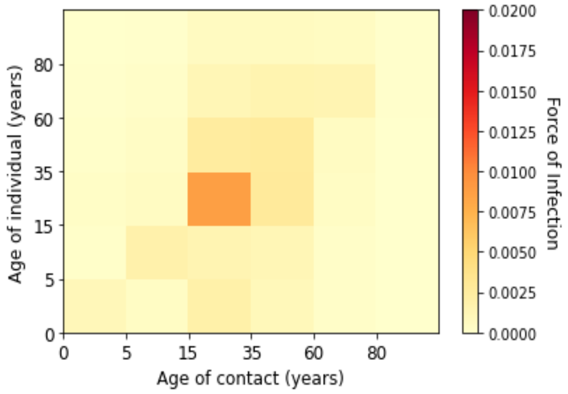


*(w) Week 44 of 2020*

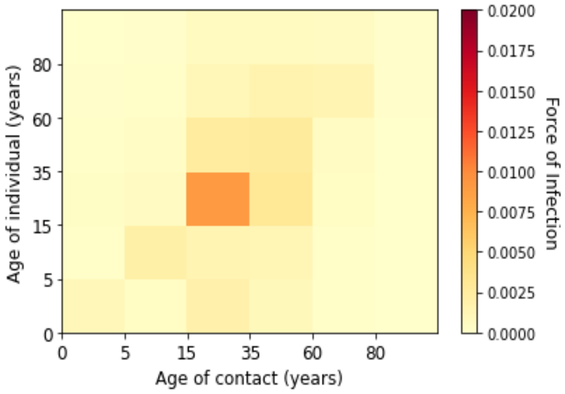


*(x) Week 45 of 2020*

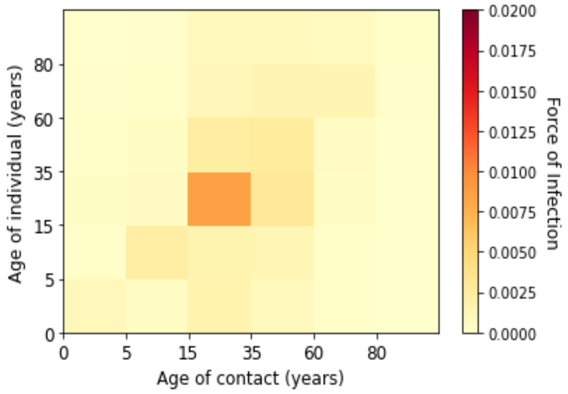


*(y) Week 46 of 2020*

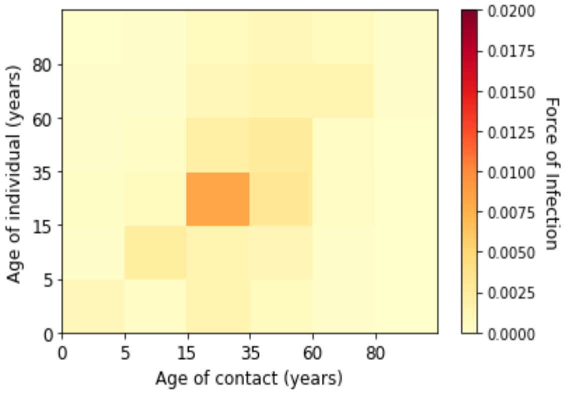


*(z) Week 47 of 2020*

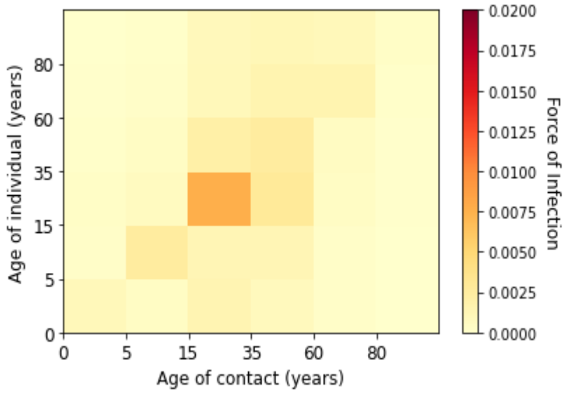


*(aa) Week 48 of 2020*

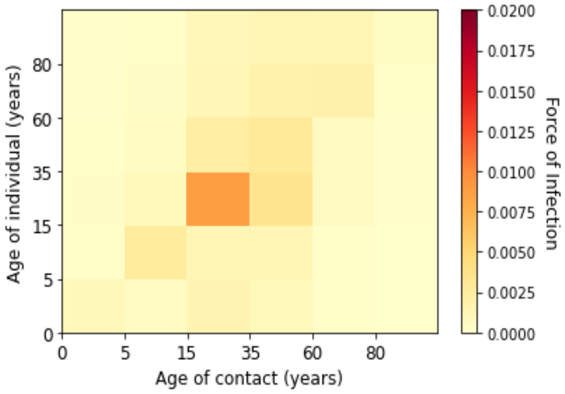


*(ab) Week 49 of 2020*

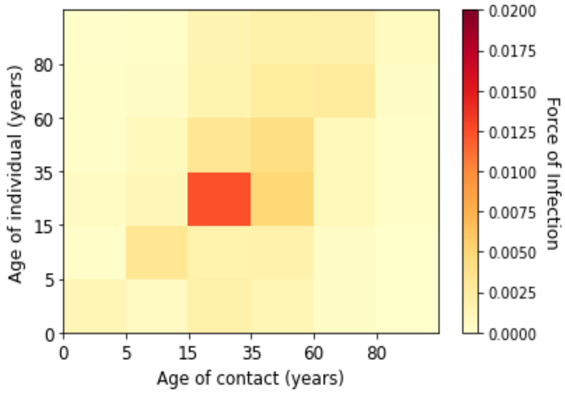


*(ac) Week 50 of 2020*

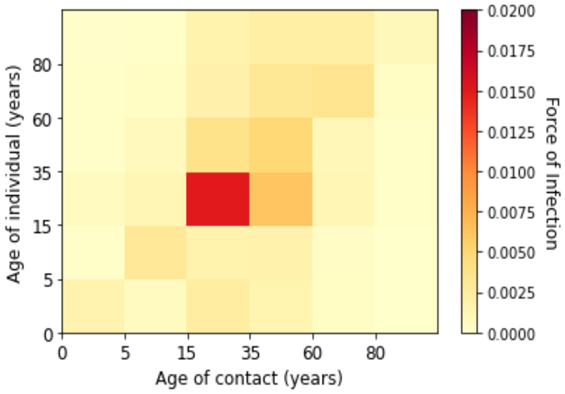


*(ad) Week 51 of 2020*

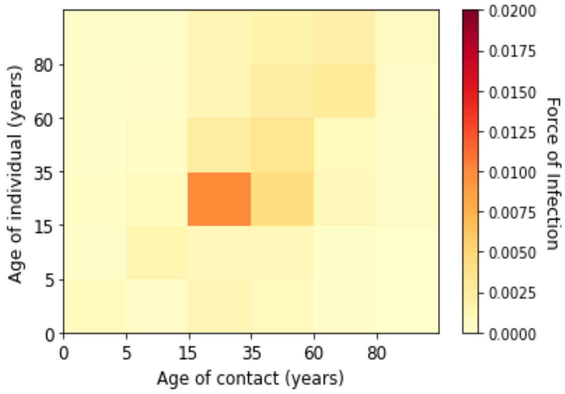


*(ae) Week 52 of 2020*

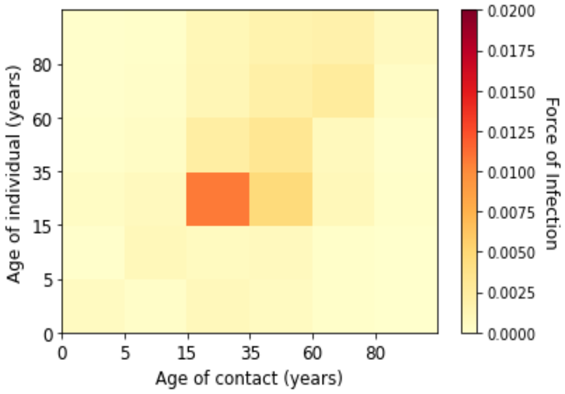


*(af) Week 53 of 2020*

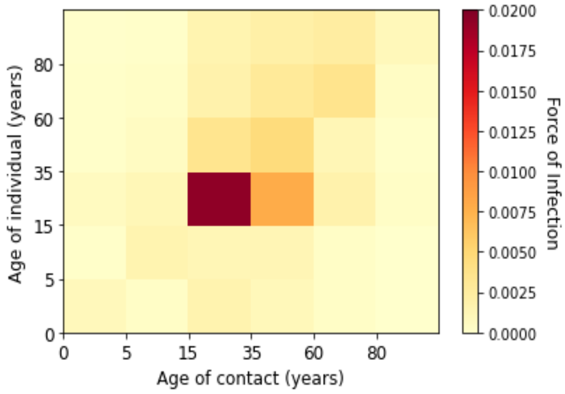


*(ag) Week 1 of 2021*

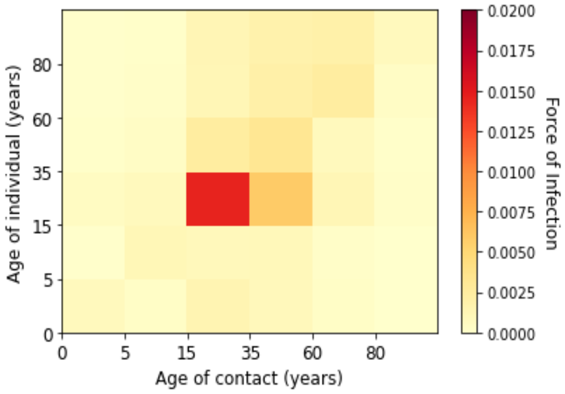


*(ah) Week 2 of 2021*

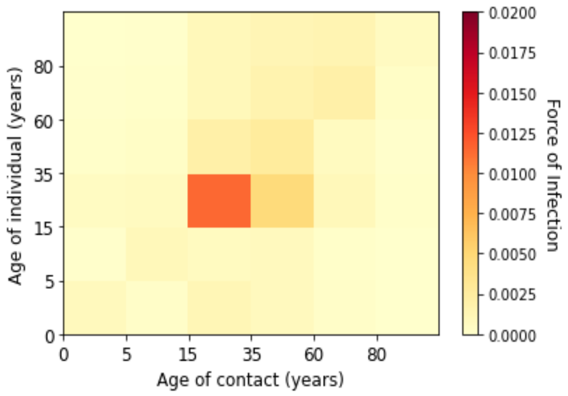


*(ai) Week 3 of 2021*

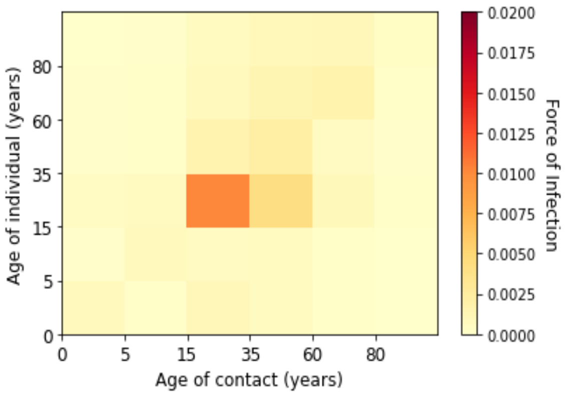


*(aj) Week 4 of 2021*

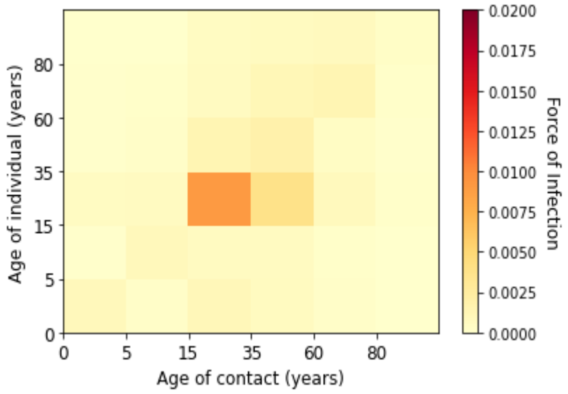


*(ak) Week 5 of 2021*

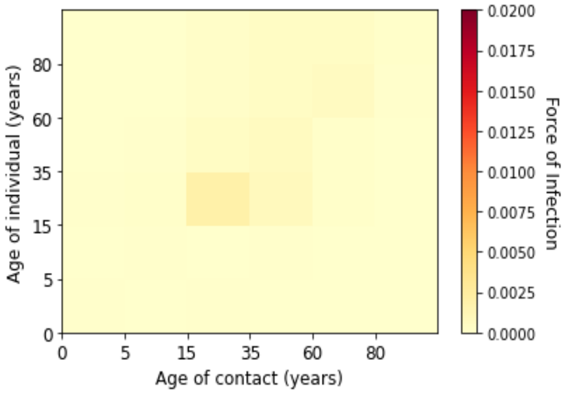


*(al) Week 6 of 2021*

*Figure 11. Estimated marginal force of infection for each age groups in the second wave*

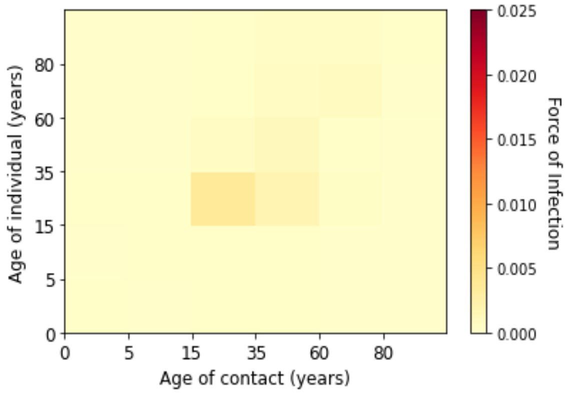


*(a) Week 7 of 2021*

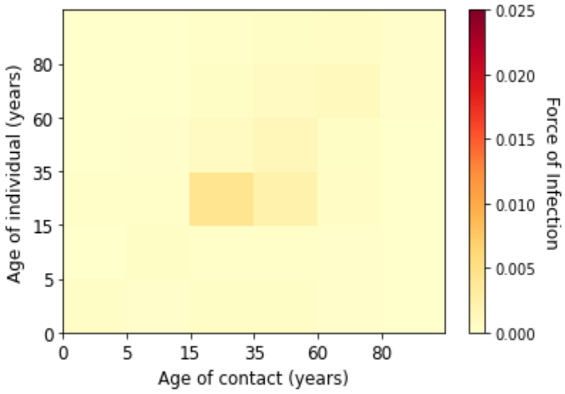


*(b) Week 8 of 2021*

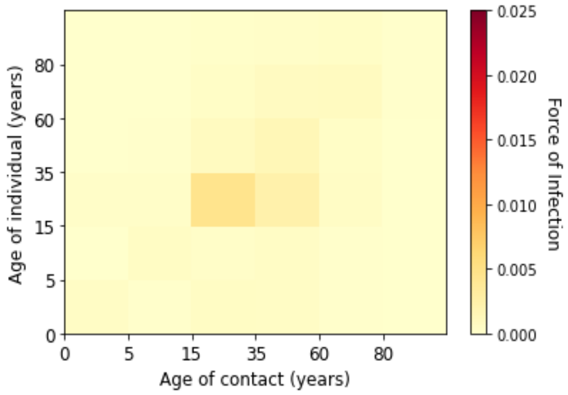


*(c) Week 9 of 2021*

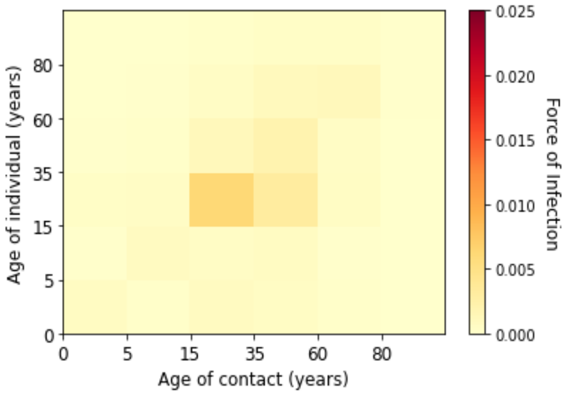


*(d) Week 10 of 2021*

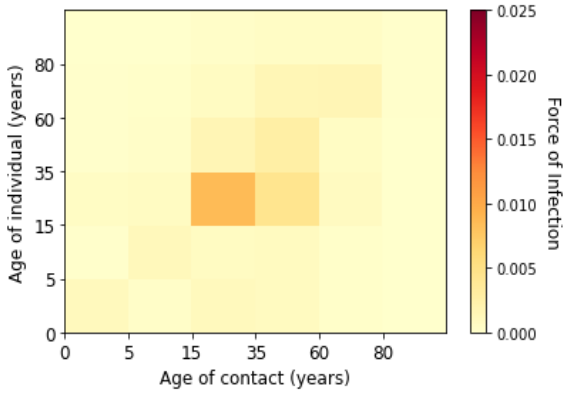


*(e) Week 11 of 2021*

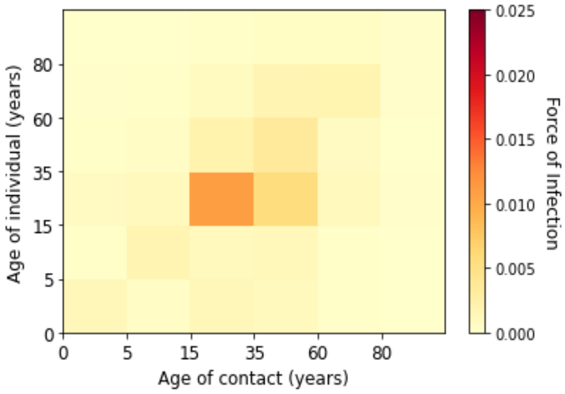


*(f) Week 12 of 2021*

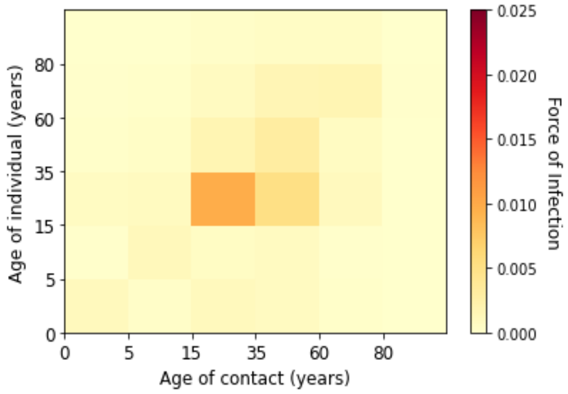


*(g) Week 13 of 2021*

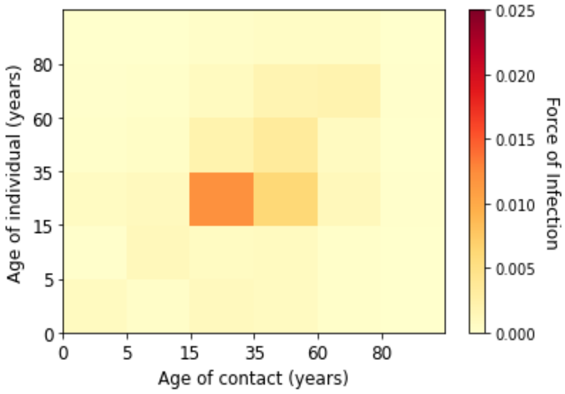


*(h) Week 14 of 2021*

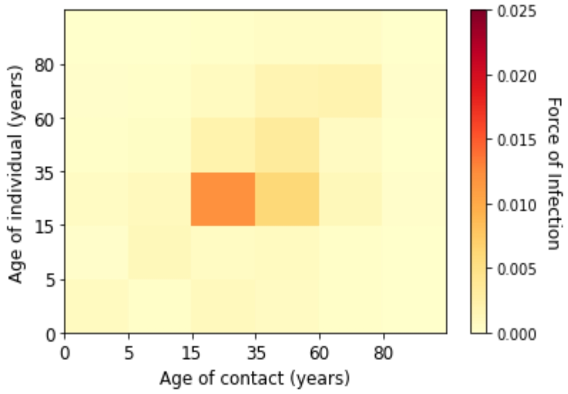


*(i) Week 15 of 2021*

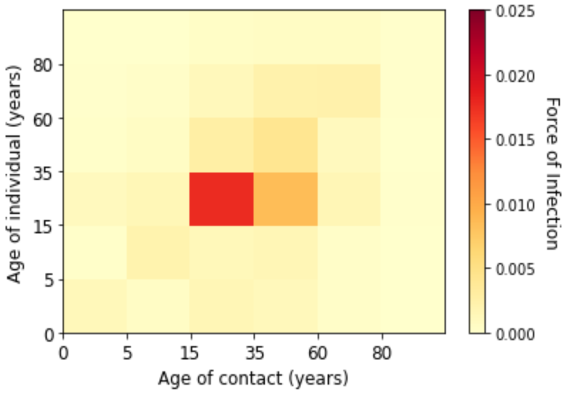


*(j) Week 16 of 2021*

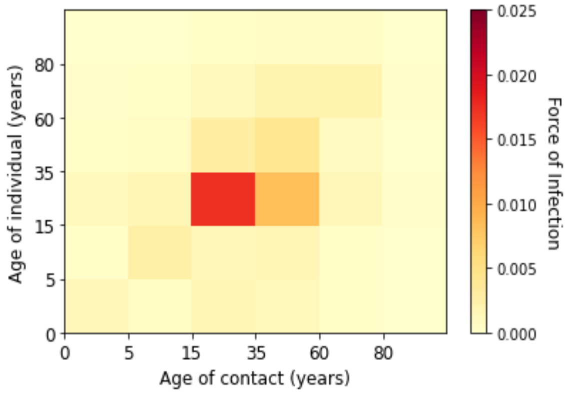


*(k) Week 17 of 2021*

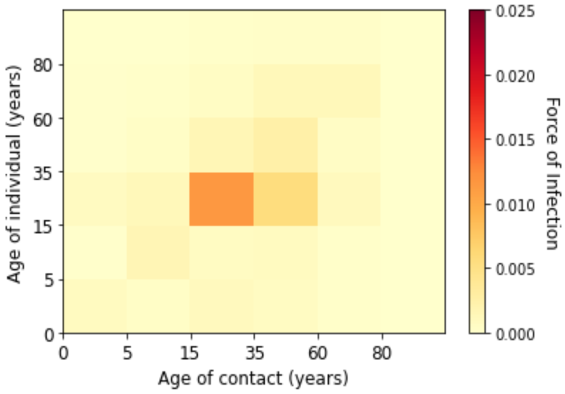


*(l) Week 18 of 2021*

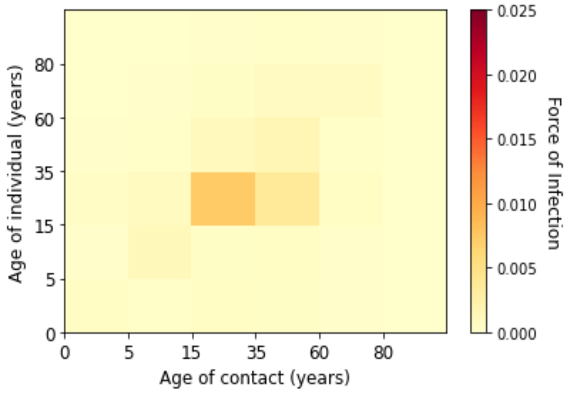


*(m) Week 19 of 2021*

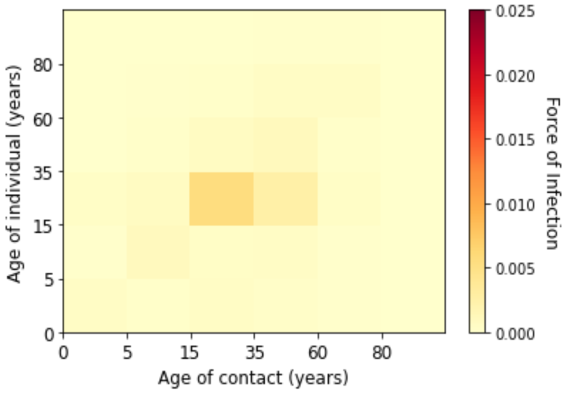


*(n) Week 20 of 2021*

*Figure 12. Estimated marginal fore of infection for each age groups in the third wave*

### Supplementary F. Estimated Contribution of Contact to Transmission with the Underdetection Ratios

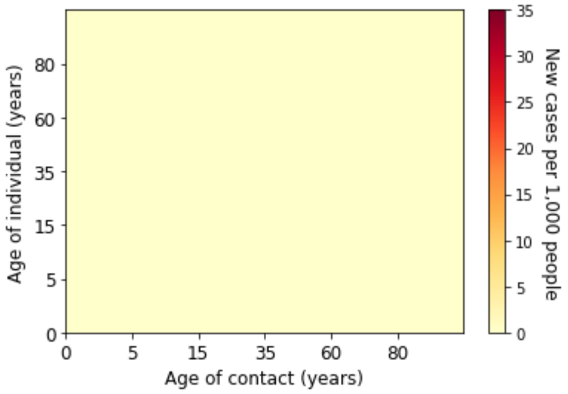


*(a) Week 6 of 2020*

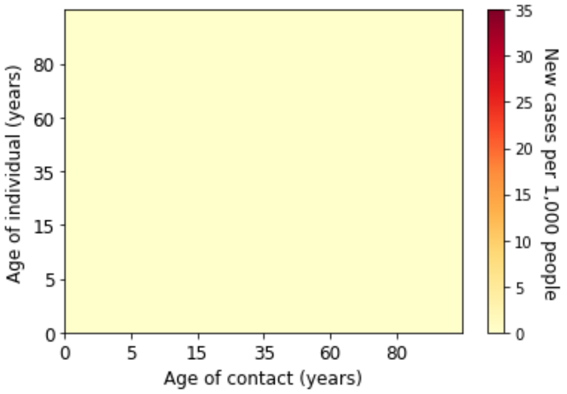


*(b) Week 7 of 2020*

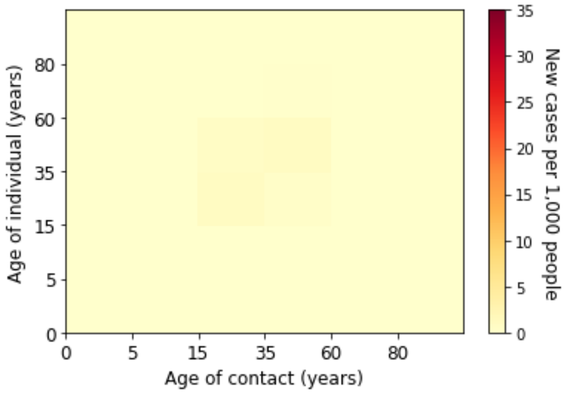


*(c) Week 8 of 2020*

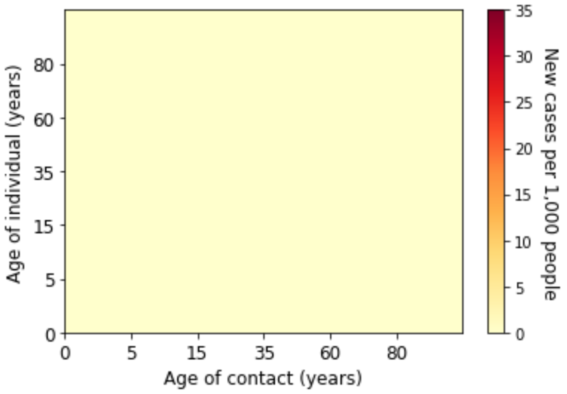


*(d) Week 9 of 2020*

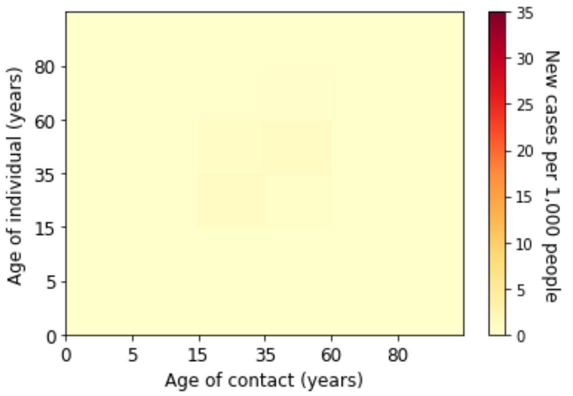


*(e) Week 10 of 2020*

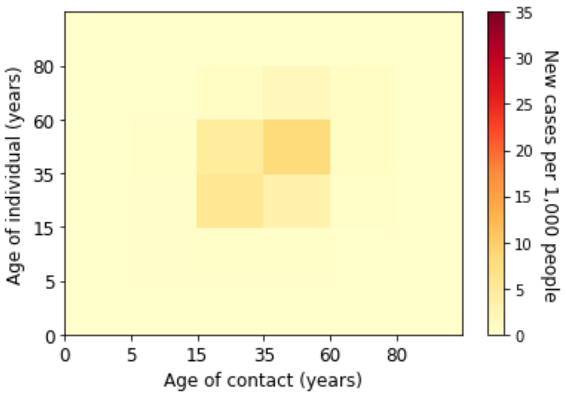


*(f) Week 11 of 2020*

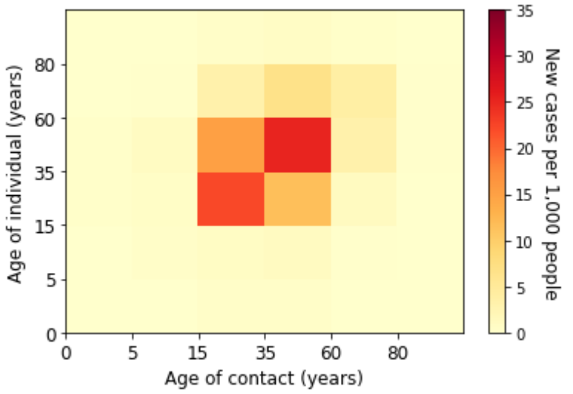


*(g) Week 12 of 2020*

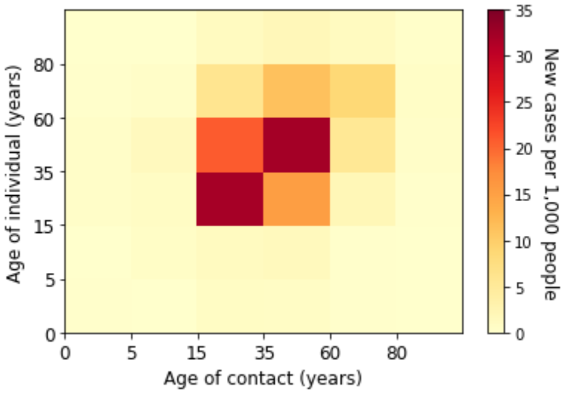


*(h) Week 13 of 2020*

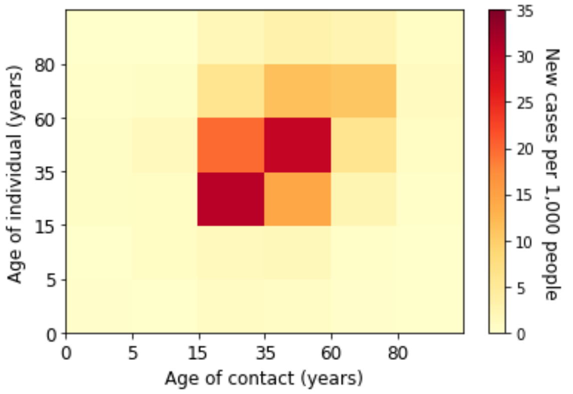


*(i) Week 14 of 2020*

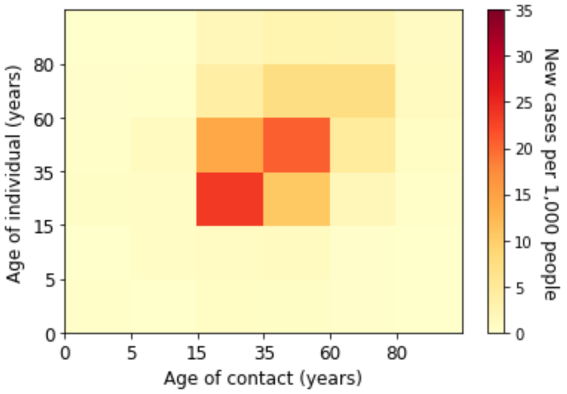


*(j) Week 15 of 2020*

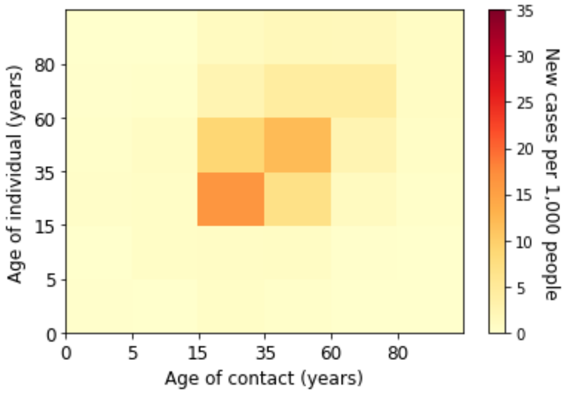


*(k) Week 16 of 2020*

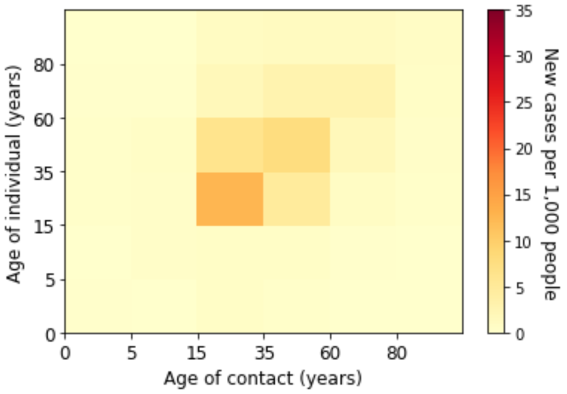


*(l) Week 17 of 2020*

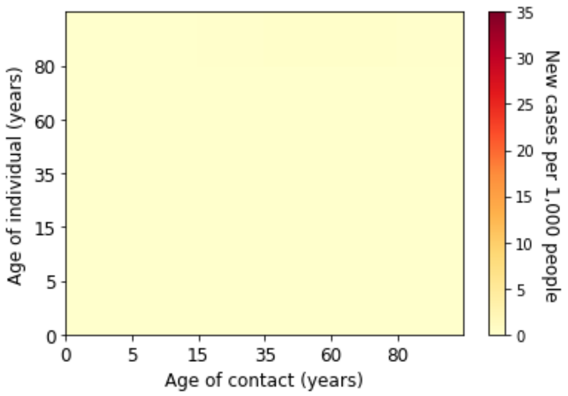


*(m) Week 18 of 2020*

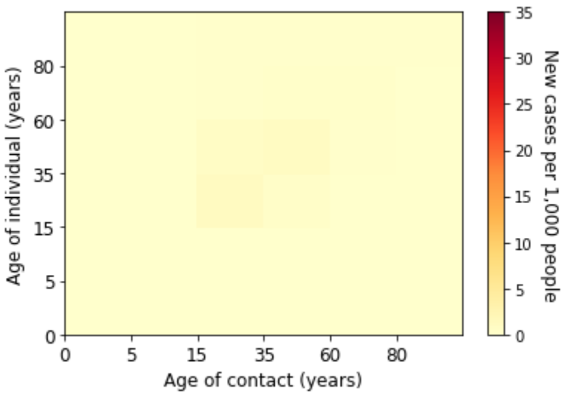


*(n) Week 19 of 2020*

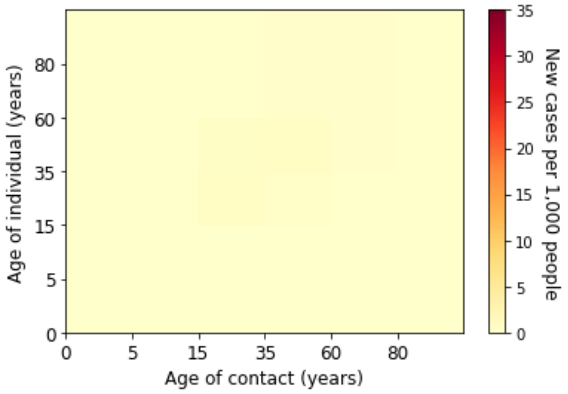


*(o) Week 20 of 2020*

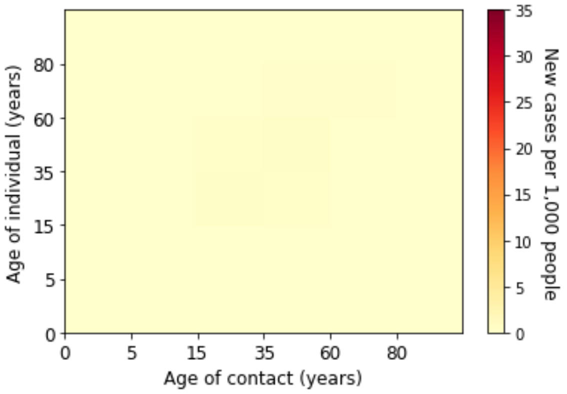


*(p) Week 21 of 2020*

*Figure 13. Estimated absolute contribution of transmission for each age groups in the first wave*

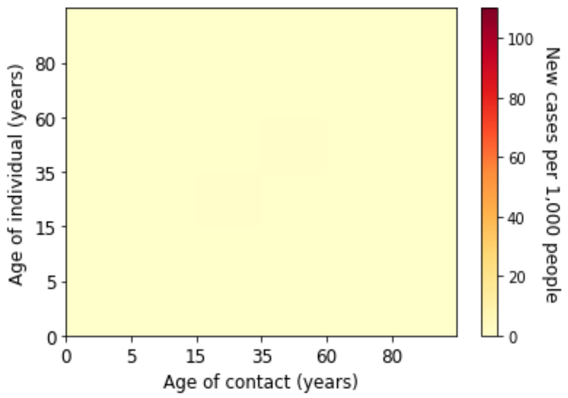


*(a) Week 22 of 2020*

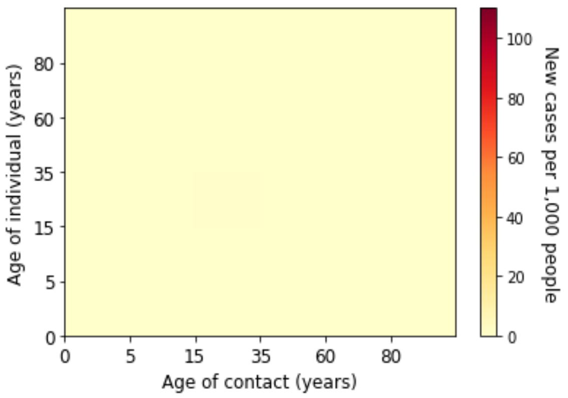


*(b) Week 23 of 2020*

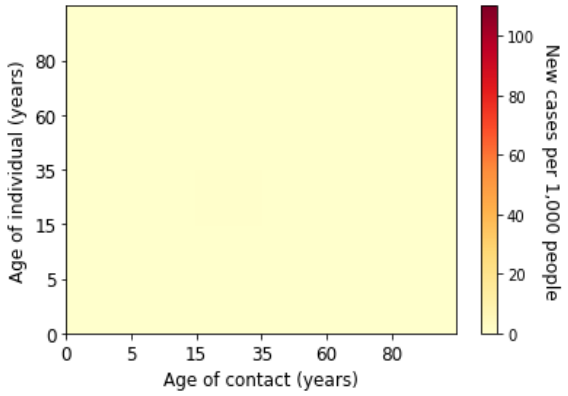


*(c) Week 24 of 2020*

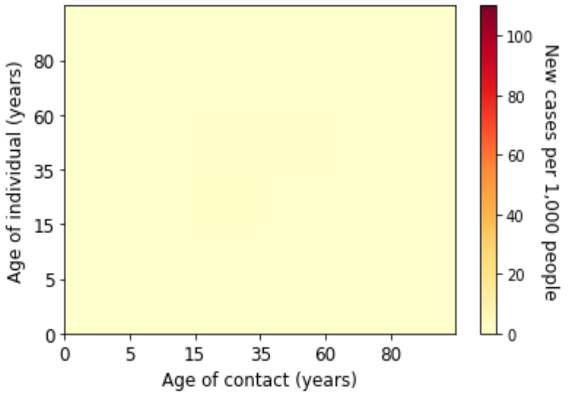


*(d) Week 25 of 2020*

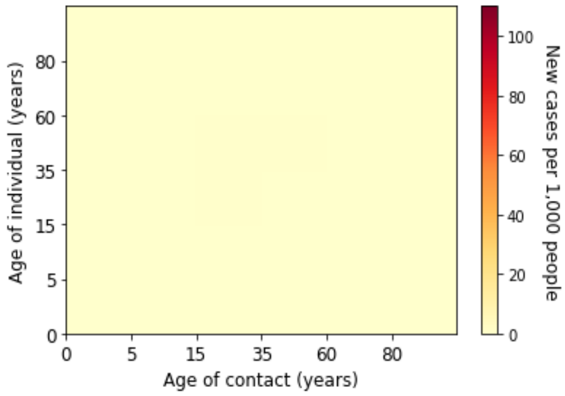


*(e) Week 26 of 2020*

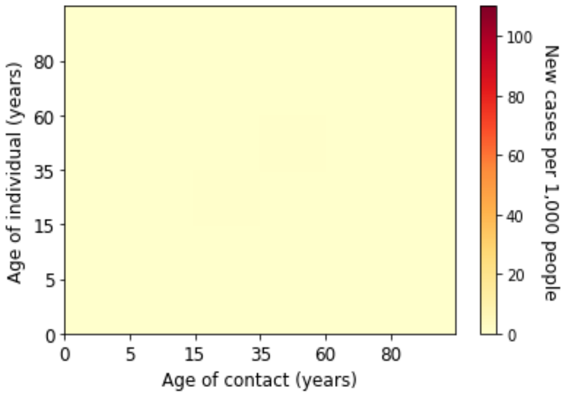


*(f) Week 27 of 2020*

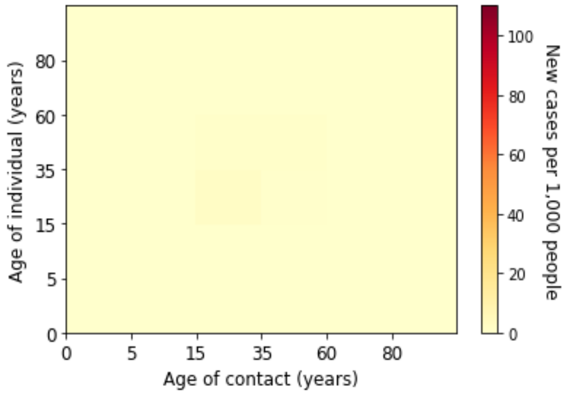


*(g) Week 28 of 2020*

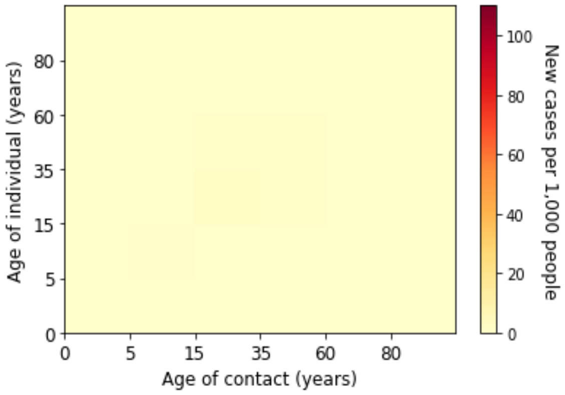


*(h) Week 29 of 2020*

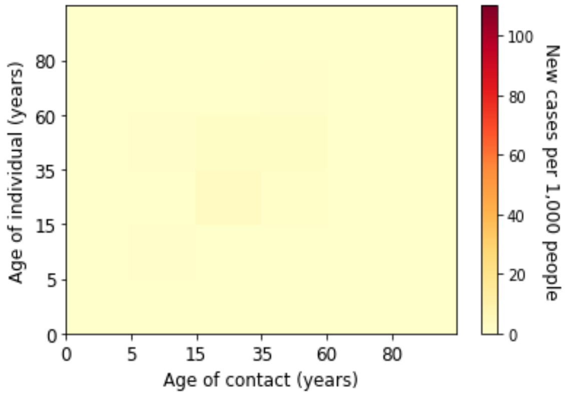


*(i) Week 30 of 2020*

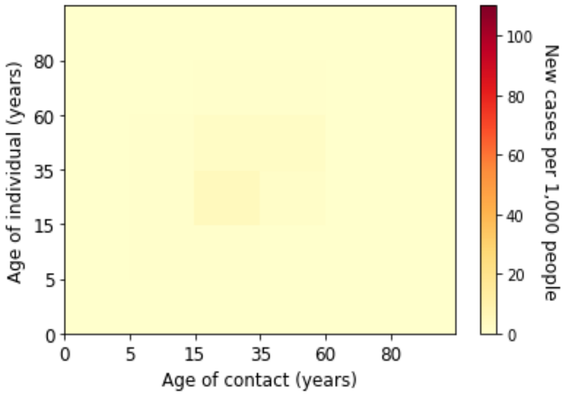


*(j) Week 31 of 2020*

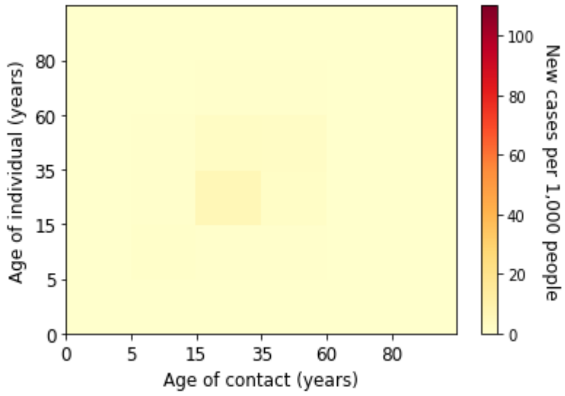


*(k) Week 32 of 2020*

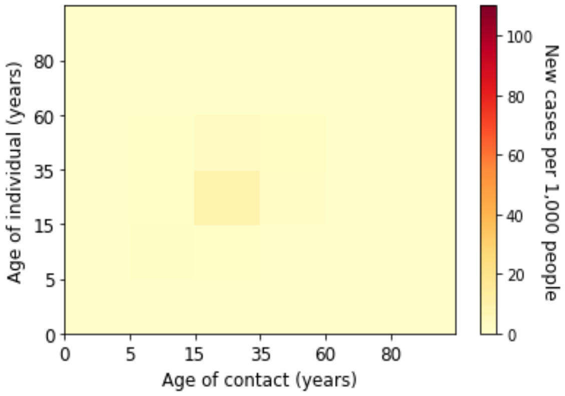


*(l) Week 33 of 2020*

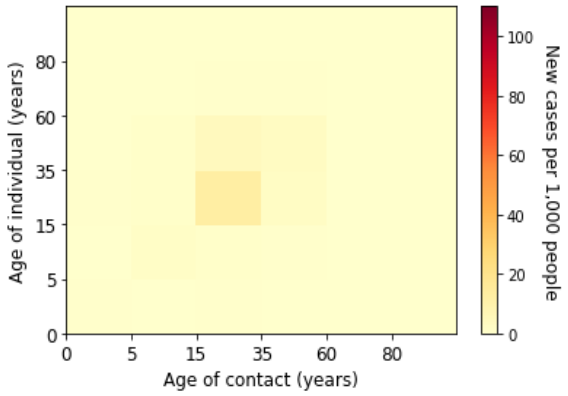

*(m) Week 34 of 2020*

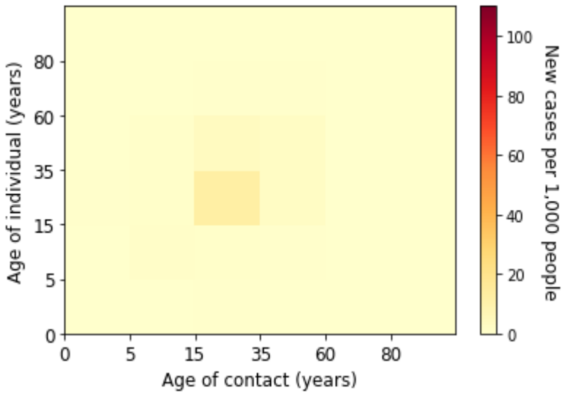

*(n) Week 35 of 2020*

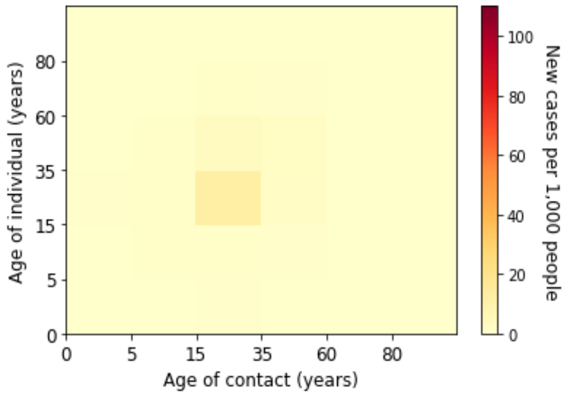

*(o) Week 36 of 2020*

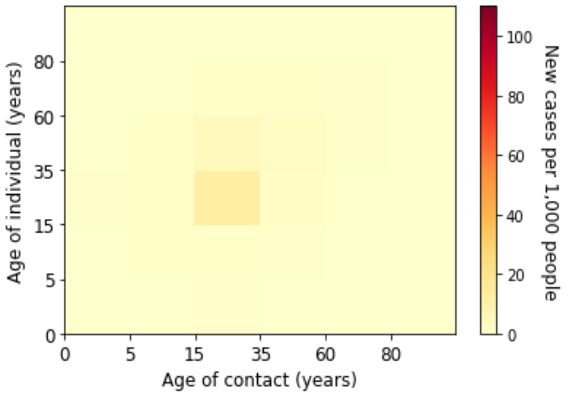

*(p) Week 37 of 2020*

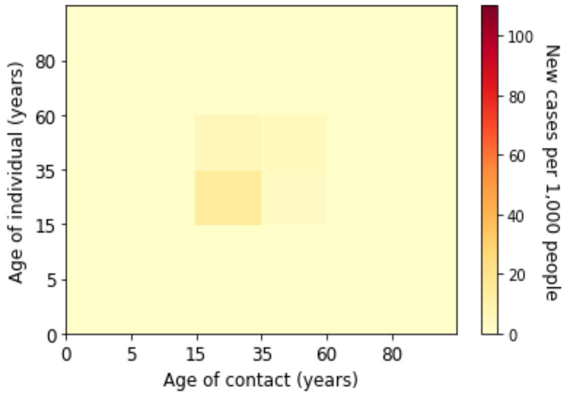

*(q) Week 38 of 2020*

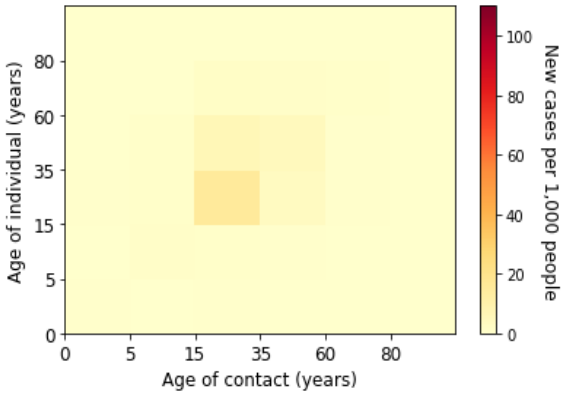

*(r) Week 39 of 2020*

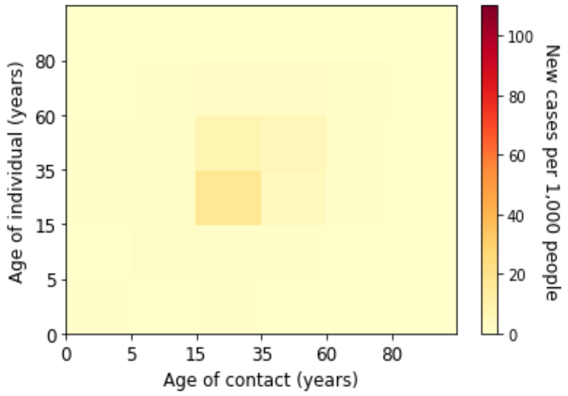

*(s) Week 40 of 2020*

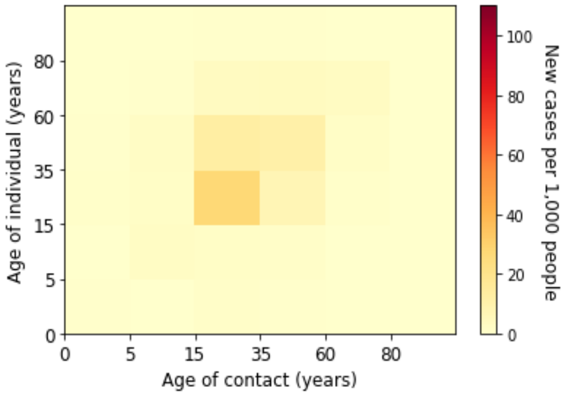

*(t) Week 41 of 2020*

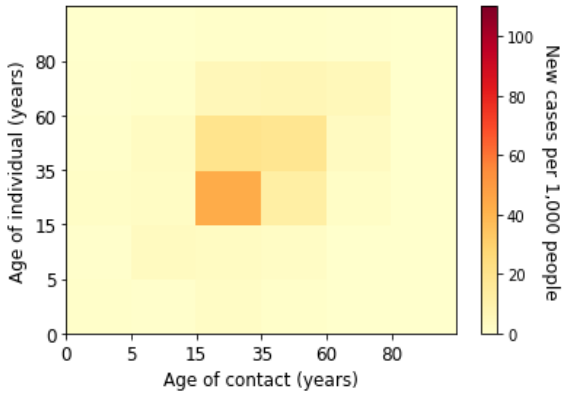


*(u) Week 42 of 2020*

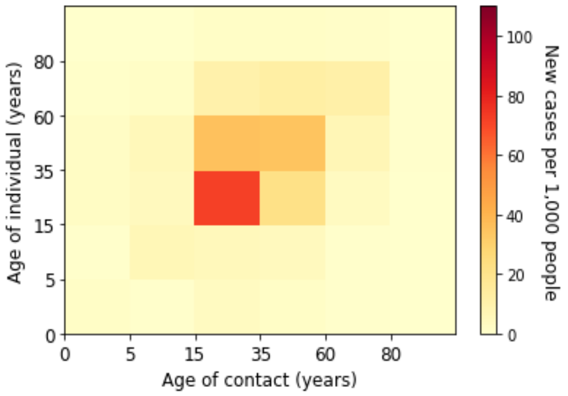


*(v) Week 43 of 2020*

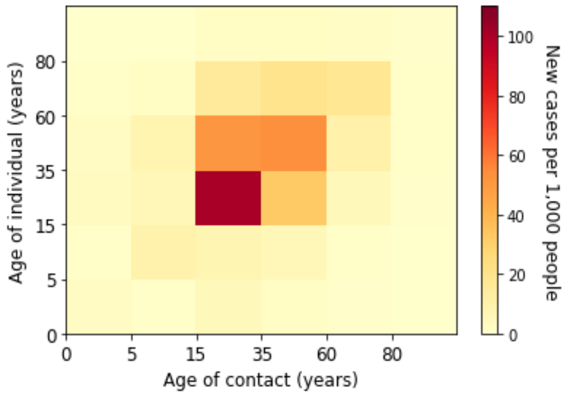


*(w) Week 44 of 2020*

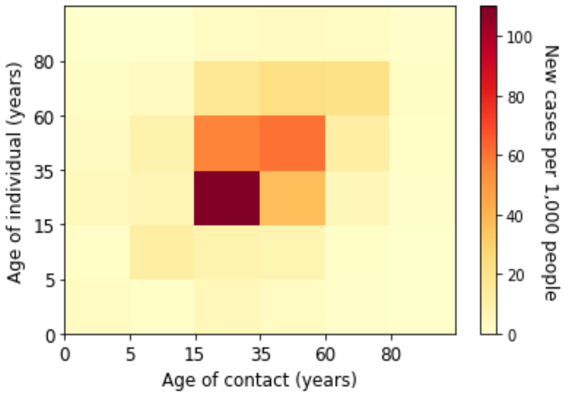


*(x) Week 45 of 2020*

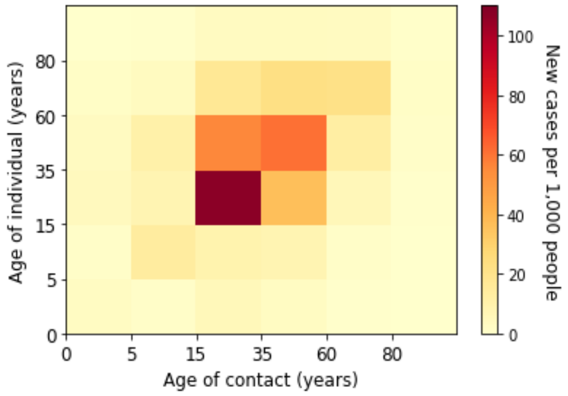


*(y) Week 46 of 2020*

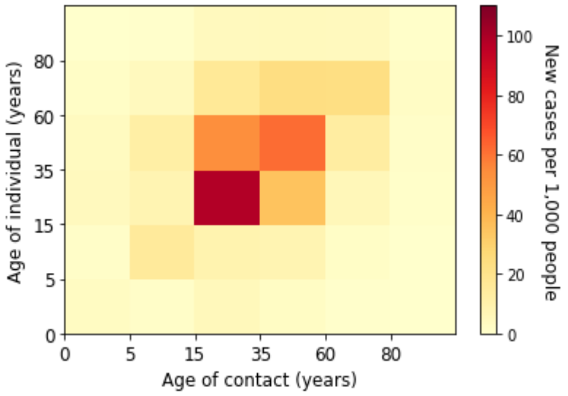


*(z) Week 47 of 2020*

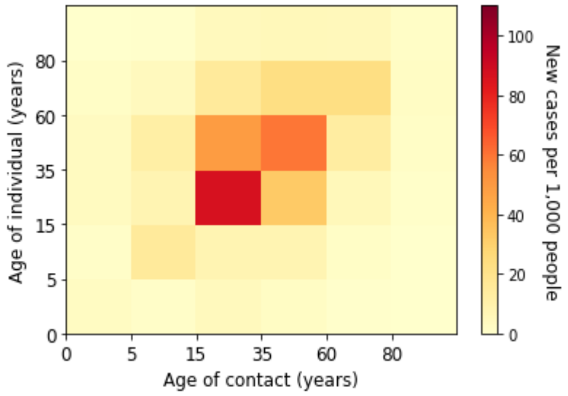


*(aa) Week 48 of 2020*

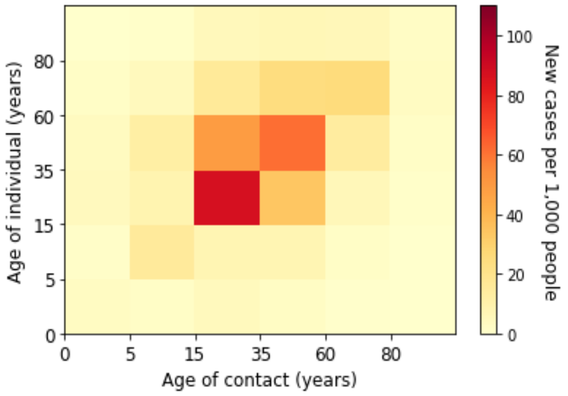


*(ab) Week 49 of 2020*

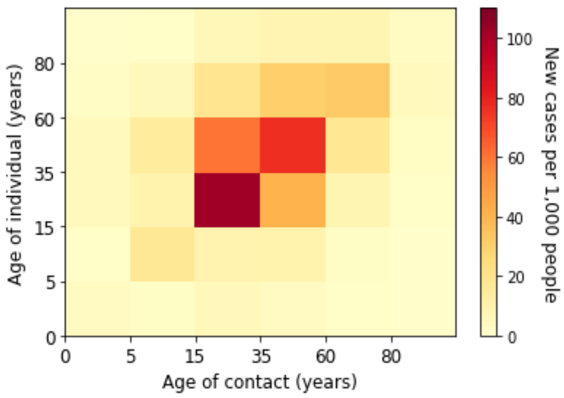


*(ac) Week 50 of 2020*

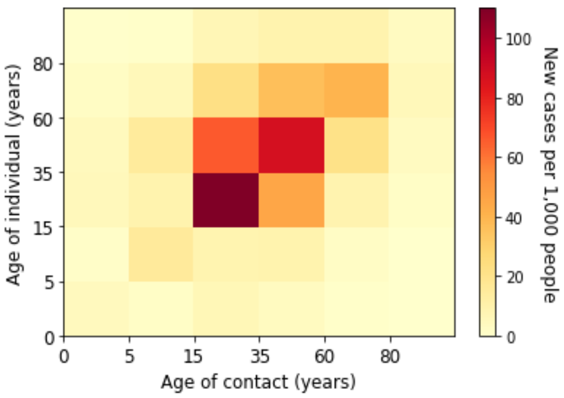


*(ad) Week 51 of 2020*

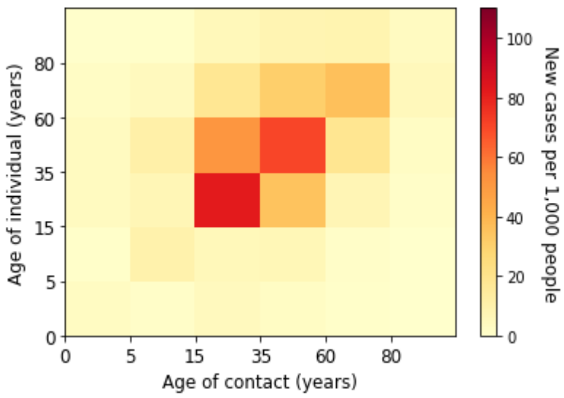


*(ae) Week 52 of 2020*

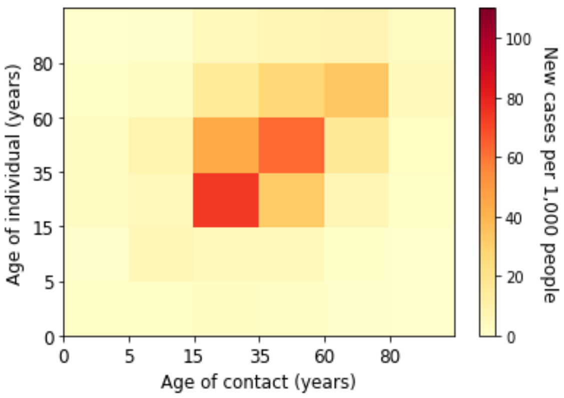


*(af) Week 53 of 2020*

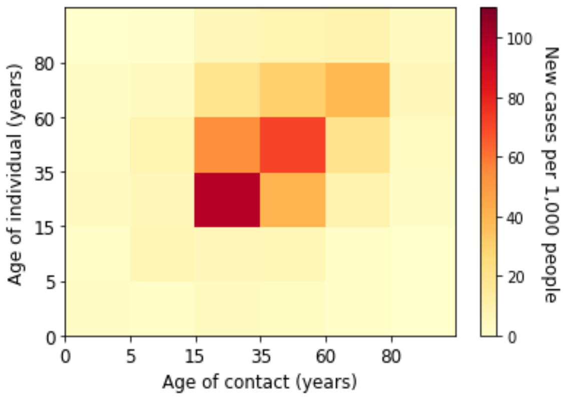


*(ag) Week 1 of 2021*

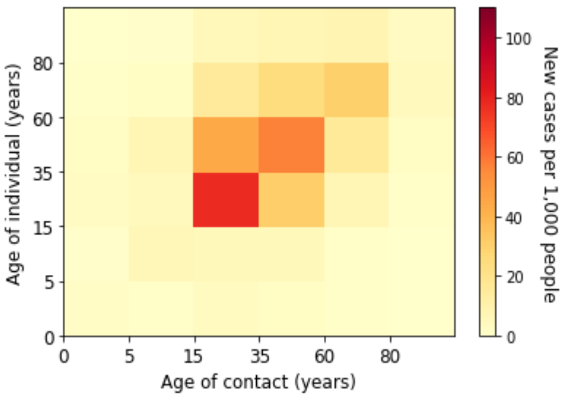


*(ah) Week 2 of 2021*

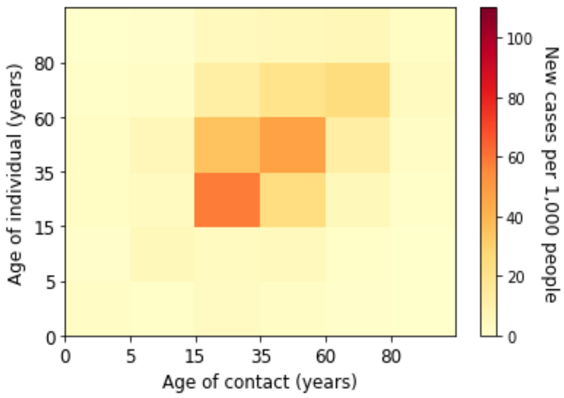


*(ai) Week 3 of 2021*

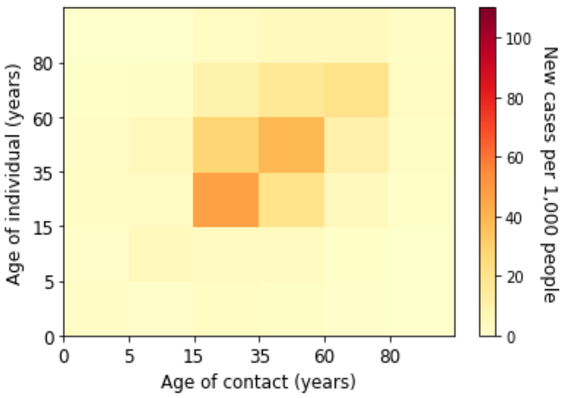


*(aj) Week 4 of 2021*

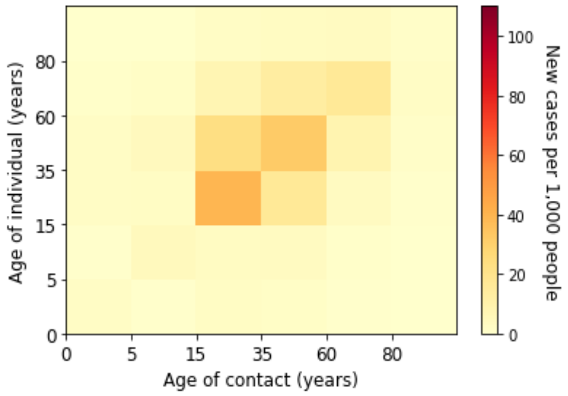


(ak) Week 5 of 2021

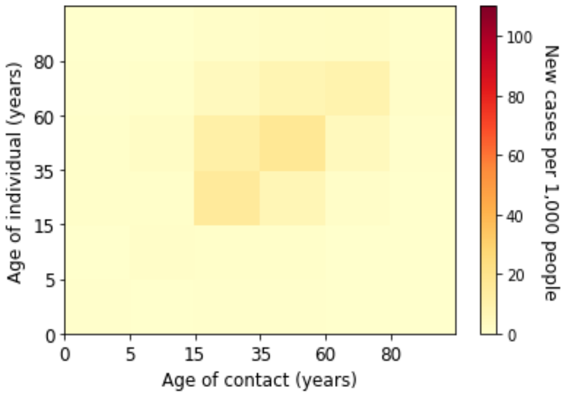


(al) Week 6 of 2021

*Figure 14. Estimated absolute contribution of transmission for each age groups in the second wave*

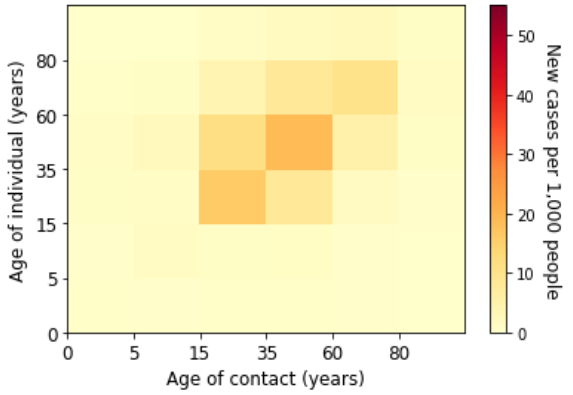


(a) Week 7 of 2021

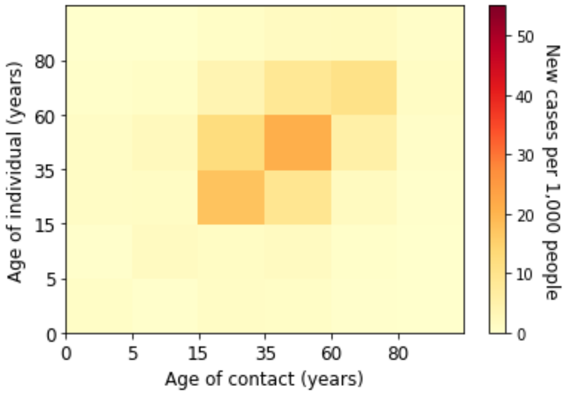


(b) Week 8 of 2021

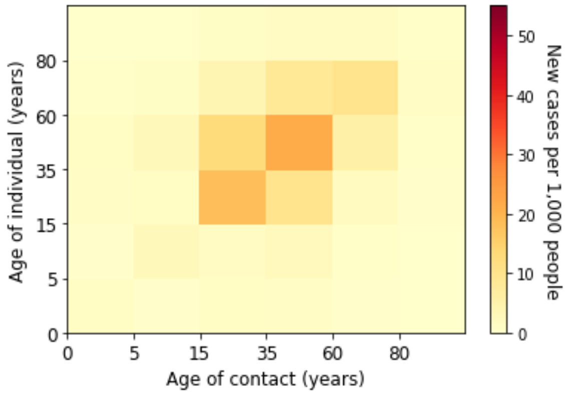


(c) Week 9 of 2021

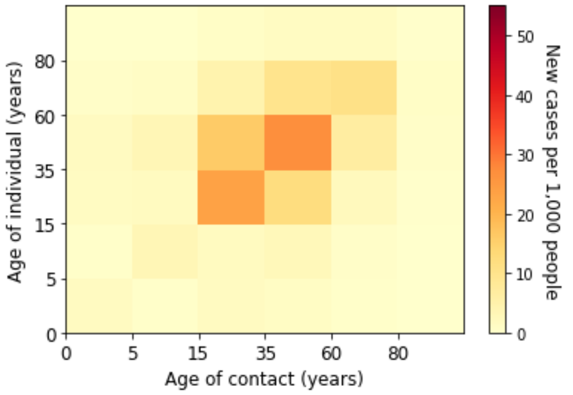


(d) Week 10 of 2021

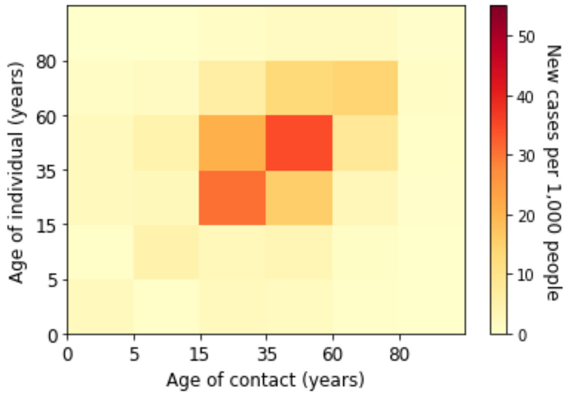


(e) Week 11 of 2021

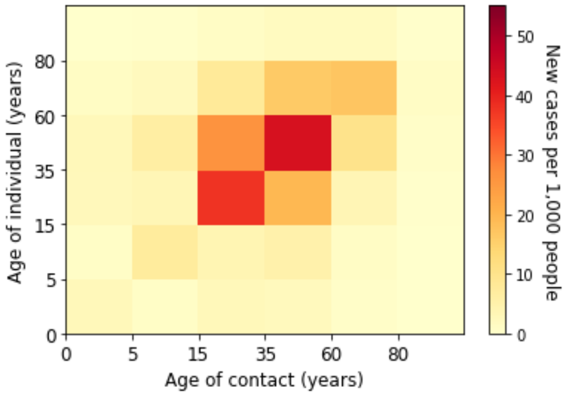


(f) Week 12 of 2021

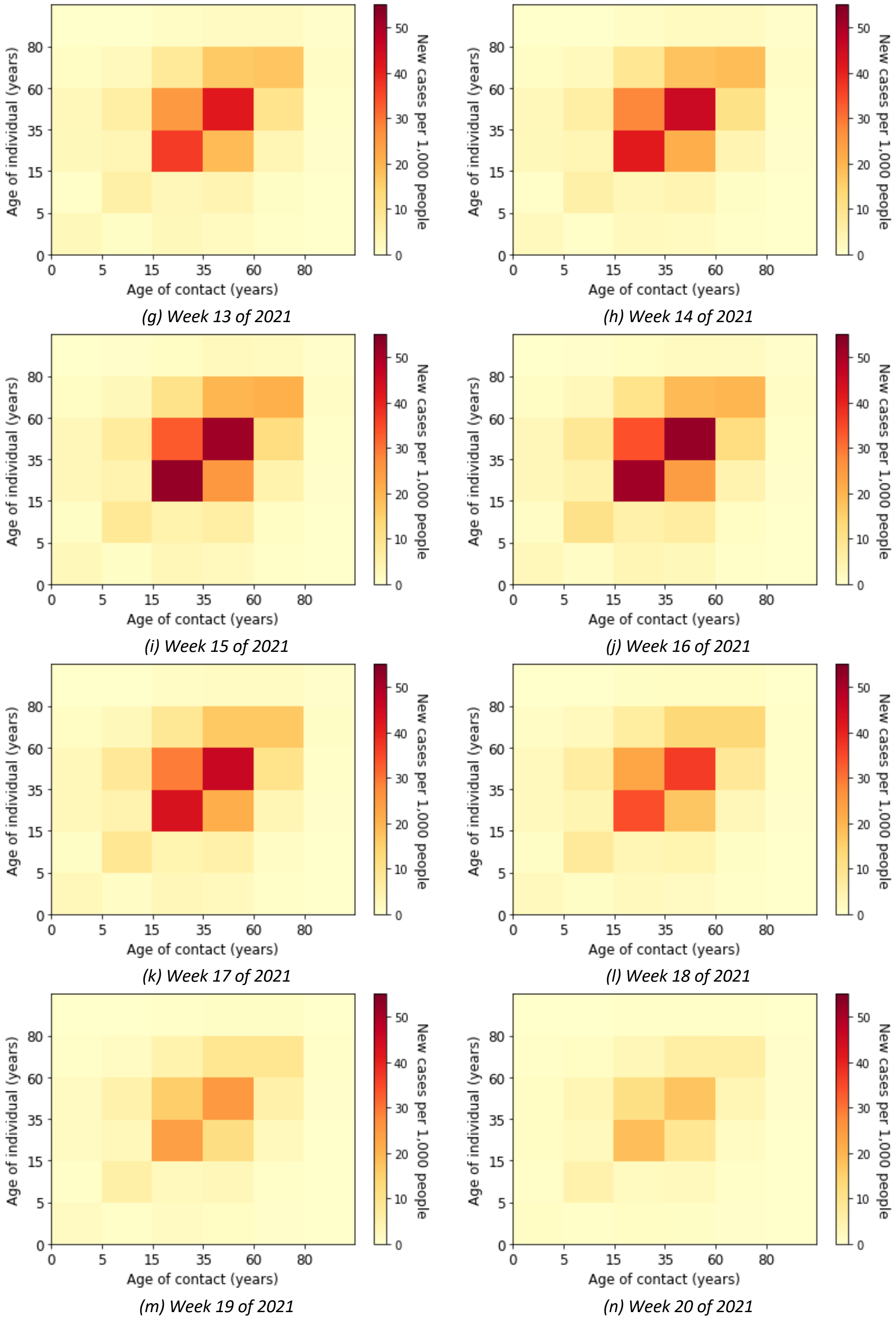


(g) Week 13 of 2021

(h) Week 14 of 2021

(i) Week 15 of 2021

(j) Week 16 of 2021

(k) Week 17 of 2021

(l) Week 18 of 2021

(m) Week 19 of 2021

(n) Week 20 of 2021

*Figure 15. Estimated absolute contribution of transmission for each age groups in the third wave*
